# Multi-model approach to understand and predict past and future dengue epidemic dynamics

**DOI:** 10.1101/2024.10.18.24315690

**Authors:** Cathal Mills, Francesca Falconi-Agapito, Jean-Paul Carrera, César V. Munayco, Moritz U. G. Kraemer, Christl A. Donnelly

## Abstract

Understanding the past, current, and future dynamics of dengue epidemics is challenging yet increasingly important for global public health. Using data from northern Peru across 2010 – 2021, we introduce a multi-model approach that integrates new and existing techniques for understanding and predicting dengue epidemics. Using wavelet analyses, we unveil spatiotemporal patterns and estimate space-varying epidemic drivers across shorter and longer dengue cycles, while our Bayesian hierarchical model allows us to quantify the timing, structure, and intensity of such climatic influences. For forecasting, as a single model is generally sub-optimal, we introduce trained and untrained probabilistic ensembles. In settings that mirror real-world implementations, we develop climate-informed and covariate-free deep learning forecasting models involving foundational time series, temporal convolutional networks, and conformal inference. We complement modern techniques with statistically principled training, assessment, and benchmarking of ensembles, alongside interpretable metrics for outbreak detection to disseminate outputs with communities and public health authorities. Our ensembles generally outperformed individual models across space and time. Looking forward, whether the public health objective is to learn from the past and/or to predict future dengue epidemic dynamics, our multi-model approach can be used to inform the decision-making of public health authorities.

## 1. Introduction

Dengue is a mosquito-borne disease which poses an increasingly global public health threat. Reported annual incidence has risen from 500,000 cases in 2000 to over 14 million cases in 2024^1^. 80% of the reported cases in 2023 occurred in the WHO Region of the Americas, yet dengue is endemic in over 100 nations and is expanding in distribution within and across nations ^2,3,4^. Primarily spread by *Aedes* mosquito vectors (female *Ae. aegypti* and *Ae. albopictus*), the viral infection is caused by the dengue virus (DENV), a orthoflavivirus with four genetically distinct serotypes and multiple lineages ^5^. Infection with an individual serotype offers long-term immunity to the specific serotype, but only short-term immunity to others ^6^. Symptoms of dengue vary in severity, and up to 80% of all infections are asymptomatic ^2^.

The increasing number of dengue outbreaks has been linked to human, viral, and environmental factors. Climatic conditions such as temperature, precipitation, and humidity influence the mosquito abundance and DENV replication, as *Aedes* mosquitoes thrive in warm, humid conditions and precipitation is necessary for a mosquito’s juvenile stages ^7,8,9,10^. Extreme climatic conditions, including drought and heavy precipitation, instigated by events such as the El Niño phenomena, have been identified as epidemic drivers ^11,12,13,14^. Meanwhile, modulating human factors like rapid urbanisation have allowed for the creation of mosquito breeding sites (e.g. water storage practices) and addition of susceptible hosts in close proximity, while increased travel, human mobility, and globalisation have similarly contributed to greater potential for rapid geographic spread ^15,16^.

Faced with a difficult-to-detect, climate-sensitive disease, public health authorities require a quantitative understanding of epidemic dynamics and drivers to inform their decision-making. In the absence of complete spatial, temporal, or population coverage of dengue surveillance, authorities may also need alternative ways to fill gaps in surveillance ^17^. Prospectively, public health authorities often aim to have early warning systems and predictive models of spread which can probabilistically predict future epidemic trajectories and the likelihood of outbreaks, thus allowing sufficient time for design of evidence-based public health policies ^18,19,20,21^. Therefore, in Brazil, South-east Asia, and Barbados, modellers have used wavelets, Bayesian hierarchical models, deep learning models, and ensemble frameworks to understand epidemic dynamics and forecast incidence and outbreaks. These approaches have separately enabled a model-based understanding of spatial patterns (e.g. highly correlated outbreaks and amplifying effects), temporal patterns (e.g. strong seasonality), demographic patterns (e.g. mean ages of infection), and key epidemic drivers (e.g. climatic and human influences) ^12,22,23,24,25,26^. In our studied region of northern Peru, dengue is a substantial, year-round public health burden where similar techniques have been used (separately) to quantify climatic effects and forecast short-term epidemic trajectories ^13,14,27^.

Here, we build upon published work, introducing an interdisciplinary modelling workflow (Figure 1). To date, modelling workflows have been presented for a single modelling discipline, yet are not dengue-specific nor enable evidence synthesis which uses a multi-model approach ^28,29,30,31^. Workflows also rarely integrate a multi-model approach that can learn from the past, monitor current, and predict future dynamics. As a model is an imperfect description of reality, there is no reason to assume (potentially restrictively) that a single technique for modelling is correct and appropriate for decision-making ^32,33^. Here, we acknowledge this fact to avoid drawing results and conclusions from a single modelling discipline, as our approach leverages strengths from statistical time series, wavelet analysis, Bayesian statistics, deep learning, and probabilistic ensemble forecasting. We demonstrate how our approach might be used in real-time to robustly analyse past, current, and future dengue epidemic dynamics. Using fourteen provinces in Peru as a case study we illustrate the potential utility of our approach, comparing it to more established dengue models by specifically focusing on the advances brought by integrating multiple techniques in terms of accuracy, reliability, and robustness. Our modelling outputs and general framework are generalisable and can be used across any context where dengue outbreaks occur frequently (endemic settings).

**Figure 1:**
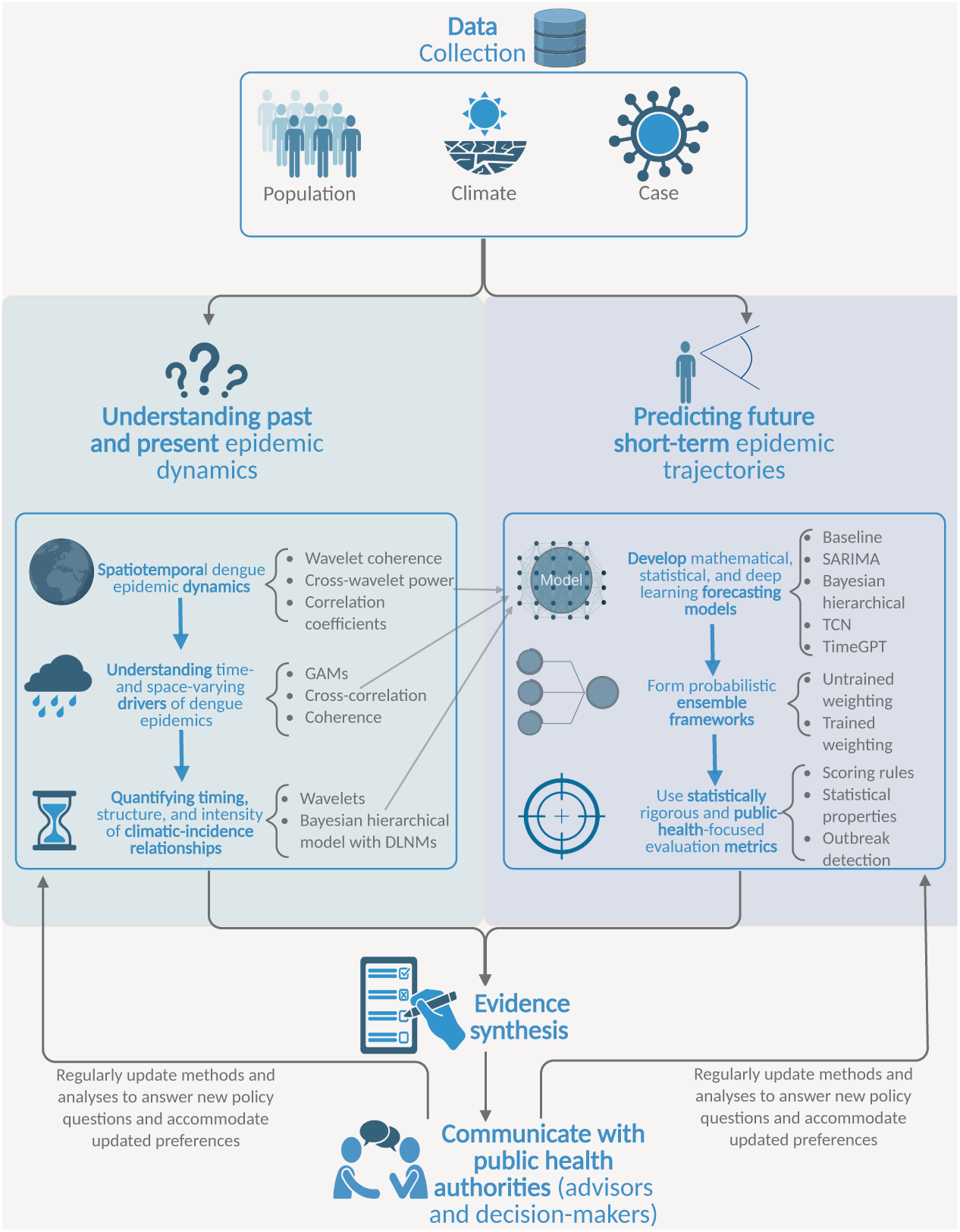
Modelling and forecasting workflow. Our multi-model workflow takes the researcher and public health authority from data collection to studying past, present, and future dengue epidemic dynamics. The workflow begins by sourcing heterogeneous data types. Our retrospective analyses use wavelets, Bayesian modelling, and other techniques to provide a quantitative understanding of past epidemic dynamics and drivers. This pillar (left) focuses on understanding dengue epidemics and may be traditionally regarded as exploratory analysis, yet we avoid this distinction as wavelets are not a model in the traditional sense yet are still a mathematical tool that requires specification of the type of wavelet and other hyperparameters. For forecasting future dynamics, we propose multi-model probabilistic forecast ensembles which predict new dengue cases with a horizon of up to one month, while our forecast evaluation involves both statistically rigorous (e.g. proper scoring rules) and public-health-focused criteria (e.g. outbreak detection metrics). The outputs of each stage of the retrospective analyses (left pillar) can inform the development of individual forecasting models. From the multi-model retrospective and forecasting analyses, evidence synthesis across space and time is a vital component for operationalising a useful workflow and for communicating the modelling outputs with different stakeholders in public health authorities (e.g. scientific advisors and decision-makers may require different levels of modelling details). See Sections 3 and 4 for an illustrative application and evidence synthesis for the workflow.

Epidemiological modelling for public health policy can be viewed through the lens of three axes; explanation, prediction, and decision analysis, and our framework enables explanatory and predictive insights from seemingly distinct approaches that aid decision-makers with monitoring and controlling dengue transmission. So, whether the objective is to understand (modelling climatic drivers) or predict (forecasting dengue cases), our framework enables robust conclusions and thus bridges some of the gaps in the literature regarding modelling and forecasting beyond single domains.

## 2. Materials and methods

### 2.1. Data

We used province-level reported dengue cases per month from May 2010 to December 2021 across the fourteen provinces in Piura, Tumbes, and Lambayeque (on the northern coast of Peru) which had registered large dengue outbreaks during this period, as per the National Centre for Epidemiology, Disease Prevention and Control (Peru CDC) in Peru’s Ministry of Health ^34^. The reported cases included both confirmed and probable cases across all serotypes (without serotype classification for each case). Probable dengue cases were defined as an individual with i) febrile illness for a maximum of seven days, ii) two or more specific symptoms, and iii) resides in or has recently visited areas with dengue transmission or known *Ae. aegypti* populations. Confirmed dengue cases met the same exposure and symptoms criteria, and had a positive dengue test ^35^. We aggregated weekly case data (using epidemiological weeks, starting from Sunday and ending on Saturday) to total monthly cases to align with other data sources (see below). In real-time applications of our framework, it is unclear the extent of delay (potentially varying across time and space) in reporting individual dengue cases in Peru, yet we suspect that the delay is 1–2 weeks and varies between health regions. Such a phenomenon which could impact the timeliness and utility of real-time modelling and forecasting analyses.

We interpolated population data from the 2007 and 2017 national censuses, and the 2022 mid-year population estimates ^36,37,38^. Then, we defined Dengue Incidence Rates (DIRs) per 100,000 by converting reported new dengue cases to rates per 100,000. Data on the proportion of provinces’ populations living in urban areas were also sourced from the 2007 and 2017 censuses but was not available more recently. We therefore used the 2017 data for years up to 2021.

Climatological data were as previously described in^[13]^ and we calculated average values for each province area. We sourced total monthly precipitation and monthly averages of daily maximum (and minimum) temperature from the WorldClim 2·1 dataset ^39,40^, a drought indicator called the Standardized Precipitation Index (SPI-6) ^41,42^ from the European Drought Observatory ^43^, and El Niño Southern Oscillation (ENSO) indicators, namely the El Niño Coastal Index (ICEN) and the Oceanic Niño Index (ONI) from the Geophysical Institute of Peru (IGP) ^44,45^ and the National Oceanic and Atmospheric Administration ^46^ respectively. Each of these climatological variables has been used in previous modelling studies in Peru and/or in other nations ^13,14,22,26,47^.

### 2.2. Wavelet and exploratory analyses

We developed wavelet methods to analyse epidemic dynamics and drivers. Wavelets have been used for dengue analyses in neighbouring Brazil and across Southeast Asia ^12,23,24^, and can analyse signals with sharp discontinuities and time-varying periodicity (i.e. non-stationarity). Using the R package Wavelet-Comp, we used a Morlet wavelet with non-dimensional frequency ω_0_ = 6^12,23,48,49^, which allowed us to decompose DIRs into reconstructed annual (maximum period of two years) and multiannual (period of two to twelve years) cycles. The wavelet transform produced a wavelet power spectrum, and we averaged across the provinces’ reconstructed annual and multiannual cycles to obtain the average wavelet power of the annual and multiannual cycles per province per month.

For both annual and multiannual cycles, to compare the relative timing of epidemics for each province pair, we computed the cross-wavelet power (the wavelet analogue of covariance) and wavelet coherence (the wavelet analogue of correlation). To measure epidemic synchrony (how the amplitudes of the incidence time series covary), for each province pair, we computed the pairwise Pearson correlation coefficient between their DIR time series and between their reconstructed (annual and multiannual) cycles. We also quantified time- and space-varying geographic, human, and climatic drivers via the cross-wavelet power and coherence between climatic conditions and reconstructed dengue cycles. We complemented this by developing generalised additive models (GAMs) for epidemic synchrony using smooth functions (thin-plate splines) of temporal random effects (month and year), spatial random effects (province), climatic variables (all variables above), and human mobility approximations (the pairwise product of provinces’ populations) ^50,51^. We fitted GAMs using the mgcv package in R version 4·2·1^48,51^, and evaluated model performance using the minimised generalised cross-validation score (GCV), Akaike Information Criterion (AIC) and Bayesian Information Criterion (BIC) ^52,53,54^.

Other exploratory analyses involved computing, for each province, the cross-correlation between past climatic conditions and current DIR at different time lags.

### 2.3. Bayesian climate-based modelling

We developed a Bayesian climate-based modelling framework which was similar to existing models from other nations ^22,26,47^ and our previous work in Peru ^13^. Here, we focus on recent modifications.

We modelled province-level dengue incidence using a zero-inflated Poisson distribution, thus accounting for anticipated excess zero counts. The final model, informed by exploratory analyses, was developed using data across 2010 to 2017, and contained: i) fixed effects (to estimate effects of recent incidence trends relative to the preceding year, shared seasonality across provinces, urbanised populations, and epidemic momentum), ii) province-level temporal effects (to estimate their monthly patterns and year-to-year heterogeneities), iii) spatiotemporal random effects via an adapted Bayesian version of the Besag-York-Mollié model ^55,56^ (to account for similarities and uniqueness across provinces), and iv) non-linear, delayed climatic influences via distributed lag non-linear models (DLNMs) ^57^. We provide additional model details in Supplementary Material 4.

We assessed models using information criteria (Widely Applicable Information Criteria, WAIC and cross-validated logarithmic score), in-sample predictive accuracy, and leave-one-time-point-out cross-validation (alongside prediction interval (PI) coverage and probabilistic calibration). We fitted the models using Integrated Nested Laplace Approximation (INLA) ^48,58^.

### 2.4. Probabilistic ensemble forecasting

By sampling from the posterior predictive distribution, the climate-based model directly enables probabilistic forecasts of dengue incidence with a forecast horizon of up to one month. However, individual models can have contrasting strengths and be wrong in different ways (e.g. predictive vs understanding transmission mechanisms). So, we consider ensemble frameworks which combine the forecasts of individual models. Ensemble frameworks have demonstrated strong predictive performance for many dynamical systems (e.g. COVID-19, seasonal influenza, dengue, and weather forecasting ^59,60,61,62^). Ensemble forecasts are often represented in one of two ways; using samples or quantiles of the predictive distribution. We focus on quantile-based forecasts as i) it provides access to a wider range of models which directly target quantiles (e.g. where there is no assumed likelihood), and ii) it is a computationally cheaper environment (versus processing thousands of samples). These have been used in recent years ^63,64^. Some ensemble approaches for dengue have not used probabilistic forecasts (e.g. ^65,66^), instead focusing on pointwise accuracy metrics which do not measure uncertainty in our predictions. These approaches also have not always separated the model development period from the testing period nor benchmarked performance using iteratively updating models, all of which is necessary to reflect a realistic application. In recent years, probabilistic ensembles have been increasingly used, often with equal weighting or trained weighting of sample distributions ^60,61,62,67^.

For the forecasts of dengue incidence at a one-month horizon, we used 23 quantile levels (identical to past forecasting studies ^64^). For untrained approaches, we used the median or mean average of individual modelling components’ predictive quantiles, while for trained approaches, we used linear stacking with the quantgen package in R ^68^ which weighted individual models’ quantile forecasts for a given month based on weights that optimised the sum of the quantile losses for forecasts over the past twelve months. We performed the trained weighting for each province independently (i.e. province-dependent weights) and for all provinces jointly (i.e. province-independent weights). We iteratively updated the model weights as each month’s data became available (i.e. a realistic online learning procedure).

While we performed our study retrospectively, to reflect a realistic application, we forecasted dengue cases with a one-month horizon using an expanding window which left all future data out of model training. The four-year period of January 2018 to December 2021 was our testing period. This allowed sufficient data for the model development period (2010 to 2017 inclusive), where we tested forecasting models with leave-future-out cross-validation (also known as time series cross-validation ^69^. We selected covariates of each model (Section 2.5) solely from performance of candidate models in the model development period.

To evaluate forecasts, using the scoringutils R package^[70]^, we applied the weighted interval score (WIS) – a proper scoring rule that approximates the continuous ranked probability score (CRPS) ^63,71^. The WIS measures the absolute distance between our predictive distribution and observed data, and simultaneously assesses model calibration (compatibility of forecasts with observations) and sharpness (concentration of predictive distributions). Lower scores indicate better performance. As the WIS accounts for distributional uncertainty, we avoided focusing excessively on pointwise accuracy metrics. It matters to our forecast assessment how we apply a scoring rule, and applying to case counts is likely inappropriate due to different epidemic properties across space and time ^72^. Here, we i) follow recommended procedures to score forecasts on a natural-logarithm-transformed scale (i.e. log(Cases + 1)), and ii) also score forecasts on a DIR scale. While we primarily focused on the recommended logarithmic scale, the latter assessment is motivated by a desire to be comprehensive and to provide interpretable, comparable information for public health authorities across space and over time. Alongside the WIS, we assessed bias, quantile coverage, and PI coverage, which measure respectively whether the model is systematically wrong in a given direction, whether the model’s quantile predictions are well-calibrated and whether the model’s prediction intervals (PIs) are well-calibrated. These statistical properties quantify reliability of forecasting models, and may guide forecasters and public health authorities when deciding whether a model can be employed for consistently, stable results.

While all such measures provided aggregate and statistically rigorous perspectives of performance, we also included a public-health-focused measure which focused attention on times of the year and areas of the predictive distribution of public health urgency. Here, we evaluated ability to correctly classify (i.e. binary forecast) the following month’s DIR reaching thresholds of 50 or 150 per 100,000. These thresholds were pre-specified by us to indicate different magnitudes of dengue outbreaks, and we report classifications which can be easily interpreted by public health authorities and the wider public. We used our model-based quantile forecasts to identify a cut-off quantile level (i.e. a decision rule based on historical performance) which determined whether we classify an outbreak (of DIR ≥ 50 or 150 per 100,000) for the next month. An outbreak is forecasted if the prediction at the (historically calibrated) cut-off quantile level exceeded the corresponding outbreak threshold (e.g. 50 per 100,000). We allowed decision rules to be iteratively updated as each month’s data became available – in each month, the rule was determined to maximise the historical AUC (the area under the receiver operating characteristic curve) for classifying outbreaks. This separated the inference stage (modelling) from the decision-making stage, and ensured that our classifications maximised public health utility (simultaneously maximising true positives and minimising false alarms avoids wasted resources and lack of public confidence). We repeated this outbreak detection evaluation for the first onsets of outbreaks, where we assessed performance for forecasts of initial yearly onset of outbreak with DIR ≥ 50 (or 150) per 100,000. This smaller dataset includes classifications for each province for months of a year corresponding to the outbreak onset, months before the onset, or in a year with no eventual onset. This classification focuses on a public health priority of dengue incidence *first* reaching pre-defined thresholds.

### 2.5. Our ensemble framework components

Our ensemble frameworks contained two or more forecasting model components whose predictive distributions (for one-month-ahead cases) were represented by predictive quantiles (23 levels as used in previous forecasting studies^[59,73]^).

Our baseline model was an epidemiologically naive model which forecasted next month’s dengue cases as the current month’s dengue cases. This was an application of an existing benchmarking model from the U.S. COVID-19 Forecast Hub, and is a random walk model with innovations (i.e. uncertainty) based on past differences in monthly dengue cases ^64^.

Our Bayesian climate-based model (described in Section 2.3) used information up to one month before the target month to generate samples from a posterior predictive distribution – from which we computed quantile forecasts. We also developed Seasonal Auto-Regressive Integrated Moving Average (SARIMA) models. These statistical time series models were fitted independently to the logarithm of monthly cases for each province independently using the Auto-ARIMA (Auto-Regressive Integrated Moving Average) implementation of the Statsforecasts package and Darts library in Python ^74,75^. This implementation selects the order of the ARIMA model based on the AIC ^52^, and quantile forecasts are generated by assuming normality in forecast errors. We refitted the model at each time point to reflect real-world conditions as new information becomes available. For the baseline, Bayesian, and SARIMA models, we generated quantile forecasts by taking quantiles of the 5,000 samples of the predictive distributions.

The models above are traditional model frameworks, yet in recent years, deep learning approaches have demonstrated strong forecasting performance for infectious diseases ^76^. Informed by our exploratory analyses, we introduce here the first known application of a foundational time series model for forecasting dengue cases. We used TimeGPT, a transformer-based foundational time series model with self-attention that was pre-trained on over 100 billion data points from different domains ^77,78^. We implemented this model (for the logarithm of cases) with and without climatic covariates (two-month rolling averages of precipitation and minimum temperature). The model readily produces quantile forecasts by using conformal inference — a distribution-free way to produce statistically rigorous PIs ^79^. We also developed a forecasting model that used a temporal convolutional network (TCN) from the Darts library in Python ^74^. This deep learning architecture has dilated causal convolutional neural networks which preserve the local and temporal structures of the data. To avoid making distributional assumptions, we used quantile regression to directly estimate the quantiles of the predictive distribution. We again used climatic covariates of two-month rolling averages of precipitation and minimum temperature. Covariates were based on exploratory analyses from the model development period.

We outline model abbreviations in Table 1 and additional details on ensemble forecasting models in Supplementary Material 5. We followed EPIFORGE 2020 guidelines ^80^ throughout the forecasting analysis (Table SI 3).

**Table 1:**
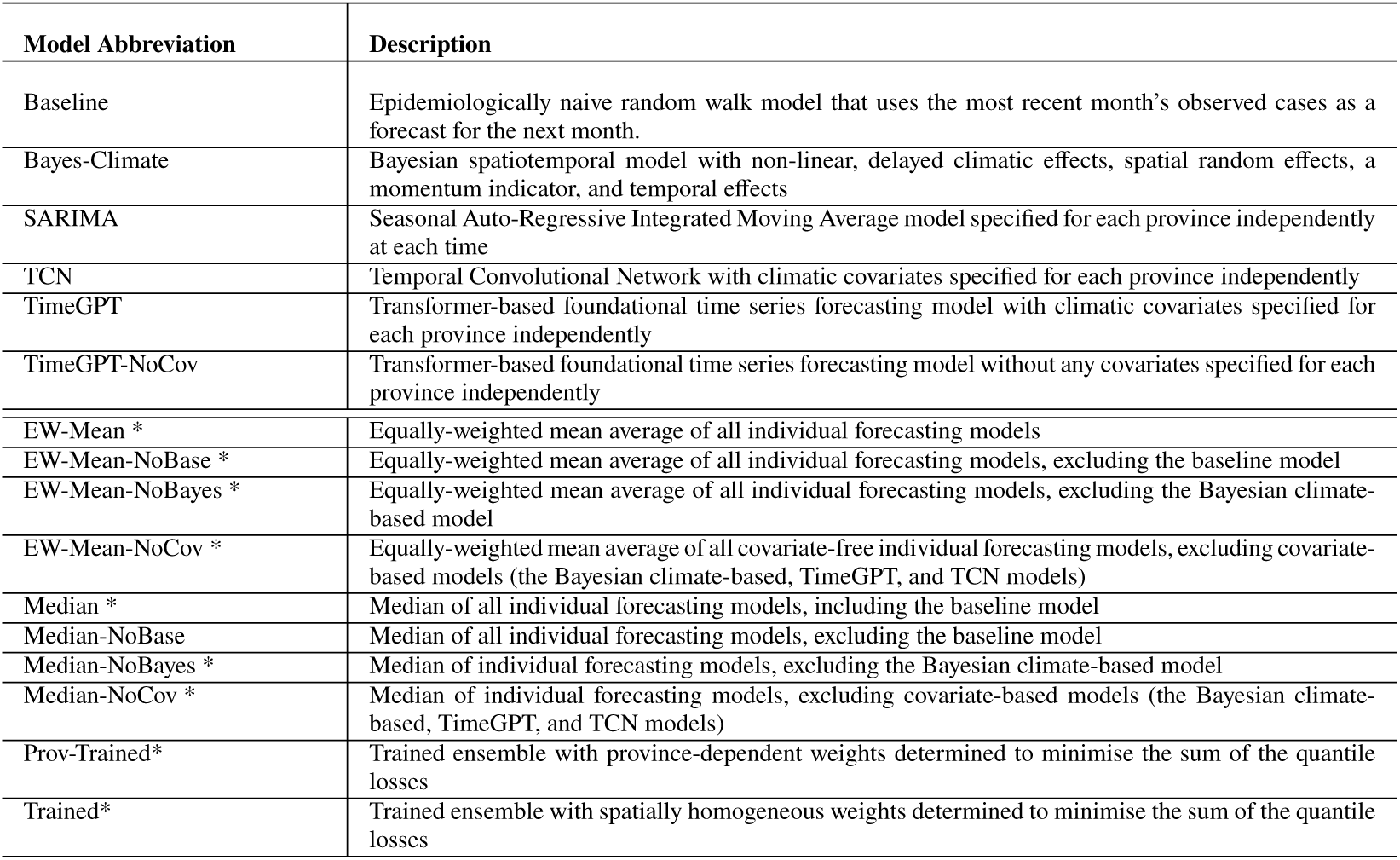
Quantile-based forecasting models and abbreviations. The six individual forecasting models are described first (above the double lines), followed by ensemble models that are highlighted with an asterisk. The equal weighting (EW) scheme applies equal weights to the quantile of that ensemble model’s individual predictions for each quantile. The median of individual models means that we take the median of the individual models’ predictions for each quantile.

## 3. Results

### 3.1. Investigation of epidemic dynamics using wavelets and climate-based analyses

We used wavelet analysis to simultaneously analyse signals (of incidence and climatic time series) in both time and frequency domains, focusing on reconstructions of annual and multiannual (two to five years) cycles of DIRs. Similar to dengue analyses from Southeast Asia ^12,24^, large dengue epidemic years (2015 and 2017) were characterised by statistically significant peaks in the average wavelet power and amplitude of reconstructed cycles (Figures 2, SI 3). Spatially, we found common large peaks across reconstructed cycles in provinces of the north and west, with slightly more widespread incidence peaks in the second large epidemic year of 2017. The mean period of multiannual cycles reduced during such years, while across the study period, we observed similar overall trajectories of the mean period (Figure SI 7) across the provinces (median pairwise Pearson correlation: 0.66, Interquartile Range – IQR: 0.25, 0.84). Complementary visualisations of monthly DIRs suggested strong similarity in the province-level dengue incidence, in terms of both timing and magnitude. Trends in dengue incidence included waves in the south preceding increases in provinces in the north and waves in the west preceding surges in the east. The trends were observed within individual years and when analysed using monthly averages over the study period (Figures SI 4, SI 5, SI 6).

**Figure 2:**
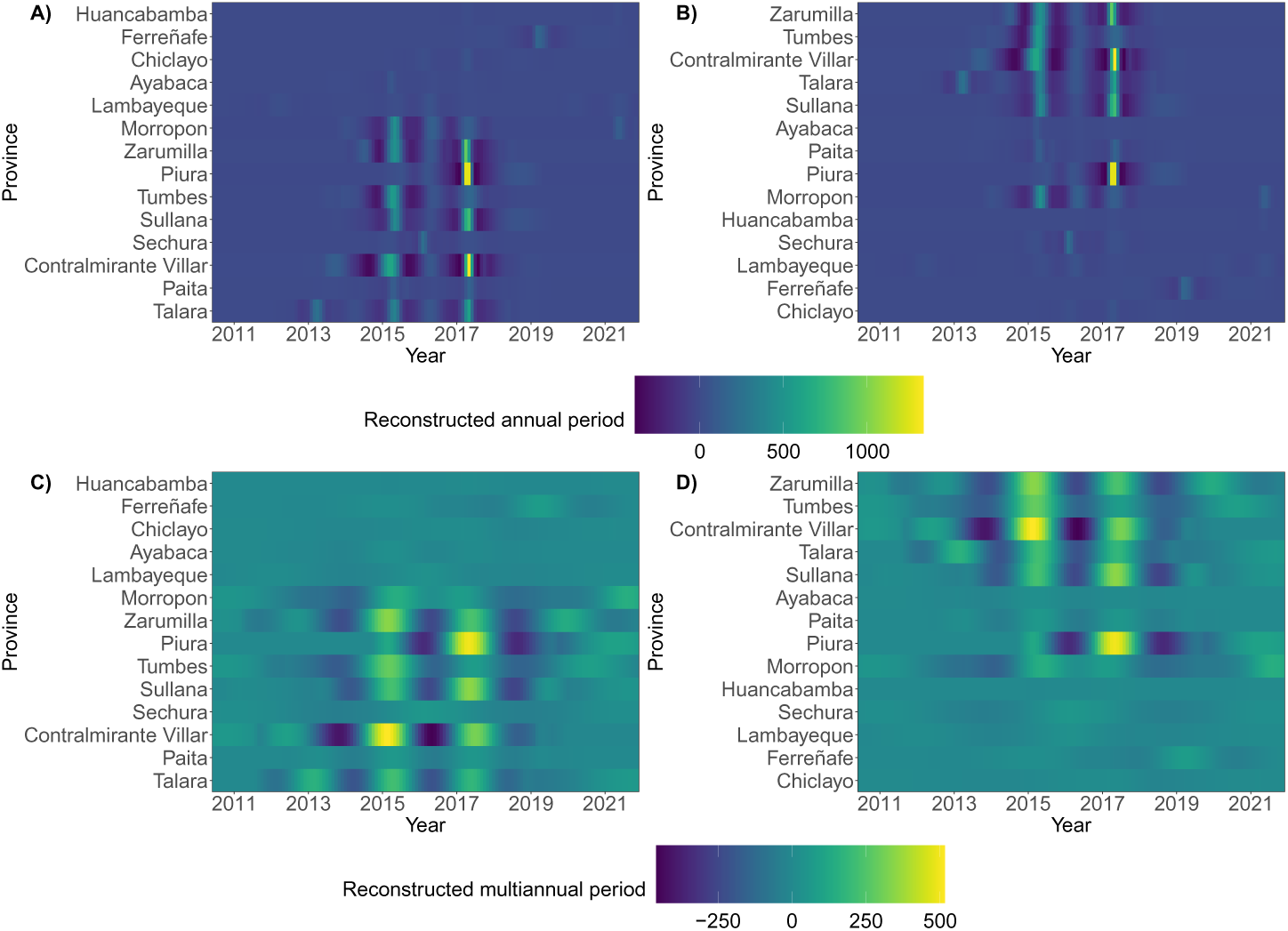
Wavelet reconstructions of cycles of Dengue Incidence Rate (DIR) time series. **(A)–(B)**: Reconstructed annual cycles of province-level DIRs, where provinces are sorted in ascending order by **(A)** longitude (from west to east) and **(B)** latitude (from south to north). **(C)–(D)**: Reconstructed multiannual cycles of province-level DIRs, where provinces are again sorted in ascending order by longitude and latitude respectively. Higher values of the annual (multiannual) period indicate peaks in DIR across the annual (multiannual) reconstructions, where the wavelet-based reconstructions are the reconstructions of province-level DIR time series using the initial decompositions into time-frequency and then, frequencies corresponding to annual (multiannual) cycles. Figure SI 2 depicts the reconstructed cycles with provinces sorted by percentage of population living in urban areas.

Wavelet coherence (measuring the relative timing of epidemics) was generally widespread across time and space (Table SI 5, Figures SI 11, SI 10). Coherence amplified (i.e. was more widespread) during epidemic years when more provinces shared statistically significant coherence with the (annual and multiannual) dengue cycles of other provinces (Figure 3). Spatially, more northerly and more westerly provinces tended to possess a greater proportion of statistically significant coherent relationships with annual and multiannual cycles of other provinces, whilst more southerly and easterly provinces had a larger number of out-of-sync (or equivalently, self-contained) cycles. We estimated these spatial trends at individual time points within years (Figure 3) and across the studied period of 140 months (Figure SI 11), yet a caveat is that Chiclayo (the most southerly province) had more coherent cycles, reflective of more urbanised provinces possessing more significantly coherent annual and multiannual cycles (Table SI 6, Figure SI 2).

**Figure 3:**
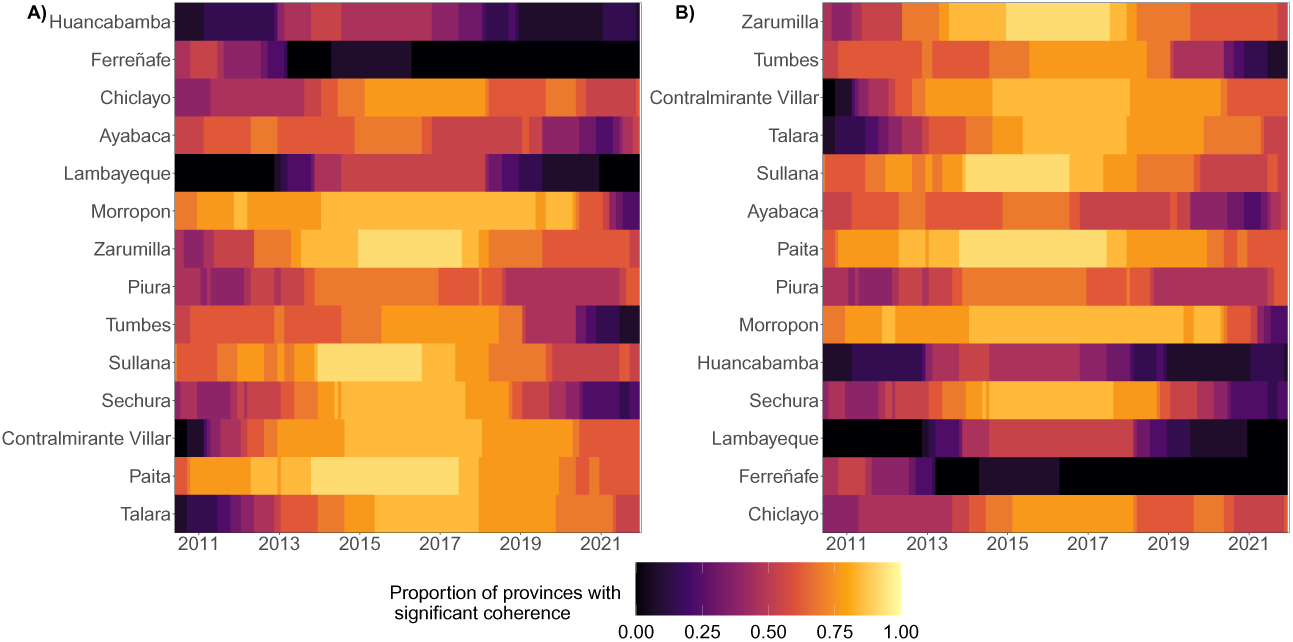
Coherence of reconstructed annual dengue cycles. Coherence is the wavelet analogue of correlation and here, compares the relative timing of provinces’ dengue epidemics. We visualise the proportion of a focal province’s annual dengue cycles which share statistically significant coherence with the cycles of other provinces at each time point. Provinces are sorted in ascending order by **(A)** longitude (from west to east) and **(B)** latitude (from south to north).

We found moderate-to-strong epidemic synchrony between annual raw DIR time series. Similar to coherence, there was strong synchrony between reconstructed cycles (both annual and multiannual, Table SI 7), and synchrony was high during large epidemic years (Figure SI 13). Spatially, particularly in reconstructed multiannual cycles, synchrony tended to be stronger in more northerly and westerly provinces (Figure SI 12), which indicates that correlations between epidemic curves shared similar patterns as the seasonality of epidemics (i.e. coherence).

Using coherence and cross-wavelet power, we quantified impacts of geographic, climatic, and human drivers on reconstructed cycles, and found widespread statistically significant coherence and consistently greater average cross-wavelet power between climatic variables (maximum temperature, precipitation, and ICEN) and dengue cycles (both annual and multiannual) during epidemic years of elevated incidence (Figure SI 14). Peaks in average cross-wavelet power were always observed during periods of elevated incidence across each of the climatic variables and across almost all of the provinces (Figures SI 15, SI 16, SI 17). More northerly provinces shared greater levels of statistically significant coherence with climatic variables. The ICEN had a greater coherence (with reconstructed cycles) in more westerly (i.e. coastal) provinces, and had a spatial gradient over time in the cross-wavelet power with annual cycles, as we estimated greater agreement with seasonality of cycles over time as one progresses from west to east and from north to south (Figure SI 17).

To assess the influence of lagged climatic conditions on human disease incidence and to inform model development, we computed cross-correlations between climatic variables and the raw monthly DIR time series (during the model development period; 2010 to 2017). There were generally moderate-to-strong, and spatially consistent, relationships between climatic conditions and DIRs across the provinces, particularly across temperature, drought, and precipitation. From a forecasting perspective, the strongest relationships were inferred with climatic lead times of one to three months, and cyclical patterns in the relationship were visibly pronounced (Figure SI 1).

We investigated the impact of pairwise distance between provinces on their corresponding average correlation and proportion of significant coherent cycles. Within some provinces, there were clear relationships between shorter distances and greater average correlation or coherence (which was most widespread for coherence between multiannual cycles). Shorter pairwise province distances were associated with similar annual epidemic dynamics (timing and amplitudes, across annual and multiannual cycles) in more northerly provinces (Figures SI 18, SI 19).

We extended previous modelling methods ^24^ (also incorporating the pairwise product of population sizes) as we developed GAMs to identify predictors of epidemic synchrony (see Materials and methods). Our best-fitting GAM, based on generalised cross-validation score and corrected AIC ^81,82^, included climatic influences (minimum temperature and precipitation), year-to-year heterogeneity, and the (logarithmic) pairwise product of population sizes. Here, positive (negative) coefficients indicate estimated enhanced (diminished) synchrony in the presence of the corresponding covariate. We estimated negative coefficients in twelve of the fourteen provinces for the pairwise distance-province interaction effect on epidemic synchrony, while six of the fourteen provinces had statistically significant (all negative) effects (Figure SI 20). Significant positive (partial) effects were also estimated for temporal (yearly) random effects during epidemic years, whilst lower (and higher) values of the pairwise product of populations were associated with statistically significant negative (and positive) effects on synchrony respectively. Climatic partial effects were subject to greater uncertainty, but greater minimum temperatures and greater precipitation were estimated to have increasingly negative and increasingly positive effects respectively (Figure SI 20 and see Discussion).

### 3.2. Bayesian climate-based modelling

The results from our climate-based Bayesian spatiotemporal model complement and enrich the details of our wavelet- and climate-based analyses, while such analyses above informed the form of our Bayesian model. During the model development period (2010–2017), we estimated strong model generalisability due to accurate estimated out-of-sample trends. The leave-one-time-point-out posterior predictive distributions’ 95% credible intervals contained 82.3% of the observations (indicating a reasonably calibrated model) and the corresponding posterior median explained 66% of the variation in the DIR (i.e. *R*^2^ = 0.66).

For model-based climatic inferences across the studied period (2010–2021), we analysed risk profiles (Figure SI 21), where relative risk of greater DIR was measured with respect to the risk induced by the mean average of each climatic variable. Higher (lower) values of the two-month-rolling-averaged, monthly average of daily minimum temperature were associated with elevated (diminished) relative risk of human disease incidence. We also estimated a consistent risk profile for the cumulative drought indicator with elevated DIR risk for extreme values (large surpluses or deficits of accumulated precipitation). Intermediate values of the indicator were associated with lower relative risk. In terms of the two-month rolling average of monthly precipitation, we estimated greater risk of disease incidence for values between 150mm and 200mm at lags of approximately one month, whilst extreme large precipitation values, except at very small time lags, were associated with diminished risk of DIR. Finally, we estimated a complex risk profile for the sea surface temperature indicator (ICEN) as extreme large values (El Niño events) and extreme small values (La Niña events) were each associated with both elevated and diminished risk at different time lags across four months. It was difficult to infer the mechanisms behind some of the complex behaviour in the risk profile for the ICEN (see Discussion).

### 3.3. Ensemble forecasting

We deployed our Bayesian model in an expanding window, forecasting environment (excluding current and all future months’ data from model training) for each month in the model development period (2010 to 2017), and found inferior results versus the leave-one-time-point-out analysis. Irrespective of assessment on a logarithmic or DIR level (see Materials and methods), we identified model calibration issues due to frequently underpredicting posterior predictions and hence, poor interval and quantile coverage values (Figures SI 28, SI 29). This suggested the Bayesian climate-based model alone would likely be inappropriate and unreliable for probabilistic forecasting, and supported our motivation for ensemble frameworks. While it would be unwise (due to bias from model tweaking after seeing the data) to focus heavily on results from the model development period, we found stronger performance for each of our other ensemble candidate members (see Section 2.5), including assessments of the whole predictive distribution (the WIS), pointwise accuracy (e.g. *R*^2^), PI and quantile coverage, and bias (Table SI 8). For example, the simple yet flexible SARIMA models improved the calibration (e.g. quantile and interval coverage in Figures SI 28, SI 28), the accuracy of pointwise predictions (e.g. *R*^2^ = 0.79, see Table SI 8 and Figure SI 23), and the overall distributional accuracy (WIS). Our deep learning models, particularly TimeGPT (the foundational time series model) which incorporated climatic covariates, also yielded strong forecasting performance relative to the baseline forecaster, the models without climatic covariates, and the Bayesian climate-based model (Figures SI 22-SI 27, Table SI 8). Simple, untrained ensembles also consistently produced accurate pointwise estimates and strong calibration and sharpness.

Our four-year testing window of 2018 to 2021 was an environment to assess performance in a setting reflective of a real-world application, as all models (including hyperparameters, covariates, and ensemble weights) were pre-defined from only historical data. We found strong predictive performance in forecasting cases for each of our ensemble models (Tables 2, SI 9), as captured by WIS, PI and quantile coverage, and pointwise accuracy metrics (*R*^2^ and RMSE – Root Mean Square Error). We found similar strong performance of ensemble models for forecasting the annual first onset of DIR reaching thresholds of 50 (or 150) per 100,000 (Figure 4 a), and more generally for forecast classification of months with DIR above/below these thresholds (Figure 4 b). The individual models handle different types of uncertainty in different ways (e.g. uncertainty in data generating processes, random error terms, parameter estimates etc.), yet individually do not account for all possible sources of uncertainty and thus, the coverage of their 95% PIs is less than the claimed coverage (95%). In contrast, relative to other ensembles and individual models, ensembles based on the median of individual models’ quantiles produced statistically valid PIs and consistently strong forecasting ability (Figure 4). This was captured by consistently sharp and well-calibrated (albeit relatively conservative) PIs (Figure SI 41), low bias, and better WIS values (e.g. pairwise comparisons of WIS values in Figure SI 37, Table 2). We observed these trends across provinces (Figure 5 e) and over time (Figure 4 c), and for different magnitudes of epidemics (Figure SI 39 and Table SI 15). The superiority of forecasts from ensemble models, particularly median-based ensembles, was also observed when we ranked models on their forecasts of DIRs. Across assessments on logarithmic and DIR scales, median-based ensembles consistently dominated the model rankings based on score ratios, outbreak detection, and pointwise accuracy metrics (Tables 2, SI 9, SI 12, SI 13, Figures 4 c, 5 a and SI 37).

**Figure 4:**
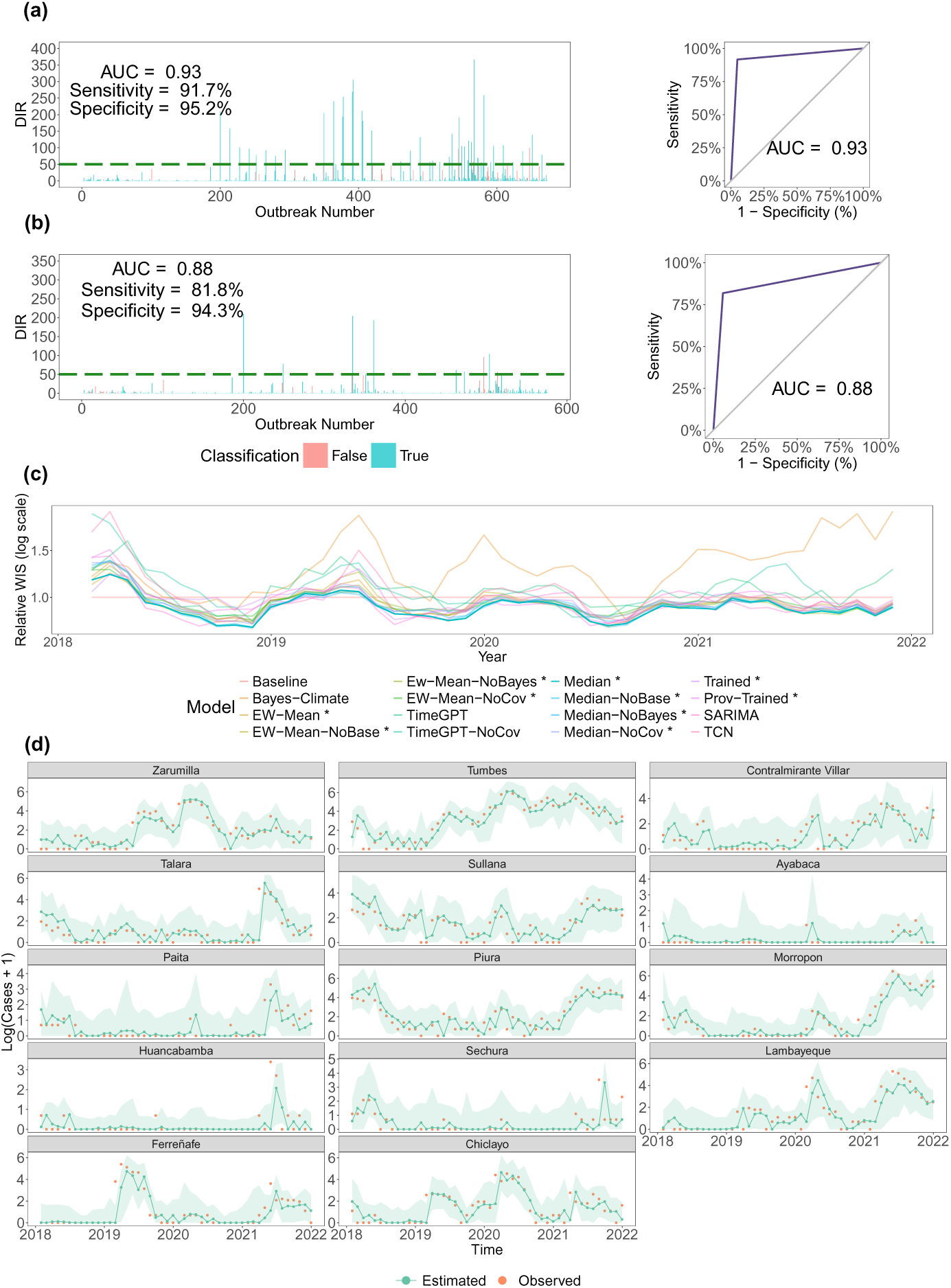
Performance of median ensemble forecasting model: This ensemble forecasting models’ probabilistic forecasts were generated by taking the median of all six ensemble forecasting models’ prediction at the 23 quantile levels (see Materials and methods). (a): Visualisations of forecasting ability for classifying whether the coming month will experience a DIR of ≥ 50 per 100,000. AUC denotes the area under the receiver operating characteristic (ROC) curve, and higher values indicate greater classification skill. (b): Analogous visualisation for classifying whether the coming month will be the first month of the year in a province with DIR of ≥ 50 per 100,000. (c): Relative Weighted Interval Score (WIS for log(Cases + 1)) is shown for all forecasting models in centred, rolling windows of three months, with the median model’s values highlighted. Relative WIS is an individual forecasting models’ WIS divided by the baseline forecasting’s WIS, where values below one indicate greater forecasting skill relative to the baseline forecaster. The corresponding summary statistics for log(Cases + 1) over time are visualised in Figure SI 39. (d): For each province (sorted in descending order by latitude from north to south), we visualise the median model’s predictions (for log(cases + 1)) where green points denote the posterior median and orange points denote the observed values. The green shaded intervals denote the 95% prediction intervals (PIs). 95.2% of all observations lay within these 95% PIs. The posterior median explained 74% of the variation in log(cases + 1) i.e. *R*^2^ = 0.74. Y-axes are province-dependent due to the varying sizes of the dengue epidemics observed.

**Figure 5:**
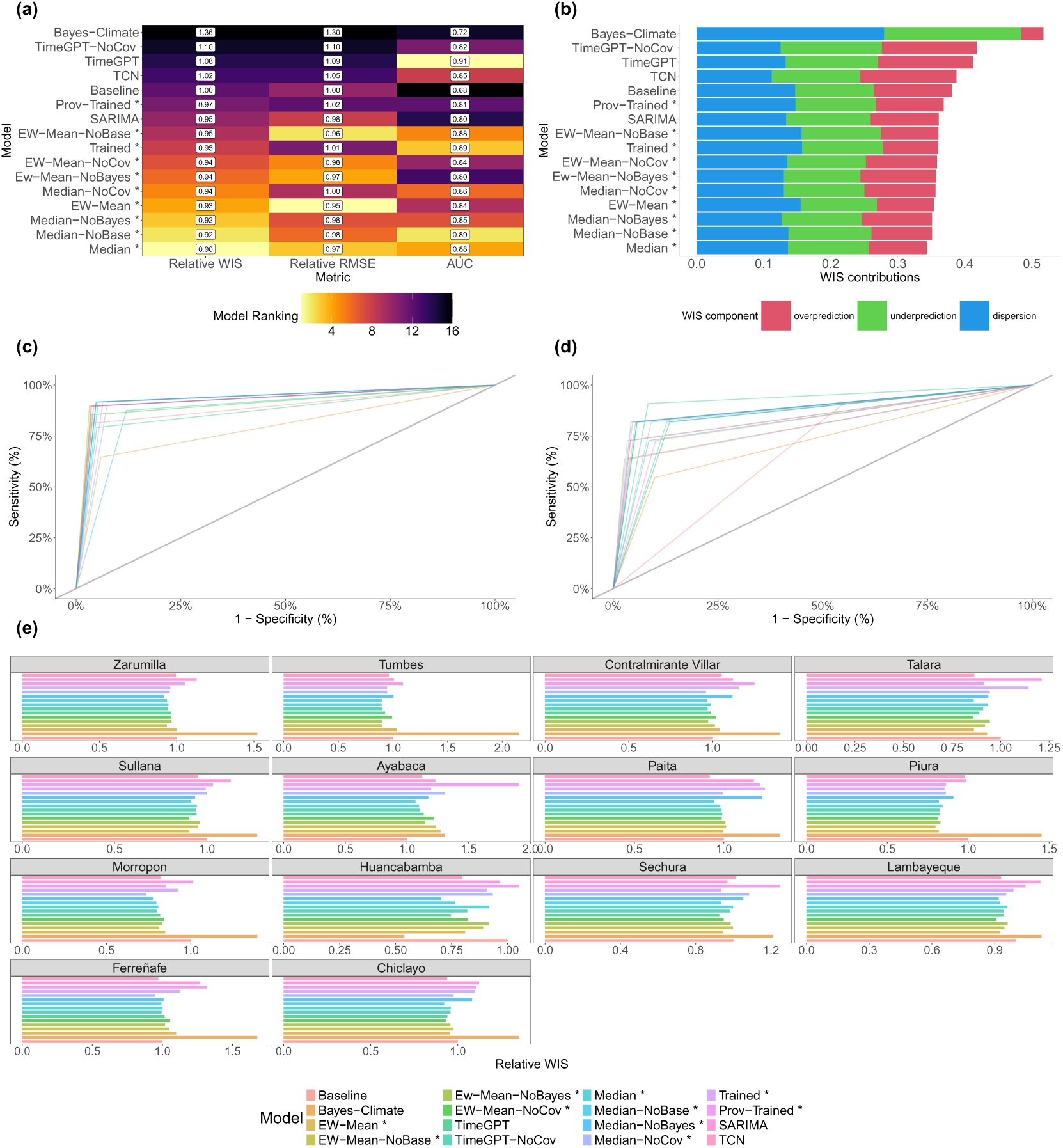
Comparing relative performance of forecasting models in the testing period of 2018 to 2021. We compare performance of our 16 ensemble frameworks (see Table 1 for abbreviations) in terms of their predictive performance for forecasting log(Cases + 1). (a) Model rankings by different metrics, where relative root mean squared error (RMSE) and mean relative Weighted Interval Score (WIS) are defined relative to the baseline model. Lower values indicate greater predictive performance for these relative metrics, but higher AUC values indicate greater forecasting skill for correctly classifying the first months with DIR ≥ 50 per 100,000 one month ahead of time. (b): WIS is decomposed into overprediciton, underprediction, and dispersion for each of the forecasting models. (c): Receiver operating characteristic (ROC) curves by forecasting model for correctly classifying months with DIR ≥ 50 per 100,000 one month ahead of time. (d): Analogous ROC plot to (c) for detection of first month with DIR ≥ 50 per 100,000 one month ahead of time. (e): Relative WIS is shown for all forecasting models per province where provinces are sorted in descending order by latitude from north to south. Relative WIS is an individual forecasting models’ WIS divided by the baseline forecaster’s WIS for that province. X-axes are province-dependent due to the varying sizes of the dengue epidemics observed.

**Table 2:**
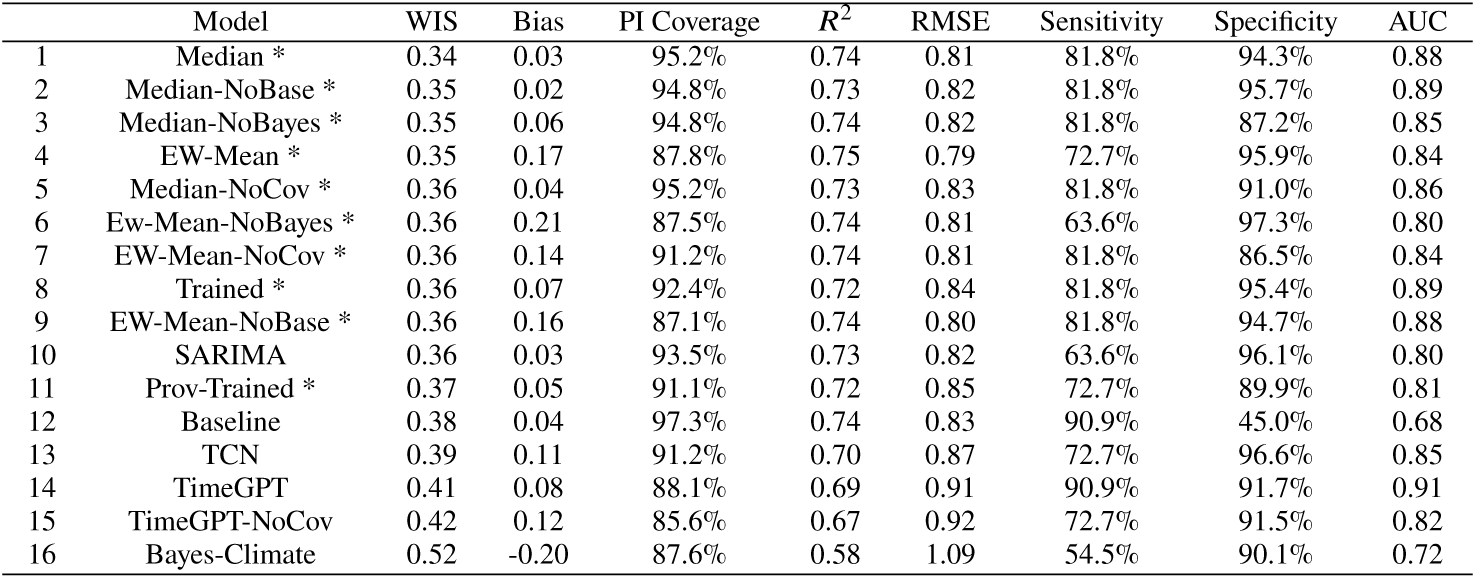
Different models’ results for forecasting log(Cases + 1) in the testing period of 2018 to 2021. We summarise proper scores, predictive performance, calibration, and outbreak detection capabilities of forecasting models. Models are arranged by mean weighted interval score (WIS), from best (lowest) to worst (highest). Further summary statistics for WIS are presented in Table SI 14, and implied similar model rankings. Bias measures the bias of each models’ predictive quantiles, PI coverage measures the coverage of the 95% prediction intervals (PIs), *R*^2^ denotes the proportion of variation in log(Cases + 1) explained by the models’ posterior median estimates, and RMSE is the corresponding root mean square error (RMSE) for log(Cases + 1) by using the posterior median. For detection of outbreak onset in each province in each year, *Sensitivity*, *Specificity*, and *AUC* represent the True Positive rate, True Negative rate, and Area Under the Curve in terms of forecasting skill for correctly classifying the first possible outbreak of DIR ≥ 50 per 100,000. These metrics are visualised in Figure 5 a for model comparison purposes. Table SI 9 is the analogous table which depicts outbreak detection for DIR first surpassing a threshold of 150 per 100,000, while Table SI 12 presents the models scored on a DIR level. Details of the model abbreviations are provided in Table 1 (where similar model rankings were obtained). All ensemble models are highlighted with an asterisk.

| | Model | WIS | Bias | PI Coverage | $R^2$ | RMSE | Sensitivity | Specificity | AUC |
| --- | --- | --- | --- | --- | --- | --- | --- | --- | --- |
| 1 | Median * | 0.34 | 0.03 | 95.2% | 0.74 | 0.81 | 81.8% | 94.3% | 0.88 |
| 2 | Median-NoBase * | 0.35 | 0.02 | 94.8% | 0.73 | 0.82 | 81.8% | 95.7% | 0.89 |
| 3 | Median-NoBayes * | 0.35 | 0.06 | 94.8% | 0.74 | 0.82 | 81.8% | 87.2% | 0.85 |
| 4 | EW-Mean * | 0.35 | 0.17 | 87.8% | 0.75 | 0.79 | 72.7% | 95.9% | 0.84 |
| 5 | Median-NoCov * | 0.36 | 0.04 | 95.2% | 0.73 | 0.83 | 81.8% | 91.0% | 0.86 |
| 6 | EW-Mean-NoBayes * | 0.36 | 0.21 | 87.5% | 0.74 | 0.81 | 63.6% | 97.3% | 0.80 |
| 7 | EW-Mean-NoCov * | 0.36 | 0.14 | 91.2% | 0.74 | 0.81 | 81.8% | 86.5% | 0.84 |
| 8 | Trained * | 0.36 | 0.07 | 92.4% | 0.72 | 0.84 | 81.8% | 95.4% | 0.89 |
| 9 | EW-Mean-NoBase * | 0.36 | 0.16 | 87.1% | 0.74 | 0.80 | 81.8% | 94.7% | 0.88 |
| 10 | SARIMA | 0.36 | 0.03 | 93.5% | 0.73 | 0.82 | 63.6% | 96.1% | 0.80 |
| 11 | Prov-Trained * | 0.37 | 0.05 | 91.1% | 0.72 | 0.85 | 72.7% | 89.9% | 0.81 |
| 12 | Baseline | 0.38 | 0.04 | 97.3% | 0.74 | 0.83 | 90.9% | 45.0% | 0.68 |
| 13 | TCN | 0.39 | 0.11 | 91.2% | 0.70 | 0.87 | 72.7% | 96.6% | 0.85 |
| 14 | TimeGPT | 0.41 | 0.08 | 88.1% | 0.69 | 0.91 | 90.9% | 91.7% | 0.91 |
| 15 | TimeGPT-NoCov | 0.42 | 0.12 | 85.6% | 0.67 | 0.92 | 72.7% | 91.5% | 0.82 |
| 16 | Bayes-Climate | 0.52 | -0.20 | 87.6% | 0.58 | 1.09 | 54.5% | 90.1% | 0.72 |

While the assessments of WIS values (and other statistical measures) provide statistically rigorous measures of the whole predictive distribution, we also focus on the insights of the forecasts for public health authorities. For instance, the epidemiologically naive baseline model’s predictive distributions could be relatively close in distribution to the observed cases (if true distributions do not change drastically in short periods), yet this model fails to offer valuable information about oncoming outbreaks (as it forecasts the past observed cases). So, looking at detection of the annual first onset of DIR reaching thresholds of 50 (or 150) per 100,000 (Figures 4 b, 5 a and 5 c), the median-based ensemble models produced AUC values of between 0.85 and 0.89, which are i) far superior to the naive baseline forecasting model (AUC: 0.68) and ii) superior to individual models. For example, if authorities used the median-based ensemble of all individual models, 81.8% of first onsets (of reaching DIR of 50 per 100,000) would have been correctly detected one month ahead of time with a corresponding false alarm rate of 5.7%. We found similarly strong performance of ensemble models for classifying whether DIR in a province in any given month would be at or above the thresholds of 50 (or 150) per 100,000. Here, nearly all forecasting models demonstrated strong classification skill (e.g. AUC of between 0.79 for DIR ≥ 50 per 100,000, see Figure 5b and Table SI 10), likely in part due to the easier nature of predicting the next month being at/above a threshold given that we know the most recently observed high DIR level.

In terms of further relative comparisons, forecasts of ensemble models which included climatic covariates outperformed the analogous models without covariates (denoted NoCov in e.g. Table 2), while trained ensembles were outperformed by untrained median-based ensembles (see Discussion) across both logarithmic and DIR scales (Figure 5 a). Ensemble models also outperformed almost all individual forecasting models (e.g. 95% PI coverage and WIS), and model rankings were similar across DIR and logarithmic scales (Tables 2, SI 9).

## 4. Discussion

We have introduced a new multi-model framework (Figure 1) which enables multi-model evidence synthesis; as a researcher can explain the dynamics and drivers of past epidemics and robustly estimate likely future short-term epidemic trajectories.

### 4.1. Wavelet analysis and climate-based modelling

Using wavelet methods allows us to identify epidemic dynamics and drivers that vary in different temporal resolutions, times of the year, and/or in different geographies. We found strong similarities in both the seasonality and epidemic curves across the provinces, and similarities were pronounced during epidemic years with major outbreaks. It is unsurprising that northerly provinces, which experienced higher DIRs, possessed more synchronous and more coherent (annual and multiannual) dengue incidence cycles. Also, the strongest wavelet coherence between climatic conditions and dengue cycles occurred in more westerly and northerly provinces (all of which were coastal provinces). Consistent with wavelet analyses in Southeast Asia ^12^, such significant peaks in wavelet coherence and cross-wavelet power were estimated during years with larger epidemics, and were widespread, all of which is indicative of an amplifying role (greater epidemic similarity) played by climatic forcing in large epidemic years. The spatially shifting impact of the ICEN during such years (from west to east and north to south) may reflect initial climatic risks imposed on coastal provinces when an El Niño (or La Niña) event initially occurs, dengue incidence climbs, and subsequent climatic conditions and human mobility induce transition of outbreaks further inland and further south. From a policy perspective, in future work, the roles played by both climatic forcing and spatially shifting influences could be combined with seasonal climate forecasts (e.g. GloSea5^83^ was used for dengue forecasting in Vietnam ^60^) to enhance early warning detection. For the causes of epidemic synchrony between provinces, we identified large outbreak years, greater pairwise products of populations and shorter pairwise distances as having enhancing effects. This may reflect greater mobility and greater connectivity driving enhanced epidemic synchrony. Spatially, shorter pairwise distances had more significant effects on both synchrony and coherence in more northerly provinces, which may be indicative of greater connectivity (although this may be confounded by similarity in other environmental conditions). The more uncertain estimated effects of climatic conditions on synchrony may be due to our limited spatial focus (only fourteen provinces) and/or potential confounding in our models of synchrony.

The estimated elevated risk for anomalous precipitation deficits and accumulations is aligned with the well-documented impacts of water shortages and heavy rainfall ^22,84,85^. Drought-induced precipitation shortages, particularly in areas with poor water supply, can induce greater risk of dengue incidence due to changes to water storage practices (including more containers around the home), creation of potential egg-laying sites (for female *Ae. aegypti* who prefer artificial water storage containers), greater survival capacity of *Ae. aegypti* eggs in dry conditions, and/or more concentrated breeding in the few remaining suitable habitats ^86,87,88^. Similar elevated drought-induced risk has been identified in both Vietnam and Brazil ^22,47^. Meanwhile, the effects of heavy rainfall are mainly due to large-scale accumulation of stagnant water, which yields creation of abundant breeding sites, and increases to vector populations. The risk of disease incidence was estimated to peak after a one-month lag for the two-month average of monthly precipitation, whilst we estimated diminished risk for the heaviest precipitation (above a two-month average of 200m). Diminished risk has been uncovered in Asian nations including Singapore, Malaysia, Sri Lanka, and Thailand ^89,90^. Our final climatic variable (the ICEN) was estimated to have a complex risk profile. Whilst the estimated diminished risk for extreme indicator values at short time lags make intuitive sense (due to time taken for onshore climatic conditions of provinces to eventually change), protection (from risk) for greater absolute values at longer time lags seem counter-intuitive. The latter issue may due to the limited size of our data and/or using our predictive model to disentangle real-world climatic inferences (which may be subject to potential confounding). The issue of potential confounding (between variables) is applicable to each climatic variable, whilst external, unobserved confounders may also have affected our conclusions and thus, we indirectly estimated such unobserved confounding effects (via unstructured spatial effects and yearly random effects).

There are several limitations to both our retrospective and forecasting analyses. These include concerns about the quality, volume, and type of data available, ranging from a lack of information on reporting rates to a lack of serotype data (See Supplementary Material 8 for further discussion). We attempt to compensate for potentially changing serotypes and reporting rates with time-varying assessments of climatic-incidence relationships in our wavelet analyses, year-to-year random effects in our Bayesian model, and consistent updating of all hyperparameters in each forecasting model (including ensembles). Nevertheless, consistent with the WHO’s cited lack of data on circulating serotypes ^91^, we advocate for future collation of age-structured dengue cases and viral sequencing data, alongside future seroprevalence surveys across at-risk areas, all of which would provide insights about immunology and virology issues (such as multitypic immunity, genomic diversity, and repeat infections) across space and time. Similarly, while often logistically and economically challenging to collect, including mosquito abundance data (conditional on appropriate alignment of likely different spatial scales^[92,93]^) would ideally allow us to build a more mechanistic model and enhance exploratory power. Data regarding the role of the human are sometimes overlooked yet vital for vector-borne transmission, and given the high spatial synchrony between provinces, we also advocate for enhanced availability of human mobility data over time. Understanding how continuous water supply modulates climatic-incidence relationship is another important investigation that would be enabled by further data. Raising the necessary caveat that more data would not guarantee enhanced accuracy/representativeness^[94]^, quantifying the additional value of these data streams (e.g. using Value of Information analyses) for different policy questions (Figure 1) and different settings will be important for encouraging the potentially costly data collection in resource-limited settings.

### 4.2. Forecasting analysis

Moving from past to future dynamics, in a four-year testing window, we demonstrate a new probabilistic ensemble framework for forecasting monthly dengue cases up to one month ahead of time.

The reliable forecasts from ensemble frameworks demonstrate benefits of combining the strengths of multiple models from different disciplines. Each of these models are "wrong" in different ways, and yet each can handle different aspects of forecasting dengue cases. The foundational time series model (TimeGPT) included in the current work leverages strengths from a transformer architecture, conformal inference, and training on over 100 billion data time points. The TCN model uses representation learning and dilated causal layers in its deep neural network architectures. In doing so, these deep learning models can explicitly account for temporal structure (from seasonality to heteroscedasticity) and handle non-linearities in the data. While deep learning models can be computationally expensive, here, there are little to no additional (e.g. versus Bayesian hierarchical models) computational barriers for operationalising these models in potentially resource-limited settings as all our computations were performed locally on a laptop (with each provinces’ monthly forecasts produced in under one hour, irrespective of model). However, the fact that deep learning models do not always outperform simple-yet-flexible and robust SARIMA models is interesting and perhaps unsurprising. Dengue epidemics are somewhat predictable in terms of their highly seasonal patterns (e.g. dengue cycles based on climatic conditions) and their very short-term trajectories with highly autoregressive patterns (e.g. we expect the next month’s DIR to somewhat similar to the current month’s DIR). So, unlike some previous studies which either benchmark against models only within the deep learning field or do not allow the SARIMA model’s hyperparameters to be updated as new information becomes available, our rigorous benchmarking reveals similar performance of these statistical time series models with deep learning methods (which may have increased propensity to overfit due to the far larger numbers of model parameters). The similarity in performance is implied by comparable (or superior) values for PI coverage, bias, WIS, *R*^2^, and outbreak detection metrics. Similar results have been observed for forecasting of COVID-19 cases and deaths ^95^ and in other unrelated forecasting tasks with strong, seasonal patterns (such as retail demand and wholesale food prices) ^96,97^. Of course, this observation comes with the caveat that future work should assess how this predictive performance varies at longer forecast horizons and finer spatial and temporal resolutions (e.g. weekly), all of which are of great importance to public health authorities. We think that our extensive model and ensemble development represents a stepping stone towards such additional objectives. Future research could consider methods for achieving multi-step forecasts (e.g. recursive posterior predictive sampling, direct multi-step forecasts, and sequence modelling for Bayesian, statistical time series, and deep learning transformer-based models respectively) and/or incorporating new and increasingly accurate short and medium-term weather forecast data (which often use artificial intelligence, e.g. diffusion models or graph neural networks) to expand dengue case forecast horizons^[98,99,100]^.

Importantly, however, our deep learning models do always outperform the Bayesian climate-based model and improve our ensembles’ forecasts when included alongside other individual models. The latter trend is reflective of a more general observation of the outperformance of ensemble models, compared to individual models (similar to previous forecasting studies for COVID-19, influenza, and dengue ^61,64,101^). Intuitively, similar to our retrospective analysis, it is unreasonable to expect a single approach (wavelets and Bayesian models also have differing strengths and weaknesses) to capture all aspects of current or future epidemic dynamics. This is often due to data distribution shifts, and learned patterns may therefore vary substantially across space and/or evolve over time. These variations may be why our province-specific deep learning forecasting models (TCN and TimeGPT) with climatic covariates and statistical time series model (SARIMA) forecasted more reliably than our semi-mechanistic Bayesian climate-based model which estimated parametric (climate-to-incidence) relationships for all provinces and time points jointly. These trends are similar to previous poorer short-term predictive performance of models with mechanistic structure (and stronger performance of ensembles) in analyses of forecasting new dengue and COVID-19 cases ^61,95^, and may capture the difficulties in using mechanistic models for forecasting imperfect (noisy, underreported, and delayed) case data. Future work may consider pairing our framework with nowcasting approaches which can infer realised events which have not been reported yet (see ^102,103,104,105^ for recent methods and applications). Despite periods of inferior performance relative to the baseline model (Figure SI 39), the forecasting ensembles (e.g. median-based ensemble) generally provided more stable performance over time and across space (Figure 5), both during periods of low and high dengue incidence. We deduce that a single model alone is likely insufficient for answering all of our research questions, yet we can answer different research questions robustly by using and where appropriate, unifying, several models. Our aim here was not to include every modelling approach in a single framework, but rather to identify which models/approaches were appropriate for different tasks (e.g. probabilistically explaining past epidemic drivers vs predicting future trajectories).

Our forecasting results are not an upper bound on what is possible, as i) we have retained a preference for realism in our independent testing window (and we could also incorporate future covariates), ii) our pipeline can readily incorporate many more exploratory and forecasting models, and iii) future work could investigate further feature selection, optimal training windows for individual models and for weighting of models, as well as model weights per individual quantile. These future works may help to improve the performance of the ensemble in provinces which experience rare, sporadic dengue cases (e.g. Ayabaca). Our focus here has instead been to showcase that we can, for any dengue epidemic, blend, rather than overemphasising, the benefits of forecasting from single modelling domains.

Our work also highlights the importance of not focusing on a single, potentially misleading performance metric. We must consider what information the forecasting metric provides for public health authorities. While used for forecast evaluation in many dengue studies to date (e.g. ^65,66^), pointwise metrics obscure the pervasive uncertainty when we predict the unknown future of a dengue epidemic. We could, for example, obtain a high *R*^2^ by just predicting the most recent value, while providing little useful information for public health authorities. Similar to other recent forecasting studies (e.g. ^59,60,63,64^), we therefore propose proper scoring rules which measure distributional accuracy (WIS) and account for deficiencies of pointwise metrics by simultaneously evaluating calibration and sharpness. This is why we focus on WIS, yet not exclusively, as i) we could again obtain reasonably good WIS values by simply forecasting past value(s) (as drastic predictive distributional shifts may be rare in short time horizons) without conveying much information, and ii) we aim for our outputs to be easily interpreted and used by public health authorities. For example, for quick space- and time-specific forecasting model comparisons, we signpost Figures 4 d and 5 e as visually helpful guides. All of this is an important contribution of our work, as we complement proper scoring rules (which measure the whole distribution) with both relative WIS values (to reflect forecasting skill above the baseline) and measures of how well our forecasting models can detect specific aspects of epidemics, such as season onset. The WIS (and/or relative WIS) may be a statistically principled measure for our evaluations, yet when communicating our outputs, such a metric may matter so little to so many. In particular, the WIS approximates the CRPS – the simplest single measure of the overall value of a forecasting model for the entire uniform family of potential users. Ideally, metrics should also be interpretable, actionable, and public-health-focused (e.g. user- and/or event-focused), and so our outbreak threshold metrics refocus evaluation on areas of the predictive distribution that are vital for decision-making. These new outbreak thresholds (and the associated optimisation procedures) are simple yet generalisable across geographies of different sizes and across different spatial resolutions (e.g. coarser or finer resolution analyses).

In terms of further public health implications, it is interesting that we can still often obtain reliable probabilistic forecasts without climatic covariates. This may be related to the one-month forecast horizon as given that we know the current level of the epidemic, the next month’s cases may not require the climatic information for reliable forecasts. On the other hand, when making longer-term seasonal forecasts and inferring epidemic drivers as in our retrospective analyses, climatic covariates are crucial (e.g. ^60^). Another benefit of our approach is the computationally cheap nature of our forecasting pipeline. Working with 23 predictive quantiles per observation to summarise the predictive distribution saves a modeller from processing potentially thousands of predictive samples. Finally, ensembles with untrained weighting outperform those which use trained weighting, likely in part due to weight uncertainty and/or the restricted spatial and temporal focus of our study. More recent trained weighting approaches should be investigated in future work^101^.

The next step will be operationalising this multi-model approach (Figure 1) into a decision support tool for the Peru CDC – a computational tool that iteratively downloads data and produces retrospective analyses, probabilistic forecasts, and associated metrics. We emphasise that the co-development of this tool with the Peru CDC does not end after detailed retrospective and forecasting analyses. Subsequent steps for operational success will include synthesising evidence, determining the necessary details for communicating modelling results, and regular dialogue for updating models and co-defining new policy questions. Maintaining such collaboration and clarity of audience will be crucial for satisfying the 3-U (useful, usable, and used) requirements of a practical, successful digital tool^[106]^. This may be aided by adopting lessons from policy-modelling experiences in previous infectious disease settings (e.g. COVID-19 and mpox^[107,108]^). In doing so, we believe that our framework and recommendations (possibly paired with decision theory, see ^33^) can satisfy the necessary usefulness principle for a digital prediction tool ^106^, yet future work (e.g. surveys and involvement of social scientists) could explore i) usability; whether our custom outputs are interpretable (for different users involved at different stages of the decision-making process), and ii) usage; evaluation of effectiveness.

### 4.3. Conclusions

The key advance of our research is to introduce a new framework and multi-model way of evidence synthesis; robustly analysing dengue epidemic dynamics across space and over time. We introduce several state-of-the-art methods across different disciplines (in standalone and unified frameworks), yet their usage alone is not our objective, as we complement these methods with traditional, reliable modelling techniques to produce a computationally cheap and generalisable multi-model approach that emphasises public health utility throughout. The outputs of our retrospective and forecasting techniques are designed to be interpretable, robust, and comprehensive, and can be used to inform long-term public health planning and real-time decision-making for dengue epidemics.

## Acknowledgements

C.M. is supported by a studentship from the UK’s Engineering and Physical Sciences Research Council. FFA is funded by the Consejo Nacional de Ciencia, Tecnología e Innovación Tecnológica (CONCYTEC) and the Programa Nacional de Investigación Científica y EstudiosAvanzados (PROCIENCIA) within the framework of the call “E067-2023-01 Proyectos Especiales: Proyectos de Incorporación de Investigadores Postdoctorales en Instituciones Peruanas” with contract number No. PE501084391-2023.

J.P.C is supported by grants from the National Secretary of Science FID-2024-146 and FID-2024-147. M.U.G.K. and C.A.D. acknowledge funding from the Oxford Martin School Programme in Digital Pandemic Preparedness. Grants from the National Secretary of Science FID-2024-146 and FID- 2024-147. M.U.G.K. acknowledges funding from The Rockefeller Foundation (PC-2022-POP-005), Google.org, the Oxford Martin School Programme in Pandemic Genomics, European Union’s Horizon Europe programme projects MOOD (No. 874850) and E4Warning (No. 101086640), Wellcome Trust grants 303666/Z/23/Z, 226052/Z/22/Z & 228186/Z/23/Z, the United Kingdom Research and Innovation (No. APP8583), the Medical Research Foundation (MRF-RG-ICCH-2022-100069), UK International Development (301542-403), the Bill & Melinda Gates Foundation (INV-063472) and Novo Nordisk Foundation (NNF24OC0094346). C.A.D. is supported by the UK National Institute for Health Research Health Protection Research Unit (NIHR HPRU) in Emerging and Zoonotic Infections in partnership with Public Health England (PHE), Grant Number: HPRU200907. The funders had no role in the study design, analysis, or interpretation of results.

## Declaration of Interests

The authors declare no competing interests.

## Data Availability Statement

The dengue incidence surveillance data are publicly available from the National Centre for Epidemiology, Disease Prevention and Control (Peru CDC) in Peru’s Ministry of Health. We sourced these from https://www.dge.gob.pe/salasituacional. Mid-year population estimates are made available by the National Institute of Statistics and Information of Peru at https://www.inei.gob.pe/media/ MenuRecursivo/indices_tematicos/proy_04.xls. The WorldClim monthly historical climate data are available at https://www.worldclim.org/. SPI-6 data from the European Drought Observatory are available at https://jeodpp.jrc.ec.europa.eu/. The El Niño indices of the ONI and ICEN are available respectively from the NOAA (https://origin.cpc.ncep.noaa.gov/) and the Geophysical Institute of Peru (http://met.igp.gob.pe). All code and data used in our analysis are available at https://github.com/cathalmills/peru_dengue_province/.

## Notes

### Competing Interest Statement

The authors have declared no competing interest.

### Funding Statement

C.M. is supported by a studentship from the UK's Engineering and Physical Sciences Research Council.
M.U.G.K. acknowledges funding from The Rockefeller Foundation (PC-2022-POP-005), Google.org,
the Oxford Martin School Programmes in Pandemic Genomics & Digital Pandemic Preparedness,
European Union's Horizon Europe programme projects MOOD (No. 874850) and E4Warning (No.
101086640), the John Fell Fund, a Branco Weiss Fellowship and Wellcome Trust grants 225288/Z/22/Z,
226052/Z/22/Z & 228186/Z/23/Z, United Kingdom Research and Innovation (No. APP8583) and the
Medical Research Foundation (MRF-RG-ICCH-2022-100069). The contents of this publication are the
sole responsibility of the authors and do not necessarily reflect the views of the European Commission or the other funders. C.A.D. is supported by the UK National Institute for Health Research Health Protection Research Unit (NIHR HPRU) in Emerging and Zoonotic Infections in partnership with Public Health England (PHE), Grant Number: HPRU200907. The funders had no role in the study design or analysis.

### Author Declarations

The dengue incidence surveillance data are publicly available from the National Centre for Epidemiology, Disease Prevention and Control (Peru CDC) in Peru's Ministry of Health. We sourced these from https://www.dge.gob.pe/salasituacional.

### Summary of Updates

All sections of the manuscript have been updated, including further details on our methods, our results in the local context, and the discussion of the next steps for an operational decision support tool.

