## Supplementary material for "Multi-model approach to understand and predict past and future dengue epidemic dynamics"

### Supplementary Information

#### Supplementary Material 1. Acronyms

Table SI 1: **Summary of acronyms used throughout the analysis.**

| Abbreviation | Definition |
|---|---|
| ***Ae. Aegypti*** | *Aedes Aegypti* |
| **ACF** | Autocorrelation Function |
| **AUC** | Area Under the Curve |
| **BYM** | Besag, York and Mollié |
| **CRPS** | Continuous Rank Probability Score |
| **CV** | Cross-Validated |
| **DIC** | Deviance Information Criterion |
| **DIR** | Dengue Incidence Rate |
| **DLNM** | Distributed Lag Non-linear Models |
| **ENSO** | El Niño Southern Oscillation |
| **ICEN** | El Niño Coastal Index |
| **IGP** | Geophysical Institute of Peru |
| **INEI** | National Institute of Statistics and Information of Peru |
| **INLA** | Integrated Nested Laplace Approximation |
| **IQR** | Interquartile Range |
| **ONI** | Oceanic Niño Index |
| **prec** | Precipitation |
| **Q1** | Lower Quartile |
| **Q3** | Upper Quartile |
| **ROC** | Receiver Operating Characteristic |
| **RMSE** | Root-Mean Square Error |
| **RR** | Relative Risk |
| **RSI** | Relative Strength Index |
| **RW** | Random Walk |
| **SARIMA** | Seasonal Auto-Regressive Integrated Moving Average |
| **SPI-6** | Standardized Precipitation Index (6-Month) |
| **SST** | Sea Surface Temperature |
| **tmax** | Maximum Temperature |
| **tmin** | Minimum Temperature |
| **WIS** | Weighted Interval Score |
| **WHO** | World Health Organization |

#### Supplementary Material 2. Ensemble model abbreviations

##### *Supplementary Material 2.1. Forecasting models*

Table SI 2: **Quantile-based ensemble framework abbreviations used in figures and tables.**

| Model Abbreviation | Description |
|---|---|
| Baseline | Epidemiologically naive random walk model that uses the most recent month's observed cases as a forecast for the next month. |
| Bayes-Climate | Bayesian spatiotemporal model with non-linear, delayed climatic effects, spatial random effects, a momentum indicator, and temporal effects |
| SARIMA | Seasonal Auto-Regressive Integrated Moving Average model specified for each province independently at each time |
| TCN | Temporal Convolutional Network with climatic covariates specified for each province independently |
| TimeGPT | Transformer-based foundational time series forecasting model with climatic covariates specified for each province independently |
| TimeGPT-NoCov | Transformer-based foundational time series forecasting model without any covariates specified for each province independently |
| EW-Mean * | Equally-weighted mean average of all individual forecasting models |
| EW-Mean-NoBase * | Equally-weighted mean average of all individual forecasting models, excluding the baseline model |
| EW-Mean-NoBayes * | Equally-weighted mean average of all individual forecasting models, excluding the Bayesian climate-based model |
| EW-Mean-NoCov * | Equally-weighted mean average of all covariate-free individual forecasting models. This excludes covariate-based models – the Bayesian climate-based, TimeGPT, and TCN models. |
| Median * | Median of all individual forecasting models |
| Median-NoBase | Median of all individual forecasting models, excluding the baseline model |
| Median-NoBayes * | Median of individual forecasting models, excluding the Bayesian climate-based model |
| Median-NoCov * | Median of individual forecasting models, excluding covariate-based models (the Bayesian climate-based, TimeGPT, and TCN models) |
| Prov-Trained* | Trained ensemble with province-dependent weights determined to minimise the sum of the quantile losses |
| Trained* | Trained ensemble with spatially homogeneous weights determined to minimise the sum of the quantile losses |

#### Supplementary Material 3. EPIFORGE 2020 guidelines

Table SI 3: **Checklist for EPIFORGE 2020 guidelines:** We provide details of our reporting for the checklist items of EPIFORGE 2020[80].

| Section of Manuscript | # | Checklist Item | Reported on Page |
|---|---|---|---|
| Title/Abstract | 1 | Describe the study as forecast or prediction research in at least the title or abstract. | 1 |
| Introduction | 2 | Define the purpose of study and forecasting targets. | 1-2 |
| Methods | 3 | Fully document the methods. | 3-7 |
| Methods | 4 | Identify whether the forecast was performed prospectively, in real time, and/or retrospectively. | 5 |
| Methods | 5 | Explicitly describe the origin of input source data, with references. | 3 |
| Methods | 6 | Provide source data with publication, or document reasons as to why this was not possible. | 17 |
| Methods | 7 | Describe input data processing procedures in detail. | 3 |
| Methods | 8 | State and describe the model type, and document model assumptions, including references. | 4-7 |
| Methods | 9 | Make the model code available or document the reasons why this is not possible. | 17 |
| Methods | 10 | Describe the model validation and justify the approach. | 4-7 |
| Methods | 11 | Describe the forecast accuracy evaluation method used, with justification. | 5-6 |
| Methods | 12 | Where possible, compare results to a benchmark or other comparator model, with justification of comparator choice. | 6 |
| Methods | 13 | Describe the forecast horizon, with justification of its length. | 5 |
| Results | 14 | Present and explain uncertainty of forecasting results. | 10-11 |
| Results | 15 | Briefly summarize the results in nontechnical terms, including a nontechnical interpretation of forecast uncertainty. | 10-11 |
| Results | 16 | If results are published as a data object, encourage a time-stamped version number. | n/a |
| Discussion | 17 | Describe the weaknesses of the forecast, including weaknesses specific to data quality and methods. | 15-16 |
| Discussion | 18 | If the forecast research is applicable to a specific epidemic, comment on its potential implications and impact for public health action and decision-making. | 16-17 |
| Discussion | 19 | If the forecast research is applicable to a specific epidemic, comment on how generalizable it may be across populations. | 16-17 |

n/a= Not Applicable

#### Supplementary Material 4. Climate-based model: Additional details

The following section provides additional details to the description of the modelling framework presented in Section 2.3.

##### Supplementary Material 4.0.1. Model formula

Let $y_{it}$ be the reported province-level ($i = 1, \dots 14$) monthly ($t = 1, \dots 140$)) case counts of dengue from 2010 to 2021. Then, $y_{it}$ was modelled with a Zero-Inflated (ZI) Poisson distribution to account for the likely presence of excess zero counts in the province-level spatial resolution.

$$\begin{aligned} y_{it} &\sim ZIPoisson(\lambda_{it}, \pi) \\ \log(\lambda_{it}) &= \log(P_{it}) + \log(\eta_{it}) \\ \log(\eta_{it}) &= \beta_0 + \boldsymbol{x_i \beta} + \gamma_{i,m(t)} + \delta_{i,a(t)} + u_i + v_i + \sum_{l=1}^{L} bs(c_{lit}, n_k^{(l)}, 4) \end{aligned} \quad \text{(Supplementary Material 4.1)}$$

where $\lambda_{it}$ denotes the rate parameter of the Poisson distribution and $\pi$ is the probability of a zero count. Similar to our previous work[13], we included a population offset $\log(P_{it})$ such that $\log(\eta_{it})$ represented the DIR per 100,000 population. The DIR was then modelled with an intercept $\beta_0$, alongside $L = 4$ delayed climatic effects ($bs(c_{lit})$) with $n_k^{(l)}$ knots for the $k^{\text{th}}$ climatic variable, fixed effects, temporal random effects ($\gamma_{i,m(t)}$ and $\delta_{i,a(t)}$ for monthly and yearly effects respectively), and spatial random effects ($u_i$ and $v_i$). We will now describe these fixed and random effects in further detail.

##### *Supplementary Material 4.1. Delayed non-linear climatic effects*

We used distributed lag non-linear models (DLNMs) to capture the potentially non-linear and delayed effects of climatic variables on dengue incidence rates. As described in[13], DLNMs are a flexible modelling framework which are based on the statistical concept of a *crossbasis*; a bi-dimensional functional space comprised of a combination of two sets of basis functions which represent the relationships with the response in the dimensions of the predictor and lags respectively[57]. In particular, usage of the DLNM framework involves the simultaneous specification of i) non-linearity in the exposure-response relationship and ii) delayed effects in the lag-response dimension. The specification is achieved via the *cross-basis*, thus creating an exposure-lag-response function.

We used the R package `dlnms` to estimate our DLNMs. Here, similar to[13], we employed basis splines (B-splines) for our two sets of basis functions, which were combined to generate our cross-basis functions[57]. For each climatic variable, in the exposure-response dimension, we specified two equally spaced knots to allow for flexibility (and hence, complexity) in the relationship between the climatic variable and dengue incidence. The basis functions were centred on the mean average of the climatic variable. Then, we considered varying maximum time lags (of between two to four months) for the lag-response dimension for each climatic variable, and specified the knots at individual months to reflect the exploratory analyses.

##### *Supplementary Material 4.2. Temporal and spatial random Effects*

The current model's temporal and spatial random effects have been described elsewhere in our previous analysis[13], except here we replaced the department-specific temporal effects with corresponding province-specific (monthly and yearly) temporal effects and the department-level BYM2 model (an adapted Bayesian version of the Besag-York-Mollié (BYM) model) with a province-level BYM2 model[55]. Similar to previous analyses which implemented the BYM2 model, penalised complexity prior distributions were employed for precision parameters ($\tau$), which meant that $P(\frac{1}{\tau} > 0 \cdot 5) = 0 \cdot 01$[22,56].

Briefly, the monthly and yearly random effects aimed to capture seasonal (cyclic) patterns in a province's DIRs and year-to-year heterogeneities (such as time-varying reporting rates). The BYM2 model aimed to capture the spatial autocorrelation between the individual provinces, whilst additionally estimated any unstructured (i.e. non-spatial) heterogeneity in the provinces.

##### *Supplementary Material 4.3. Fixed Effects*

Our fixed effects, expressed as a matrix of four columns $\boldsymbol{x_i}$ in Equation Supplementary Material 4.1, comprised of; i) our seasonality indicator for the summer months (= 1 for months of December to April, and 0 otherwise) common to each department and ii) our momentum oscillator; Relative Strength Index (RSI), (which combined lagged RSI values of the past two months), iii) the proportion of a province's population declaring to be living in an urban area, and iv) the preceding month's year-to-year difference in DIR.

First, the objective of the simple binary seasonality variable was to capture seasonal trends of the summer months which are common across the departments. Whilst our climatic variables accounted for the influence of environmental conditions on vector abundance and incidence rates, the seasonality variable provided a means of accounting for possible population transience and increased outdoors activity during summer months.

Second, the proposed concept of the RSI variable was motivated by its usage as a technical indicator in financial markets to measure the strength and momentum of price movements [109]. For each department, we employed the past two values of the RSI, which were each calculated the smoothed moving average of the previous three month's DIRs. The formula used for RSI was:

$$\text{RSI}_t = 100 - -\frac{100}{(1 + \text{RS})}$$

where RS is the *relative strength*, defined as the smoothed ratio of average gains (sum of the upward DIR movements over the past three months, divided by three – the number of months) over the average losses (sum of the downward DIR movements over the past three months, divided by three). By using the product of the past two RSI values ($\text{RSI}_t \times \text{RSI}_{t-1}$), we tracked i) the recent momentum in large DIR movements and ii) the overall magnitude (or recent levels) of DIR (via the product). So, even if we have recently observed downward movements in DIR, two strong consecutive momentum values would indicate that the DIR has recently been (or is currently) at high levels.

Third, the continuous urban variable was included to estimate the possible amplifying effect of a highly urbanised population on DIR, possibly arising due to greater human-to-vector contact and greater opportunities for larval habitat creation. This was a fixed variable over calendar time, indicating the percentage of the population living in urban (as opposed to rural) area.

Finally, we included the preceding month's year-to-year difference in DIR as a proxy for possible outbreak years where larger proportions of the population may have entered the susceptible sub-population. We modelled the effect using a natural spline with four knots to allow for sufficient flexibility for the likely non-linearity in both the negative and positive effects in non-outbreak and outbreak years respectively. For example, if the preceding month's DIR was far (slightly) greater than the preceding year, we would expect the current month's DIR would be far (slightly) greater than the previous year.

#### *Supplementary Material 4.4. Covariates*

Table SI 4: **Covariates used in our Bayesian climate-based model**. All covariates were determined by exploratory analyses and performance of the Bayesian climate-based model in the model development period of 2010-2017.

| **Covariate** | **Description** |
|---|---|
| Monthly random effect | Cyclic random walk (of order 1) prior distribution used for monthly random effects for each province. |
| Yearly random effect | Exchangeable Normal(0,1) prior distribution used for yearly random effects for each province. |
| BYM2 spatial effects | Structured spatial and unstructured components to capture spatial correlation and individual province uniqueness respectively in an adapted Bayesian version of the Besag-York-Mollié (BYM) model with penalised complexity prior distributions[55,56]. |
| Minimum temperature (*tmin*) | Two-month rolling average (right-aligned) of the monthly average of daily minimum temperature, specified using a DLNM (Distributed Lag Non-linear Model). The DLNM featured B-splines with two equally-spaced knots in the exposure-response dimension and knots at one and two months in the lag-response dimension (which included a maximum lag of three months). |
| Precipitation (*prec*) | Two-month rolling average (right-aligned) of the monthly total precipitation (mm), specified using a DLNM. The DLNM featured B-splines with two equally-spaced knots in the exposure-response dimension and knots at one and two months in the lag-response dimension (which included a maximum lag of three months). |
| Standardized Precipitation Index (SPI-6) | Drought indicator (SPI), measuring precipitation anomalies over a six month period[43], specified using a DLNM. The DLNM featured B-splines with two equally-spaced knots in the exposure-response dimension and knots at one and two months in the lag-response dimension (which included a maximum lag of two months). |
| El Niño Coastal Index (ICEN) | The ICEN, a sea-surface temperature indicator specific to Peru[44,45], specified using a DLNM. The DLNM featured B-splines with two equally-spaced knots in the exposure-response dimension and knots at one and two months in the lag-response dimension (which included a maximum lag of four months). |
| Seasonality | A binary summer-winter seasonality variable to account for the non-climatic effects of possible population transience and/or general DIR patterns (across the provinces) in the absence of other amplifying effects (such as climatic influences). |
| Urban population | The percentage of a population living in an urban (as opposed to rural) area, as reported by the 2017 national census. Note that we assumed a constant (as opposed to time-varying) value (from 2017) to allow us to estimate the overall effect of a highly urbanised province. |
| Lagged monthly difference with previous year | One-month lagged value of the difference between that month's DIR and the DIR from the corresponding month in the previous year. The effect was estimated using a natural spline with four knots to allow for the likely non-linear effects of negative and positive differences, and the variable was included in an attempt to capture outbreak years (as opposed to non-outbreak years). |
| Relative Strength Index (RSI) | The product of monthly momentum indicator, RSI (calculated over a horizon of three months), lagged by one month and two months, which aimed to capture momentum movements. Whilst it may seem counter-intuitive to also square negative momentum, in our exploratory analysis, we found stronger correlation between DIR and the squared lag of RSI, which may be explained by large negative movements subsequent preceding higher DIR (possibly due to reporting artefacts or seasonal patterns). |

#### Supplementary Material 5. Ensemble framework: Additional details

##### *Supplementary Material 5.1. Background to probabilistic ensemble forecasting*

A probabilistic ensemble forecasting model is a statistical machine learning approach which combines information from several forecasting models and assigns probabilities to individual events or targets. An ensemble approach is increasingly used in infectious disease forecasting due to frequent outperformance of forecasts from individual models, whilst often providing decision-makers with more consistent, stable information across both time and space[59,110]. In the context of forecasting dengue incidence, a study (in the cities of Iquitos, Peru and San Juan, Puerto Rico) found that ensemble approaches produced superior forecasting skill (as measured by proper scoring rules) over an eight-year period[61].

##### *Supplementary Material 5.2. Trained and untrained ensemble frameworks*

For quantile forecasts, we use the following equation to weight the components of each ensemble framework:

$$q_r^E = \sum_{k=1}^{K} w_k q_r^k \qquad \text{(Supplementary Material 5.1)}$$

where $w_k$ is defined as before and $q_r^k$ is the $r^{\text{th}}$ quantile value estimated by the $k^{\text{th}}$ model component. We produced forecasts for 23 quantile levels (identical to those used in the COVID-19 Forecast Hub in the United States[64]). For trained weighting of the ensemble ($E$), weights $w_k$ are determined such that they minimised the sum of quantile losses over the past 12 months. These weights were determined for all provinces jointly (i.e. province-independent model weights) and for each province independently (i.e. province-dependent weights). For untrained weighting of the ensemble, the weighted sum (in Equation Supplementary Material 5.1) is replaced with the median or mean average function applied again to individual modelling components' predictive quantiles. To evaluate probabilistic sharpness and bias, we used the weighted interval score (WIS), a proper scoring rule which is a discrete approximation to the CRPS[63,71]. We assessed WIS on a logarithmic scale to follow recommended procedures, and lower scores indicate better performance.

#### Supplementary Material 6. Ensemble component forecasters: Additional details

As described in the main text, we developed Seasonal Auto-Regressive Integrated Moving Average (SARIMA) models. These statistical time series models were fitted independently to the logarithm of monthly cases for each province independently using the Auto-ARIMA implementation of the Statsforecasts package and Darts library in Python[74,75]. This implementation selects the order of the ARIMA model based on the AIC[52], and quantile forecasts are generated by assuming normality in forecast errors. We refitted the model at each individual time point to reflect real-world conditions as new information becomes available. Here, we highlight that the updating of our SARIMA model, including the order (i.e. its parameters), at each time step is an important task. This updating differs to past forecasting studies for dengue (e.g.[61,65]) where the order of the ARIMA/SARIMA models were fixed across the testing set. We allow for this updating to i) ensure equitable comparison and ii) to allow the most recent data determine the order of the model that we should use. Indeed, for deep learning models (see below), one would always allow its weights (and potentially other hyperparameters) to be updated at each time point, and so we perform a similar procedure for the SARIMA model.

The first deep learning model that we implemented was TimeGPT, a transformer-based model with self-attention that has been pre-trained on over 100 billion data points from different domains. Briefly, the model uses an encoder-decoder structure with multiple layers, each consisting of residual connections (which allow bypassing certain layers) and layer normalisation (which helps to stabilise the model)[77,78]. We implemented this model for the logarithm of cases with and without climatic covariates (two-month rolling averages of precipitation and minimum temperature)[77,78]. While the TimeGPT model allows for zero-shot inference, we used a fine-tuning process which refines the model's parameters to suit the task of forecasting dengue cases. This fine-tuning process consists of ten training iterations of model fitting to the training data to minimise forecasting error. Then, the updated TimeGPT model (after fine-tuning) readily produces quantile forecasts by using conformal inference — a distribution-free way to produce statistically rigorous prediction intervals[79].

Our second deep learning model was a temporal convolutional network (TCN) from the Darts library in Python[74]. Again, the TCN architecture consists of an encoder-decoder structure. The encoder has stacked dilated causal convolutional neural networks which aim to describe longer-term temporal dependencies in the data. The decoder uses a residual block and an output dense layer. For each observation, we used a lookback period (or input chunk length) of 48 months which means that we allowed for the past 48 months of data to be considered by the TCN model when making predictions. To avoid making potentially invalid distributional assumptions, we used quantile regression to directly estimate the quantiles of the predictive distribution. We again used climatic covariates of two-month rolling averages of precipitation and minimum temperature. These covariates were based on exploratory analyses from the model development period.

#### Supplementary Material 7. Outbreak detection: Additional details

##### *Supplementary Material 7.1. Classification of DIR exceeding thresholds*

Our modelling frameworks (see Materials and methods) each produced predictive distributions which we represented using predictive quantiles.

First, using the samples from the predictive distribution, we derived probabilities of DIR surpassing thresholds of 50 (and 150) per 100,000. Rather than using an arbitrary cut-off probability for classifying whether an outbreak would occur, we devised an optimisation scheme which maximised the historical performance of the forecasting model. Hence, when classifying whether DIR would surpass a specific threshold for each given month, a cut-off probability was calibrated using past (i.e. training) data that maximised the historical AUC, the area under the Receiver Operating Characteristic (ROC) curve. The procedure ensured that we used the cut-off probability that maximised the difference between the true positive rate and false alarm rate. The aim was to ensure that the classification would provide public health authorities with a more reliable forecasts of whether DIR would surpass the threshold. To reflect a real-world scenario, the cut-off probabilities were iteratively updated as new data became available.

#### Supplementary Material 8. Additional discussion of limitations

Another key limitation across the study is the quality of data available. For dengue surveillance data, we did not have access to data on public health interventions (e.g. concurrent vector control programmes) or reporting rates (e.g. number of participating healthcare centres and hospitals) which can help to gauge underreporting. However, years with greater DIRs (relative to historical averages) were often associated with lower average proportions of cases being confirmed (hence, more being classified as probable cases), which may indicate time-varying pressures on the healthcare centres and surveillance system, and provides further motivation for our modelling of year-to-year heterogeneities in our climate-based model (Figure SI 42). The surveillance data also did not provide stratification by serotype, and the associated unavailability of seroprevalence data restricted us from accounting for potentially time-and-space-varying serotype dominance and immunity profiles of populations. Similarly, the lack of demographic data (such as age of case) for probable and confirmed cases limited us from assessing how mean age of infection influences the epidemic dynamics (such as the force of infection, as performed by[24]) or how the demographics relate to our computed wavelet-based measures (such as synchrony, coherence, or the period of multiannual cycles). Thus, consistent with the WHO's cited lack of data on circulating serotypes[91], we advocate for future collation of age-structured dengue cases, alongside enhanced genomic and serological surveillance across at-risk areas, all of which would provide insights about immunology and virology issues (such as multitypic immunity, genomic diversity, and repeat infections) across space and time.

### Supplementary Material 9. Figures and tables

#### *Supplementary Material 9.1. Wavelet and climate-based analyses*

##### Supplementary Material 9.1.1. Exploratory analyses

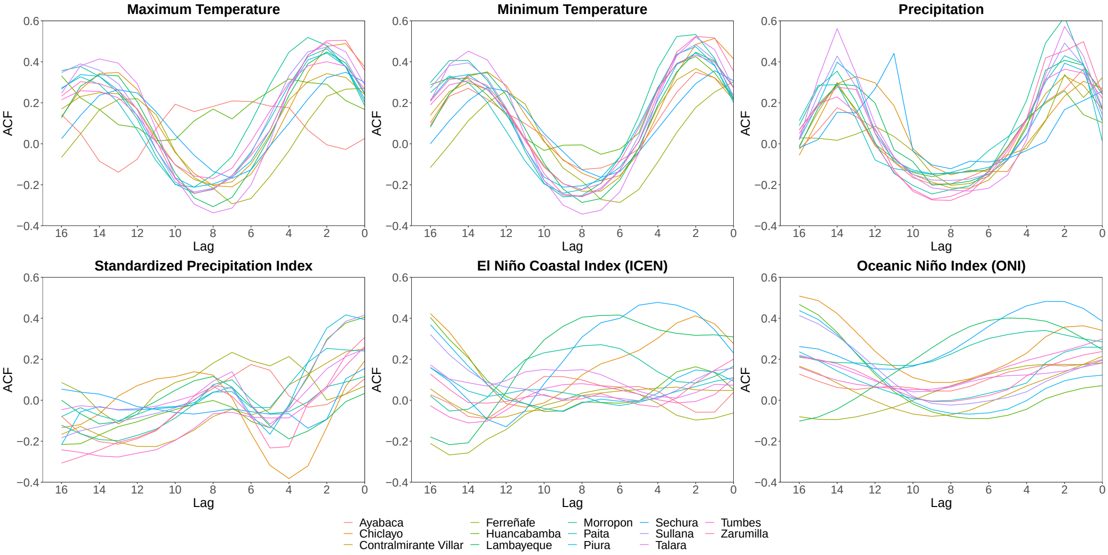


Figure SI 1: **Relationships between lagged climatic variables and dengue incidence rates.** Across the model development period (2010 to 2017), the cross-correlation plots depict the province-level relationship between six individual climatic variables and the monthly dengue incidence rate (DIR) per 100,000. ACF (y-axis) denotes the auto-correlation function, and the lag (x-axis) denotes the lag (in months) from the climatic exposure to subsequent monthly DIR.

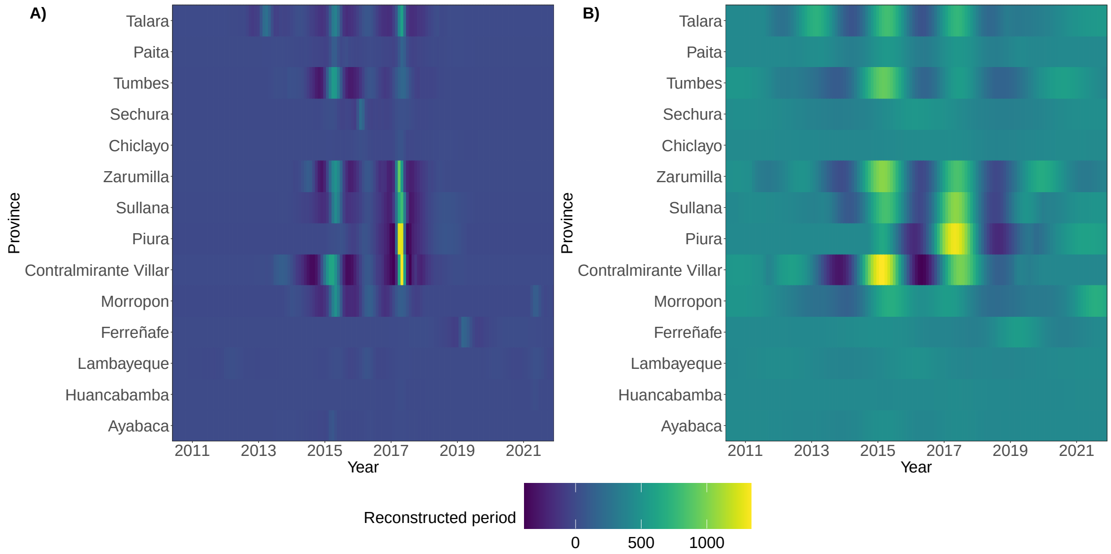


Figure SI 2: **Wavelet reconstructions of cycles of Dengue Incidence Rate (DIR) time series.** Left: Reconstructed annual cycles of province-level DIRs, where provinces are sorted in ascending order by percentage of population living in urban areas. Right: Reconstructed multiannual cycles of province-level DIRs, where provinces are again sorted in ascending order by percentage of population living in urban areas. Higher values of the annual (multiannual) period indicate peaks in DIR across the annual (multiannual) reconstructions, where the wavelet-based reconstructions are the reconstructions of province-level DIR time series using the initial decompositions into time-frequency and then, frequencies corresponding to annual (multiannual) cycles.

#### Supplementary Material 9.1.2. Spatiotemporal incidence trends and properties of reconstructed cycles

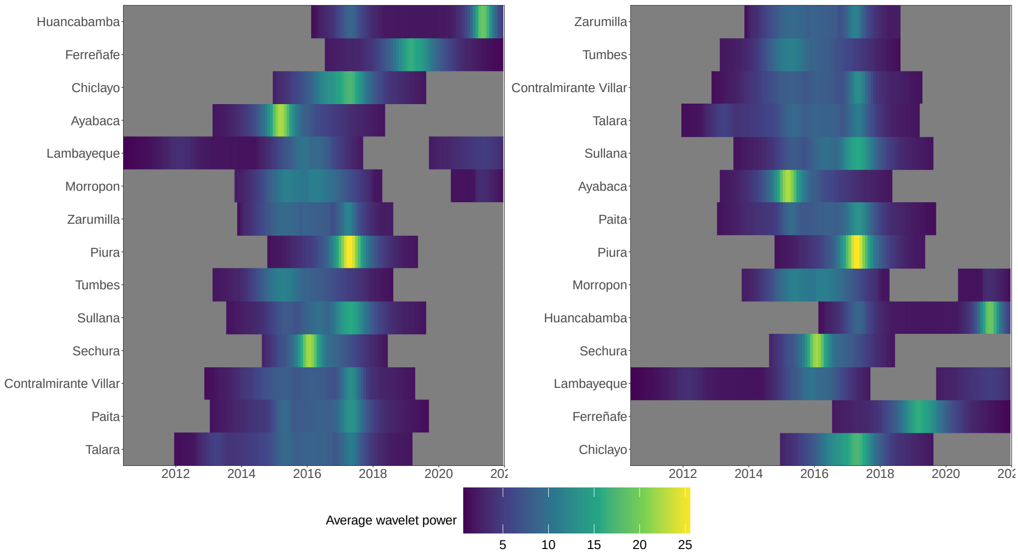


Figure SI 3: **Average wavelet power of dengue incidence.** Cells depict average wavelet power of annual cycles per time step in provinces **(A)** sorted by longitude and **(B)** sorted by latitude, where non-filled (grey) cells depict non-significant wavelet power.

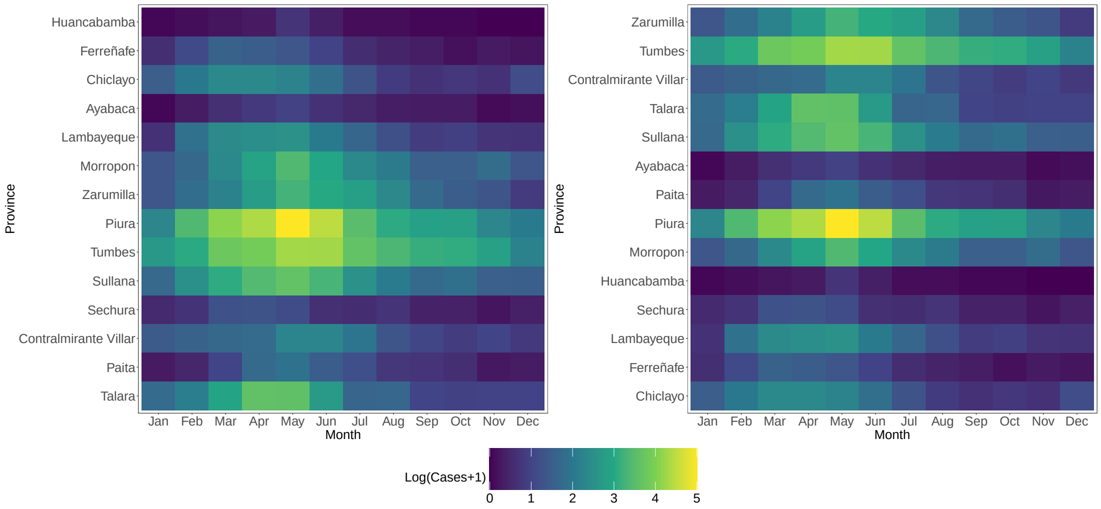


Figure SI 4: **Average spatiotemporal trends in dengue epidemics.** Monthly mean average of provinces' log(Cases + 1) where provinces are sorted **(A)** by longitude (from west to east) and **(B)** by latitude (from south to north). We visualise the logarithm of cases to enable comparison of epidemics of different sizes across different geographies. This is aligned with our proper scoring in our forecasting, as recommended by recent guidelines [72].

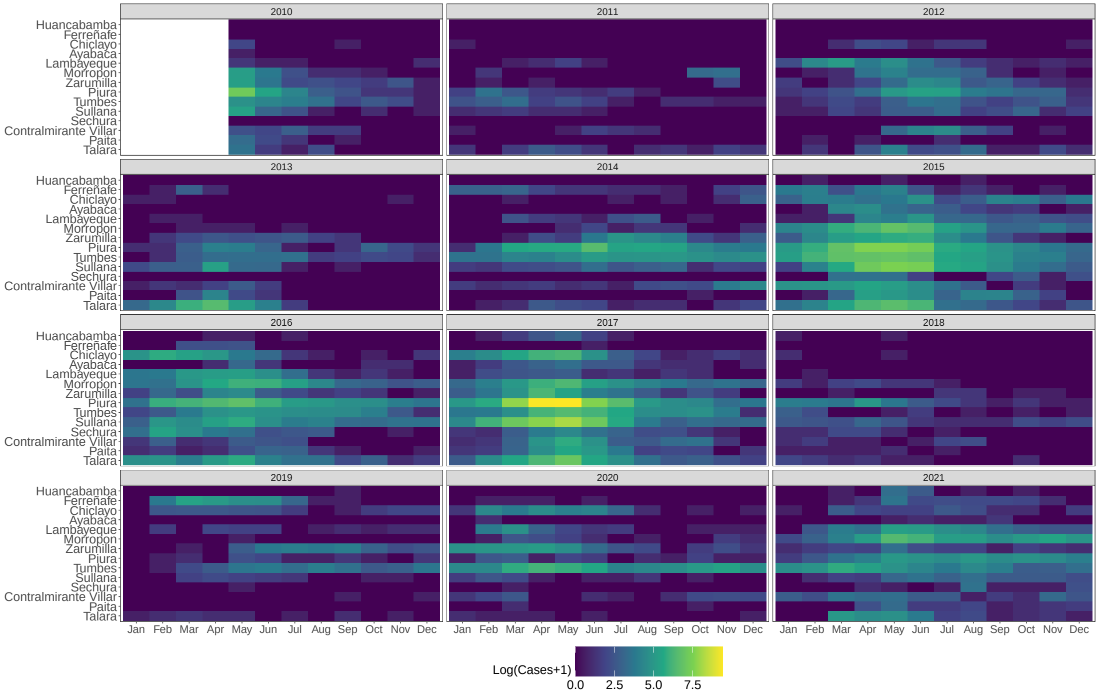


Figure SI 5: **Longitudinal spatiotemporal trends in dengue epidemics.** Provinces' log(Cases + 1) are shown, where provinces are sorted by longitude (from west to east). We visualise the logarithm of cases to enable comparison of epidemics of different sizes across different geographies. This is aligned with our proper scoring in our forecasting, as recommended by recent guidelines[72].

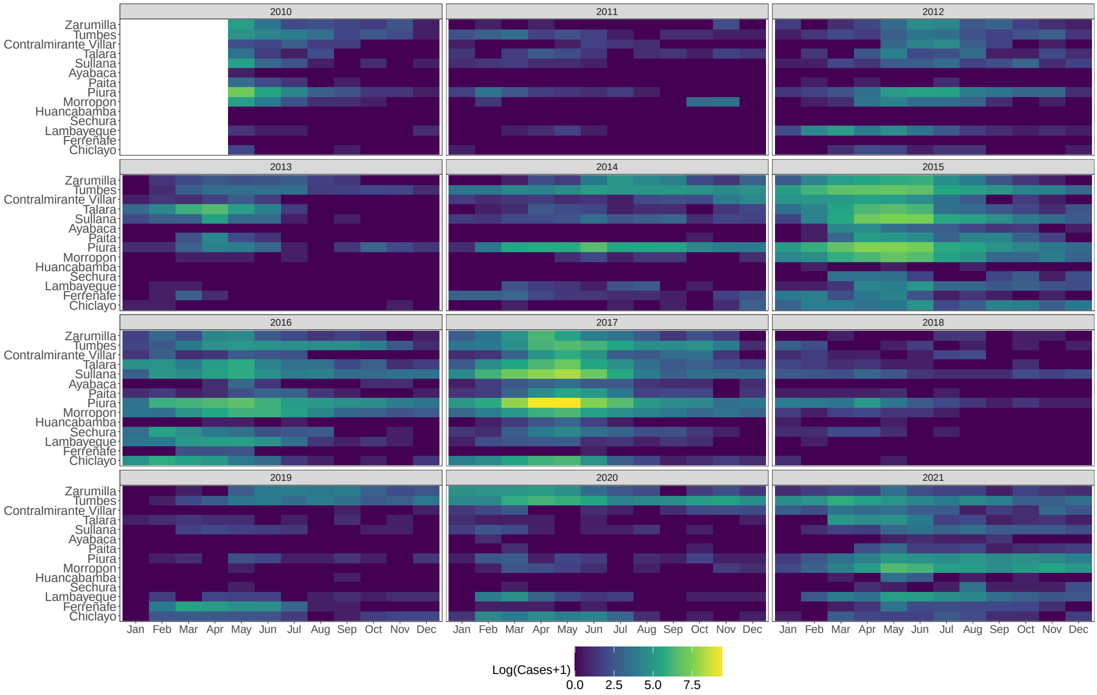


Figure SI 6: **Latitudinal spatiotemporal trends in dengue epidemics.** Provinces' log(Cases + 1) are shown, where provinces are sorted by latitude (from south to north). We visualise the logarithm of cases to enable comparison of epidemics of different sizes across different geographies. This is aligned with our proper scoring in our forecasting, as recommended by recent guidelines [72].

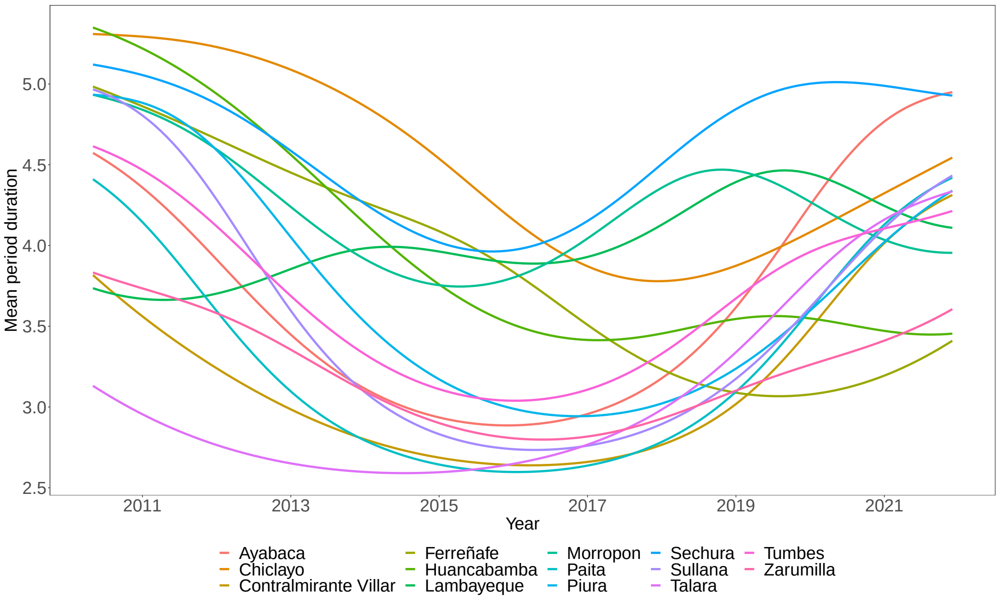


Figure SI 7: **Mean duration of multiannual dengue cycles.** Provinces' mean duration (in years) of wavelet-based reconstructed multiannual cycles of dengue incidence rate (DIR) per 100,000 across the studied period of 2010–2021.

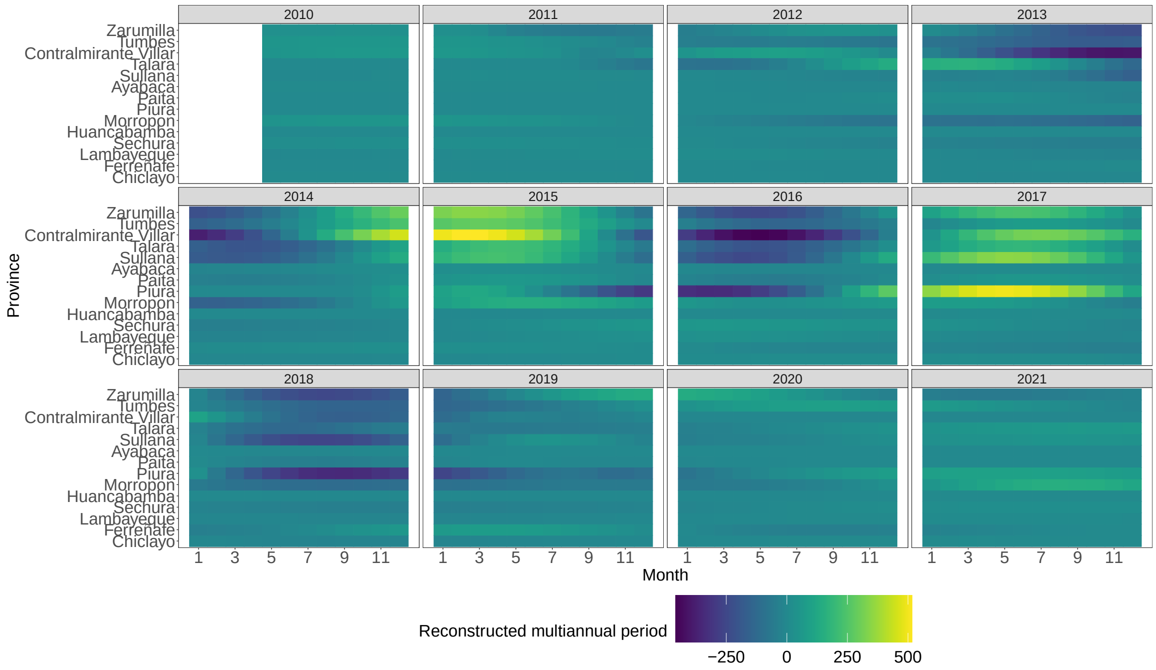


Figure SI 8: **Wavelet reconstructions of multiannual dengue cycles in higher temporal resolution.** Multiannual cycles of province-level dengue incidence are sub-divided into individual years, where provinces are sorted by latitude in ascending order (from south to north).

#### Supplementary Material 9.1.3. Phase coherence and epidemic synchrony

Table SI 5: **Summary statistics of provinces' phase coherence.** For both reconstructed annual and multiannual dengue cycles, for each (focal) province at each individual time point, we calculated the proportion of provinces with whom the focal province shared a statistically significant coherence. Here, we present the median, lower quartile (Q1), and upper quartile (Q3) of these proportions as a summary of the province-level coherence between the cycles across the study period.

| Time Series | Median | Q1 | Q3 |
|---|---|---|---|
| Reconstructed Annual Cycle | 0.62 | 0.46 | 0.77 |
| Reconstructed Multiannual Cycle | 0.46 | 0.31 | 0.62 |

Table SI 6: **Relationship between urbanised provinces and coherence.** For both annual and multiannual dengue cycles, the correlation between the proportion of a province's cycles with statistically significant coherence and the province's percentage of population living in urban areas.

| Time Series | Correlation (95% CI) |
|---|---|
| Reconstructed Annual Cycle | 0.58 (0.07, 0.85) |
| Reconstructed Multiannual Cycle | 0.46 (-0.09, 0.80) |

Table SI 7: **Summary statistics of provinces' epidemic synchrony.** For raw reported dengue incidence, reconstructed annual cycles, and reconstructed multiannual cycle, we calculated each province's median correlation ($r$) with the corresponding time series of the other 13 provinces. Here, we report the median, lower quartile (Q1), and upper quartile (Q3) of these correlations as a summary of the average province-level correlations between the time series.

| **Time Series** | **Median** $r$ | **Q1** | **Q3** |
|---|---|---|---|
| Raw Dengue Incidence Rates | 0.46 | 0.34 | 0.54 |
| Reconstructed Annual Cycle | 0.76 | 0.69 | 0.80 |
| Reconstructed Multiannual Cycle | 0.60 | 0.28 | 0.66 |

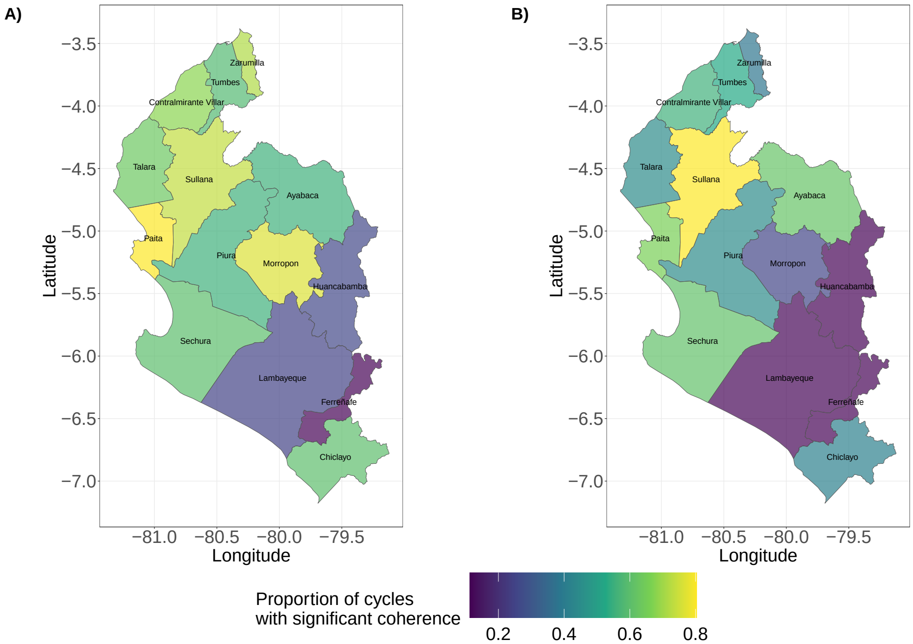


Figure SI 9: **Spatial distribution of coherence.** Maps depict the proportion of each provinces' reconstructed **A)** annual and **B)** multiannual cycles of Dengue Incidence Rates (DIRs) which share statistically significant coherence with other provinces.

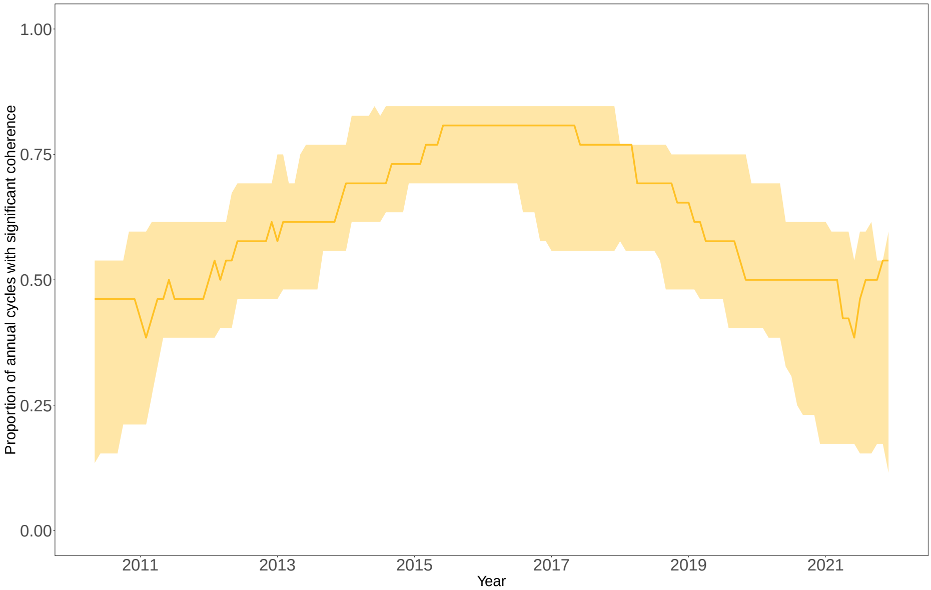


Figure SI 10: **Temporal distribution of annual phase coherence.** For each province (the so-called focal province) at each time point, we calculated the proportion of provinces with whom the focal province shared a statistically significant coherence. Here, we summarise the distribution of these proportions over time by depicting the median proportion at each time point (gold line) and the lower and upper quartiles of proportions.

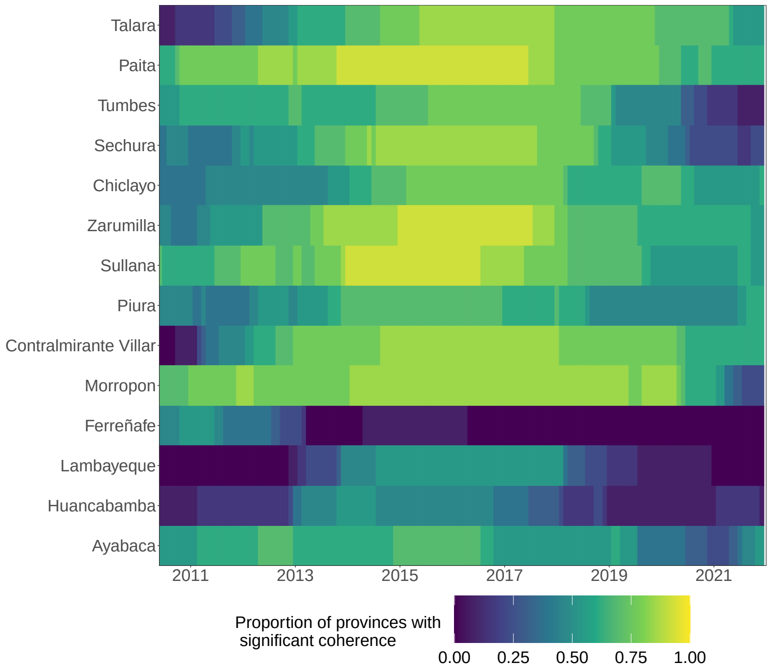


Figure SI 11: **Coherence by level of urbanisation.** Proportion of a focal province's annual dengue cycles which share statistically significant coherence with the cycles of other provinces at each time point. Provinces are sorted in ascending order by proportion of the population that lives in an urban area.

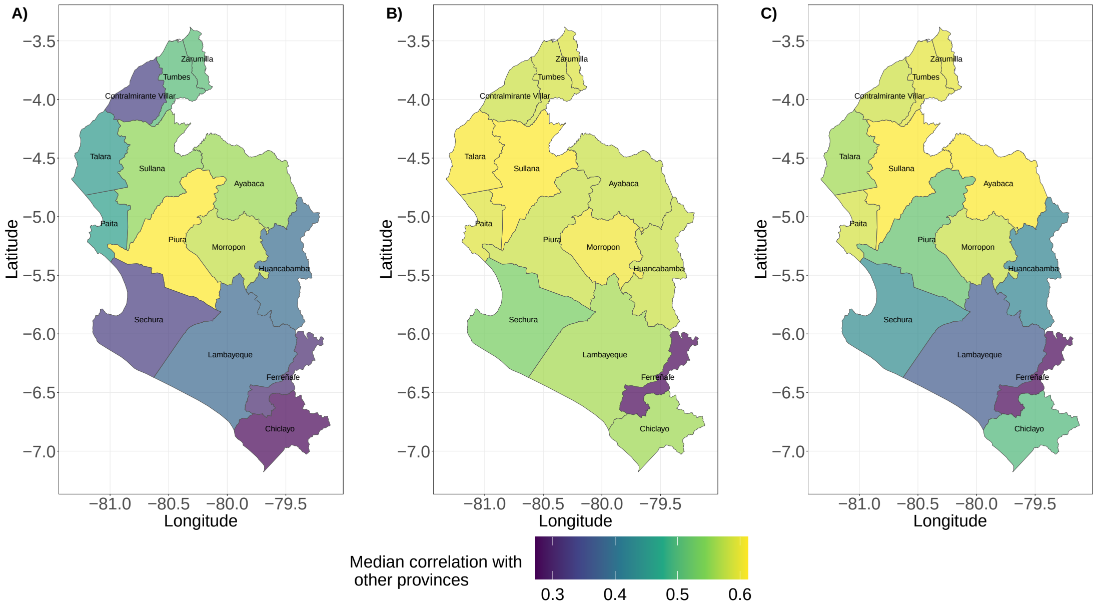


Figure SI 12: **Spatial distribution of epidemic synchrony.** Maps depict the median correlation between a focal province's **A)** raw Dengue Incidence Rate (DIR) time series, **B)** reconstructed annual cycle or **C)** multiannual cycle, and the corresponding time series of all other provinces.

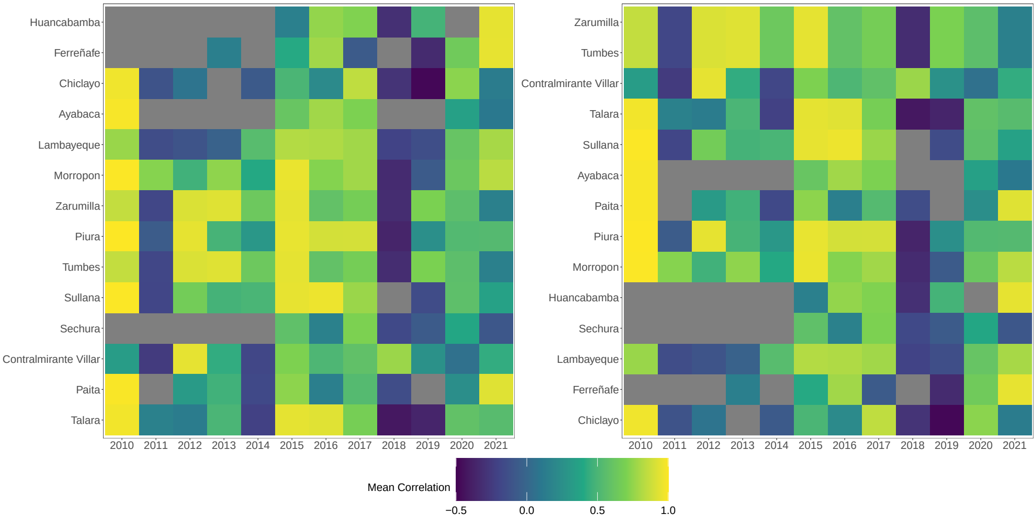


Figure SI 13: **Epidemic synchrony across time and space.** Visualisation of the mean average correlation between a province's raw monthly Dengue Incidence Rate (DIR) time series and the other provinces. Provinces are sorted in ascending order **(A)** by longitude (from west to east) and **B** by latitude (from south to north).

#### Supplementary Material 9.1.4. Climatic, geographic, and human influences on epidemic dynamics

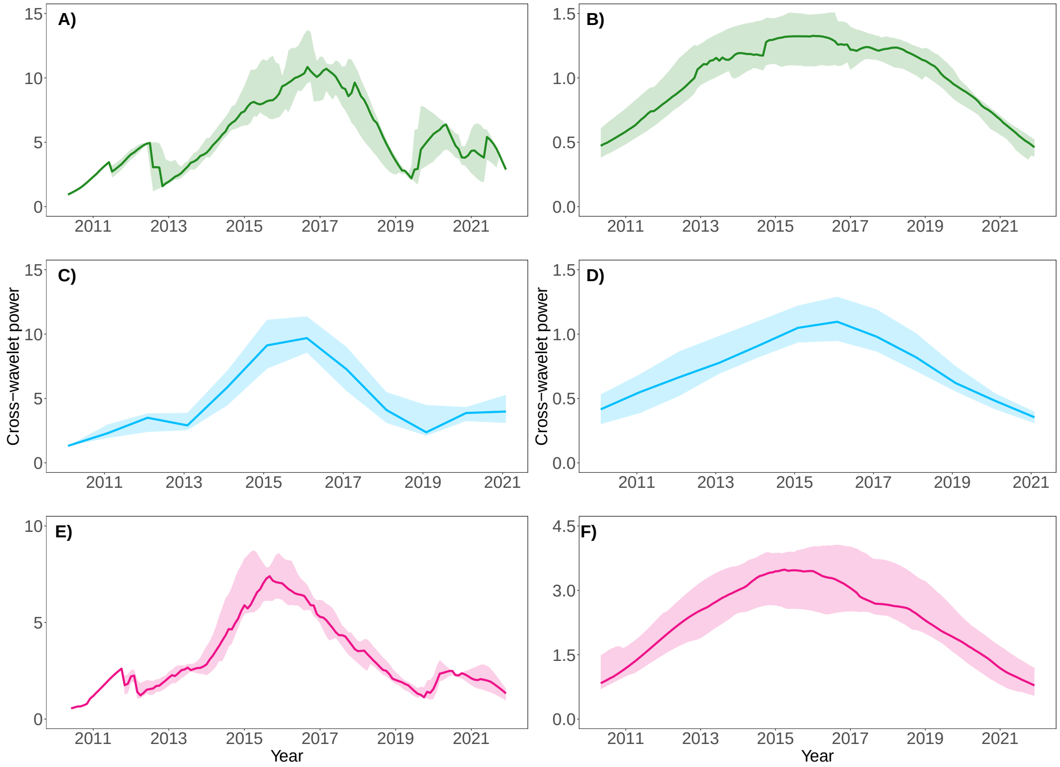


Figure SI 14: **Distributions of cross-wavelet power between climatic conditions and reconstructed dengue cycles.** Panels summarise the distribution of cross-wavelet power between either reconstructed annual (left column) or multiannual dengue cycles (right column) and climatic conditions. **(A)-(B)**: Cross-wavelet power between reconstructed annual/multiannual cycles and monthly average maximum temperature. **(C)-(D)**: Cross-wavelet power between reconstructed annual/multiannual cycles and monthly total precipitation. **(E)-(F)**: Cross-wavelet power between reconstructed annual/multiannual cycles and monthly El Niño Coastal Index. Solid lines depict the median cross-wavelet power at each time, whilst the shaded regions depict the interquartile range – IQR; ranging from the first/lowest quartile (Q1) to the third/upper quartile (Q3).

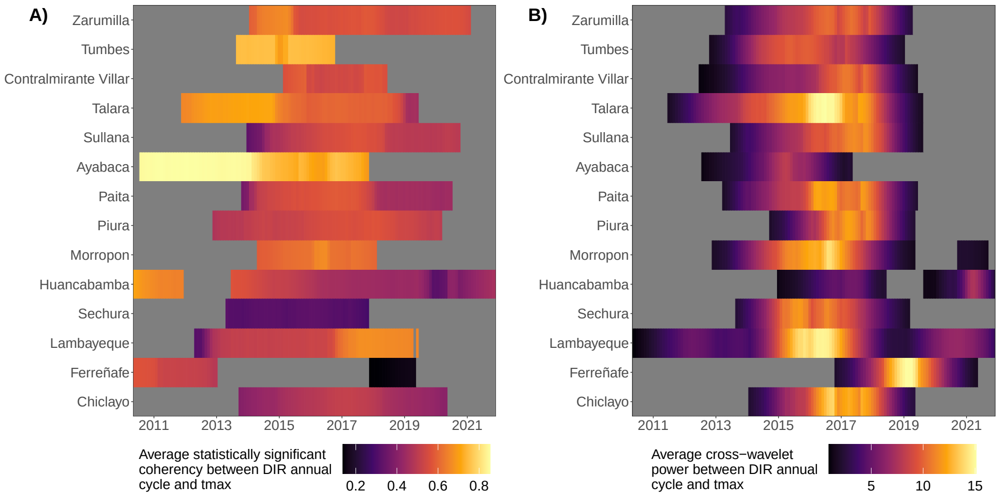


Figure SI 15: **Maximum temperature coherence and cross-wavelet power (annual cycles).** (**A**): Provinces' average statistically significant coherence between reconstructed annual dengue cycles and monthly average daily maximum temperature (denoted tmax above). (**B**): Provinces' average cross-wavelet power between reconstructed annual dengue cycles and monthly average daily maximum temperature. Provinces are sorted in ascending order by latitude (from south to north), and non-filled (grey) cells depict non-significant wavelet coherency/cross-wavelet power.

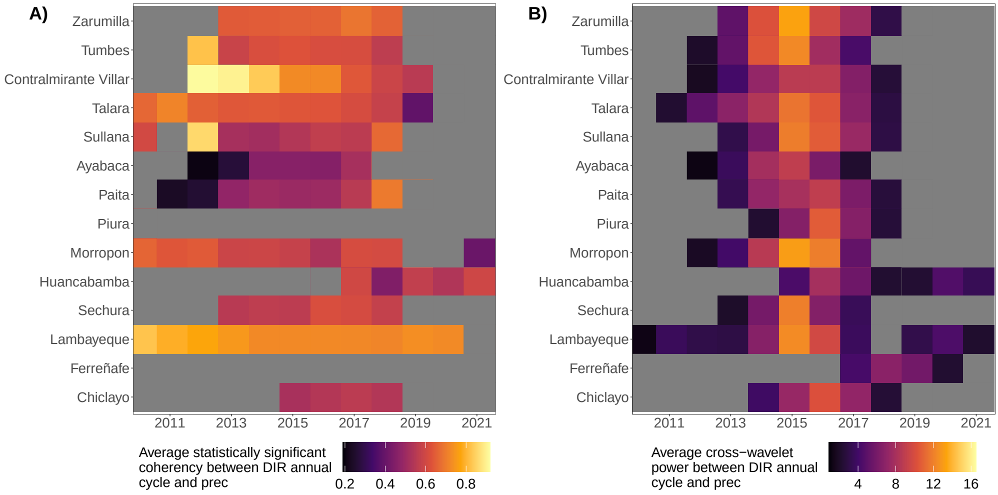


Figure SI 16: **Monthly precipitation coherence and cross-wavelet power (annual cycles).** (**A**): Provinces' average statistically significant coherence between reconstructed annual dengue cycles and monthly total precipitation (denoted prec above). (**B**): Provinces' average cross-wavelet power between reconstructed annual dengue cycles and monthly total precipitation. Provinces are sorted in ascending order by latitude (from south to north), and non-filled (grey) cells depict non-significant wavelet coherency/cross-wavelet power.

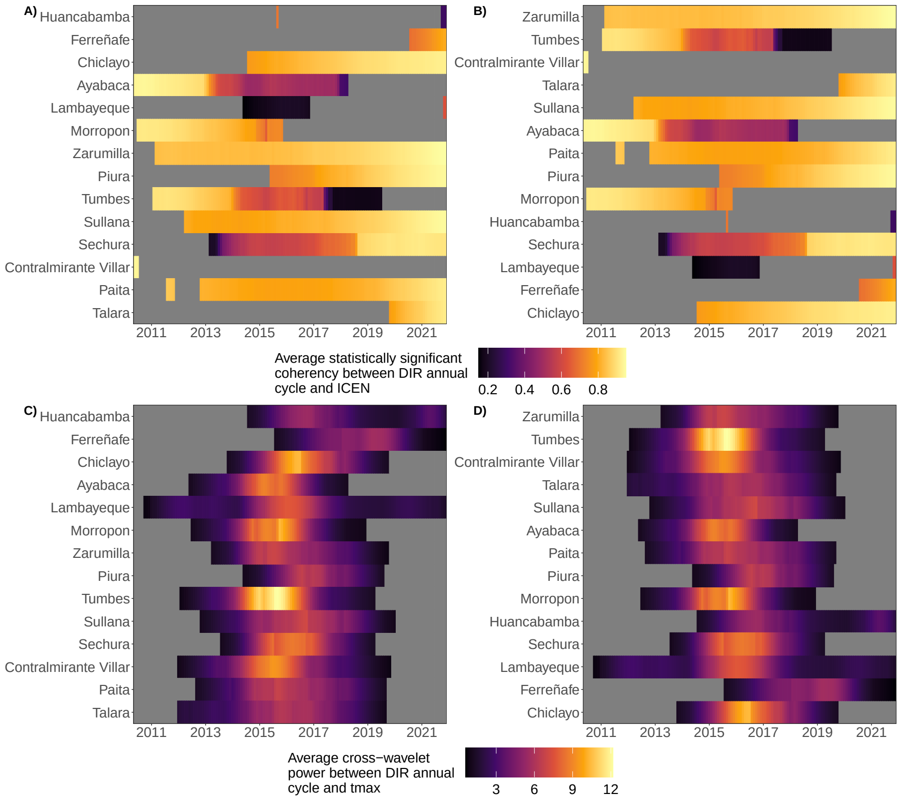


Figure SI 17: **El Niño Coastal Index (ICEN) coherence and cross-wavelet power (annual and multiannual cycles).** **(A)** and **B**: Provinces' average statistically significant coherence between reconstructed annual dengue cycles and monthly El Niño Coastal Index (denoted ICEN above), where provinces are arranged in ascending order by longitude (from west to east) and latitude (from south to north) respectively. **(C)** and **D**: Provinces' average cross-wavelet power between reconstructed multiannual dengue cycles and monthly El Niño Coastal Index, where provinces are again arranged in ascending order by longitude and latitude respectively. Non-filled (grey) cells depict non-significant wavelet coherency/cross-wavelet power.

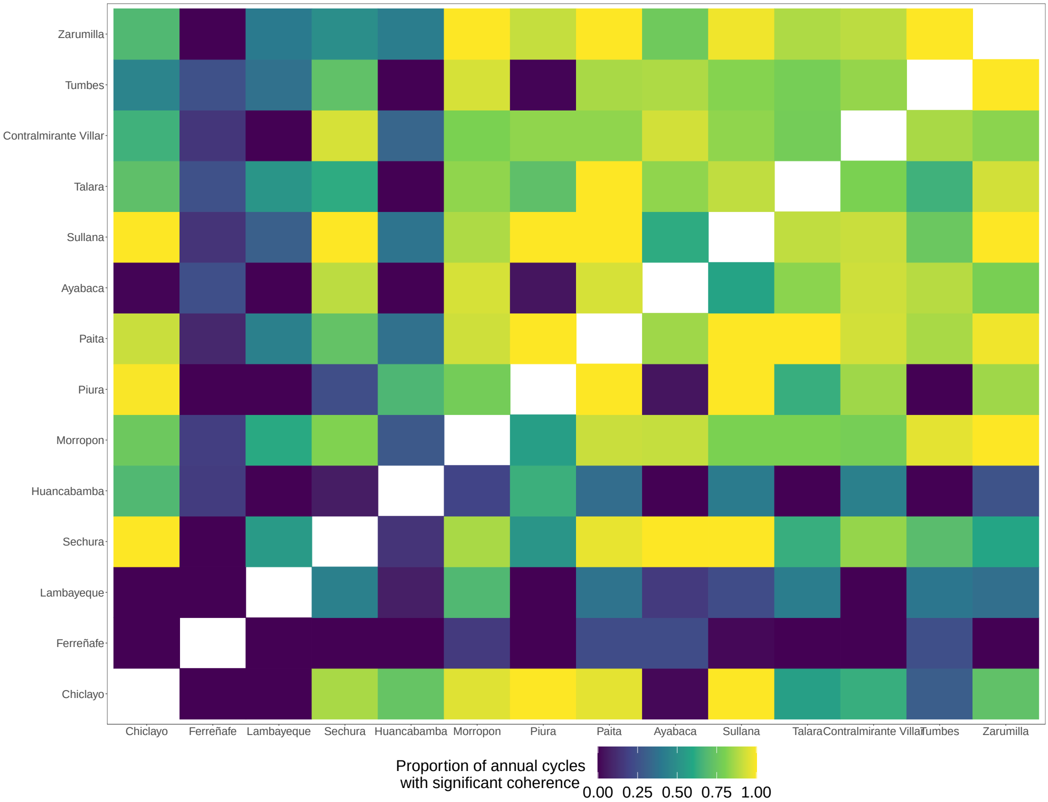


Figure SI 18: **Pairwise coherence visualisation.** Cells depict proportion of annual cycles between provinces that are statistically significant, where provinces are sorted by latitude (from south to north). White filled cells denote the diagonal entries of a focal province's coherence with its own time series. The visualisation captures the strength of relationships between a province and other provinces, ranging from close neighbours to provinces further away.

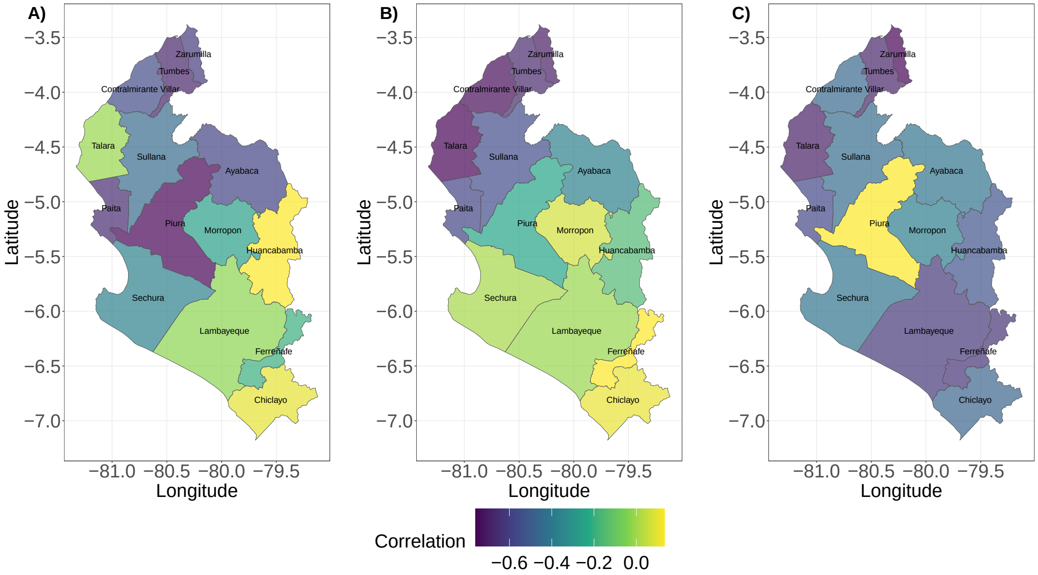


Figure SI 19: **Relationship between provinces' pairwise distance and epidemic dynamics.** (**A**): Correlation between provinces' pairwise distances and the corresponding median pairwise Pearson correlation of Dengue Incidence Rate (DIR) time series. (**B**): Correlation between provinces' pairwise distances and the proportion of the reconstructed annual dengue cycles sharing significant coherence. (**C**): Correlation between provinces' pairwise distance and the proportion of the reconstructed multiannual dengue cycles sharing significant coherence.

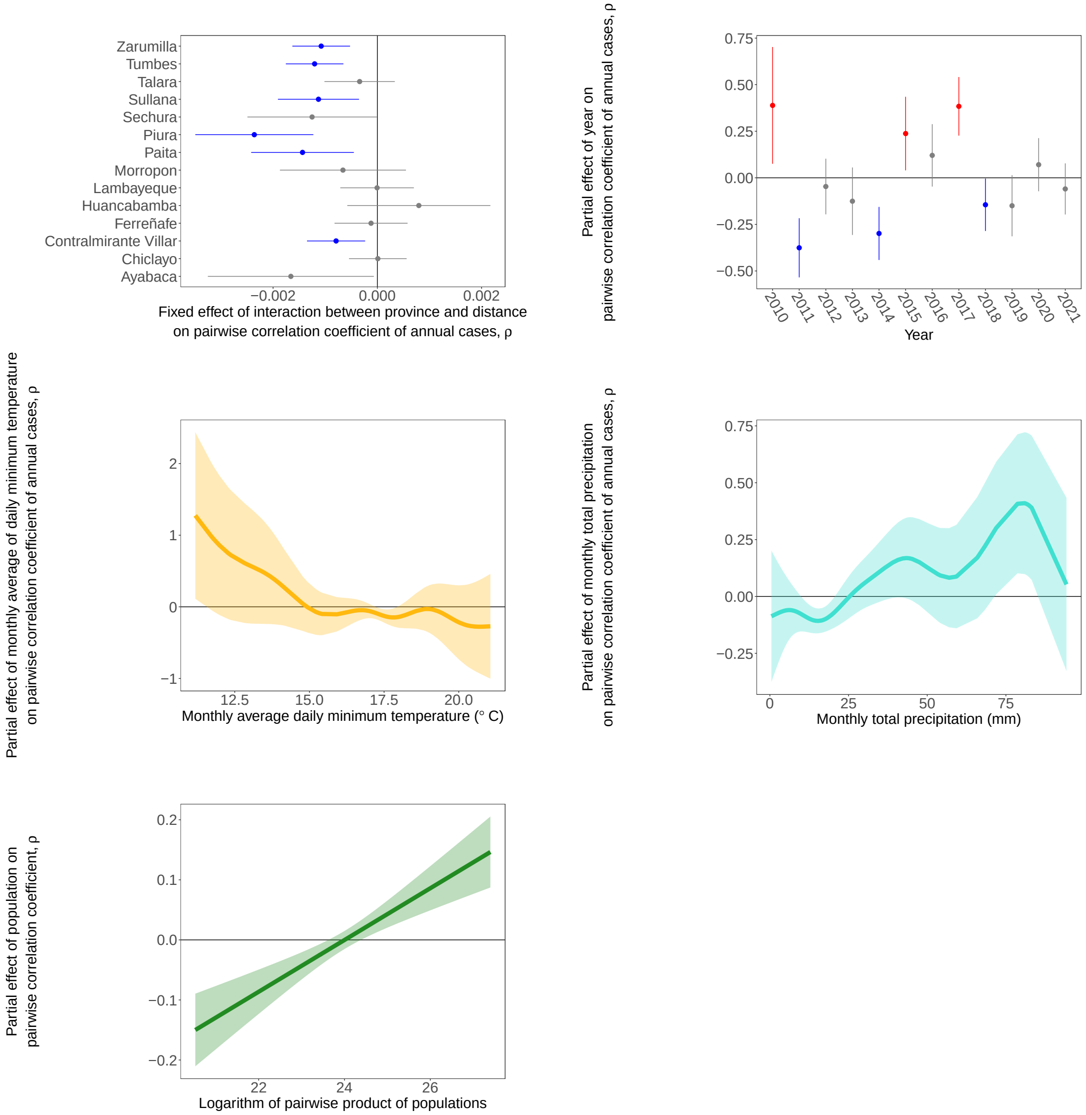


Figure SI 20: **Results from best-fitting model of epidemic synchrony.** Top panel: Visualisation of fixed effect of interaction between and distance and partial effect of year on the pairwise annual correlation coefficient (between annual case cycles), where blue and red denote negative and positive statistically significant estimated effects respectively. Middle panel: Visualisation of the partial effects of minimum temperature (left) and precipitation (right) on the pairwise annual correlation coefficient, where the x-axes denotes the yearly average of the corresponding monthly averages. Bottom panel: Visualisation of the partial effect of the pairwise product of logarithm of populations on the pairwise annual correlation coefficient. Shaded regions depict the 95% confidence intervals in each case.

#### *Supplementary Material 9.2. Climate-based modelling*

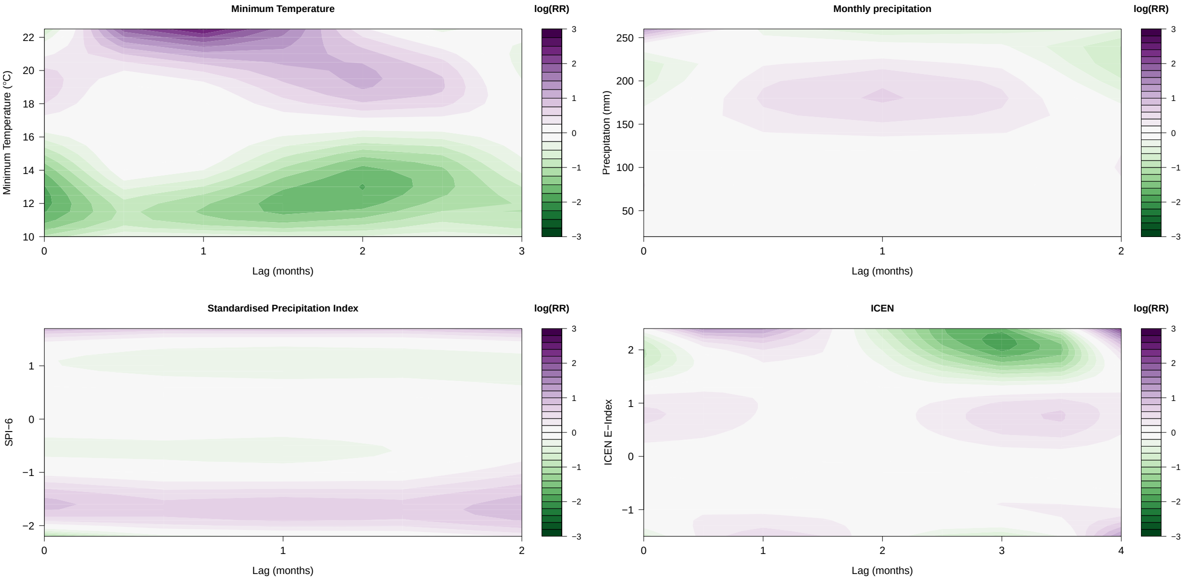


Figure SI 21: **Model-based exposure-lag-response relationships between climatic variables and dengue incidence.** Across 2010 to 2021, the relative risk (RR) of dengue incidence, on a logarithm scale, for each of our climatic DLNMs (Distributed Lag Non-linear Models) in our climate-based Bayesian spatiotemporal model. RR was defined relative to the risk induced by the mean observed value of each climate variable, and thus, log RR values above 0 (pink to purple) correspond to elevated RR of dengue incidence, whilst values below 0 (green) correspond to reduced RR. The DLNMs were included in a model alongside various temporal, spatial, and spatiotemporal effects, as well as fixed effects.

##### *Supplementary Material 9.3. Probabilistic ensemble forecasting*

The following section presents results and tables for probabilistic forecasting of dengue cases across i) the model development period (2010 to 2017 inclusive) and ii) the testing period (2018 to 2021 inclusive). All probabilistic forecasts were made with a forecast horizon of one month and excluded all future data from model training.

##### Supplementary Material 9.3.1. Model development period – Summary

Table SI 8: **Results for forecasting log(Cases + 1) in model development period.** In these model rankings, we summarise proper scores, predictive performance, calibration, and outbreak detection capabilities of forecasting models. Models are arranged by mean weighted interval score (WIS), from best (lowest) to worst (highest). Bias measures the bias of each models' predictive quantiles, PI coverage measures the coverage of the 95% prediction intervals (PIs), $R^2$ denotes the proportion of variation in log(Cases + 1) explained by the models' posterior median estimates, and the corresponding root mean square error (RMSE) for log(Cases + 1) by using the posterior median. Details of the model abbreviations are provided in Table SI 2. All ensemble models are highlighted with an asterisk. As some forecasting models require a minimum number of observations, these comparisons are from the first month at which all forecasting models can make a prediction (August 2014) until the end of the model development period (December 2017).

| | Model | WIS | Bias | PI Coverage | $R^2$ | RMSE |
|---|---|---|---|---|---|---|
| 1 | TimeGPT | 0.41 | -0.04 | 93.2% | 0.84 | 0.86 |
| 2 | Median-NoBayes * | 0.45 | -0.16 | 94.4% | 0.82 | 0.95 |
| 3 | Median * | 0.46 | -0.22 | 93.9% | 0.82 | 0.97 |
| 4 | Ew-Mean-NoBayes * | 0.47 | -0.13 | 90.1% | 0.82 | 0.96 |
| 5 | Median-NoBase * | 0.49 | -0.25 | 95.5% | 0.80 | 1.04 |
| 6 | Median-NoCov * | 0.49 | -0.10 | 90.6% | 0.79 | 1.01 |
| 7 | SARIMA | 0.50 | -0.04 | 87.5% | 0.79 | 1.02 |
| 8 | EW-Mean * | 0.51 | -0.19 | 89.5% | 0.79 | 1.08 |
| 9 | EW-Mean-NoCov * | 0.52 | -0.16 | 88.7% | 0.79 | 1.06 |
| 10 | EW-Mean-NoBase * | 0.55 | -0.21 | 90.9% | 0.76 | 1.18 |
| 11 | Baseline | 0.57 | 0.01 | 78.2% | 0.79 | 1.00 |
| 12 | TCN | 0.59 | -0.22 | 87.3% | 0.75 | 1.15 |
| 13 | TimeGPT-NoCov | 0.89 | -0.32 | 88.7% | 0.46 | 1.95 |
| 14 | Bayes-Climate | 1.03 | -0.41 | 76.3% | 0.28 | 2.39 |

#### Supplementary Material 9.3.2. Model development period – Individual forecasting models

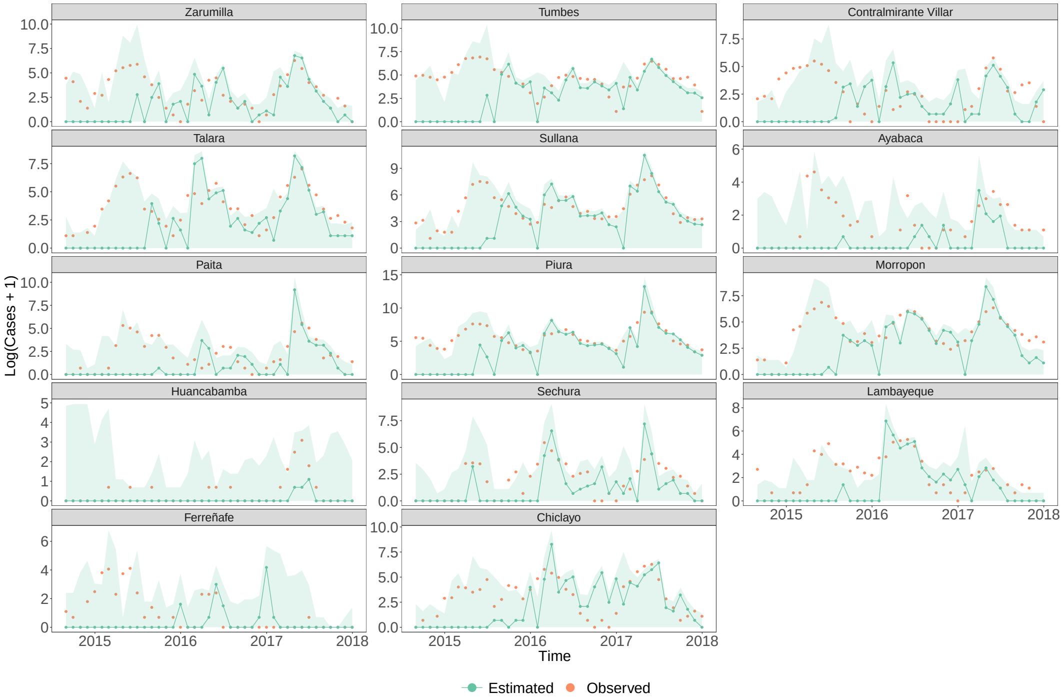


Figure SI 22: **Bayesian climate model's predictive performance across the model development period.** For each province, the observed cases (on a logarithmic scale) is shown (orange) alongside the corresponding posterior median estimates (green) and the 95% PIs (shaded green). The provinces (facets) are sorted in descending order by latitude (from north to south), and y-axes are province-dependent due to differences in each province's average size of epidemics. As some forecasting models require a minimum number of observations, for equitable comparison across models for the model development period, we present results from the first month at which all forecasting models can make a prediction (August 2014) until the end of the model development period (December 2017).

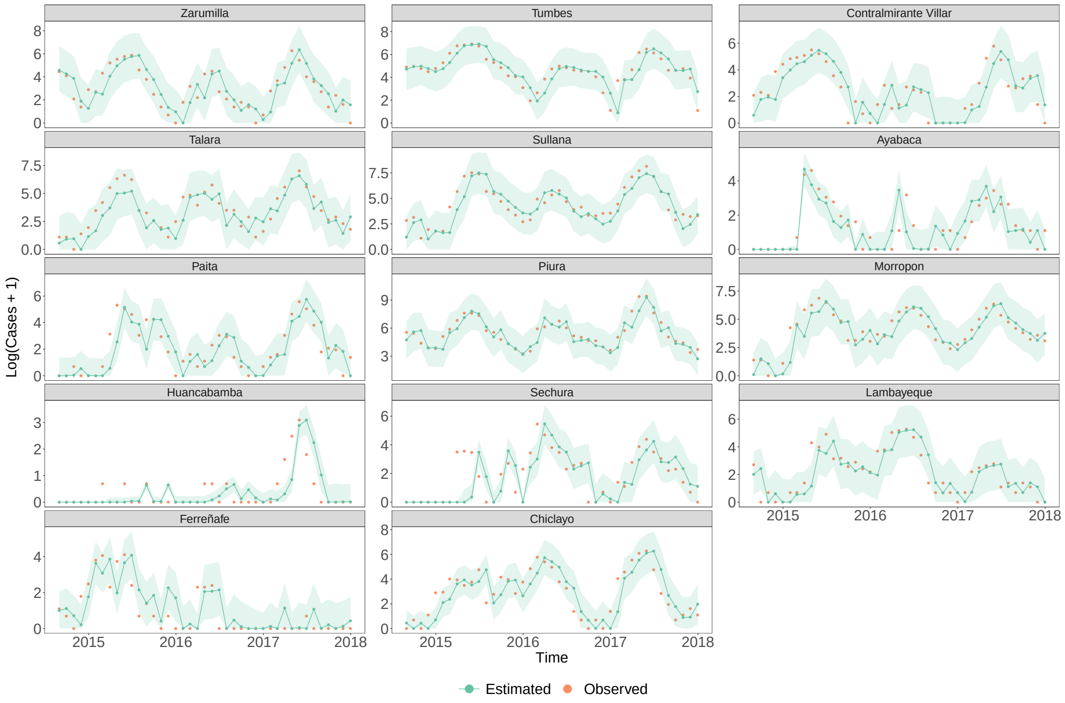


Figure SI 23: **SARIMA models' predictive performance across the model development period.** For each province, the observed cases (on a logarithmic scale) is shown (orange) alongside the corresponding posterior median estimates (green) and the 95% PIs (shaded green). The provinces (facets) are sorted in descending order by latitude (from north to south), and y-axes are province-dependent due to differences in each province's average size of epidemics. As some forecasting models require a minimum number of observations, for equitable comparison across models for the model development period, we present results from the first month at which all forecasting models can make a prediction (August 2014) until the end of the model development period (December 2017).

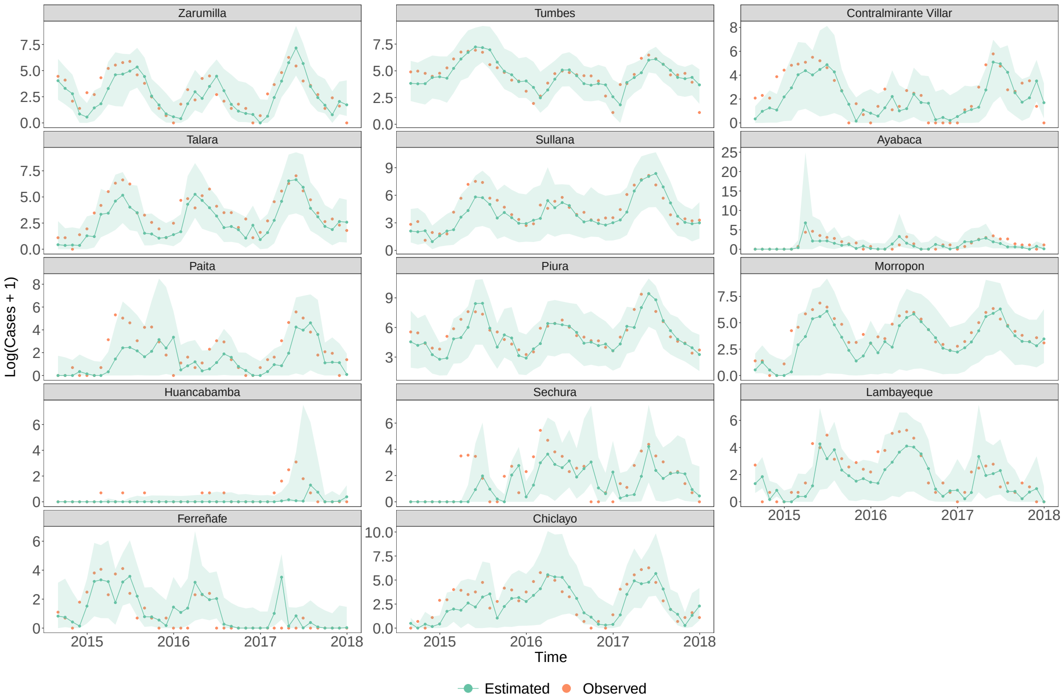


Figure SI 24: **TCN models' predictive performance across the model development period.** For each province, the observed cases (on a logarithmic scale) is shown (orange) alongside the corresponding posterior median estimates (green) and the 95% PIs (shaded green). The provinces (facets) are sorted in descending order by latitude (from north to south), and y-axes are province-dependent due to differences in each province's average size of epidemics. As some forecasting models require a minimum number of observations, for equitable comparison across models for the model development period, we present results from the first month at which all forecasting models can make a prediction (August 2014) until the end of the model development period (December 2017).

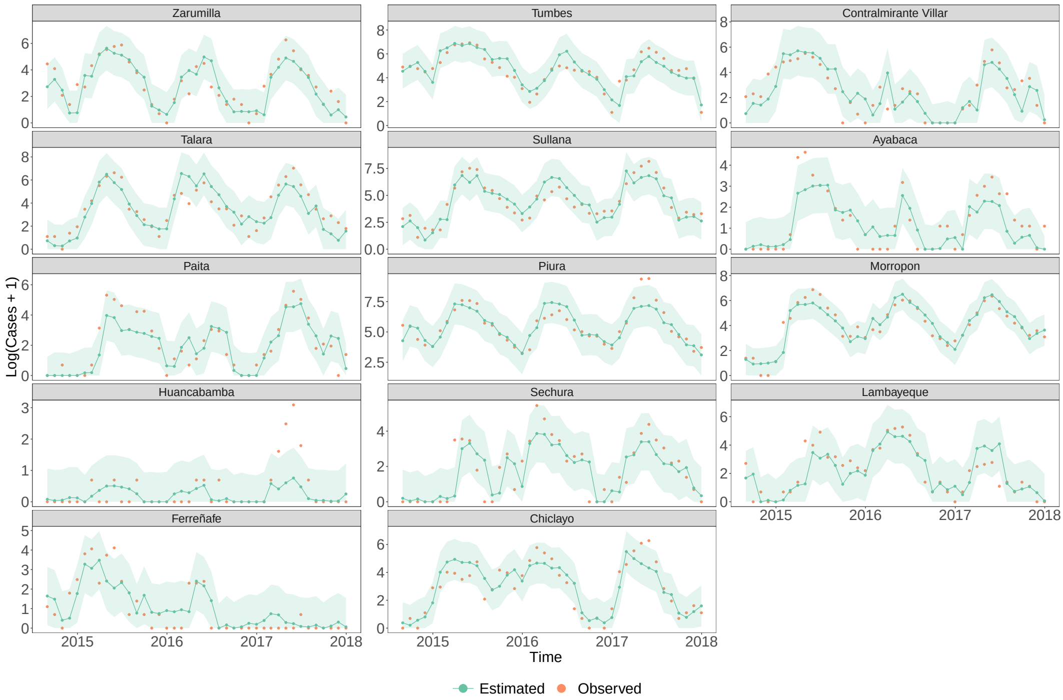


Figure SI 25: **TimeGPT model's predictive performance across the model development period.** For each province, the observed cases (on a logarithmic scale) is shown (orange) alongside the corresponding posterior median estimates (green) and the 95% PIs (shaded green). The provinces (facets) are sorted in descending order by latitude (from north to south), and y-axes are province-dependent due to differences in each province's average size of epidemics. As some forecasting models require a minimum number of observations, for equitable comparison across models for the model development period, we present results from the first month at which all forecasting models can make a prediction (August 2014) until the end of the model development period (December 2017).

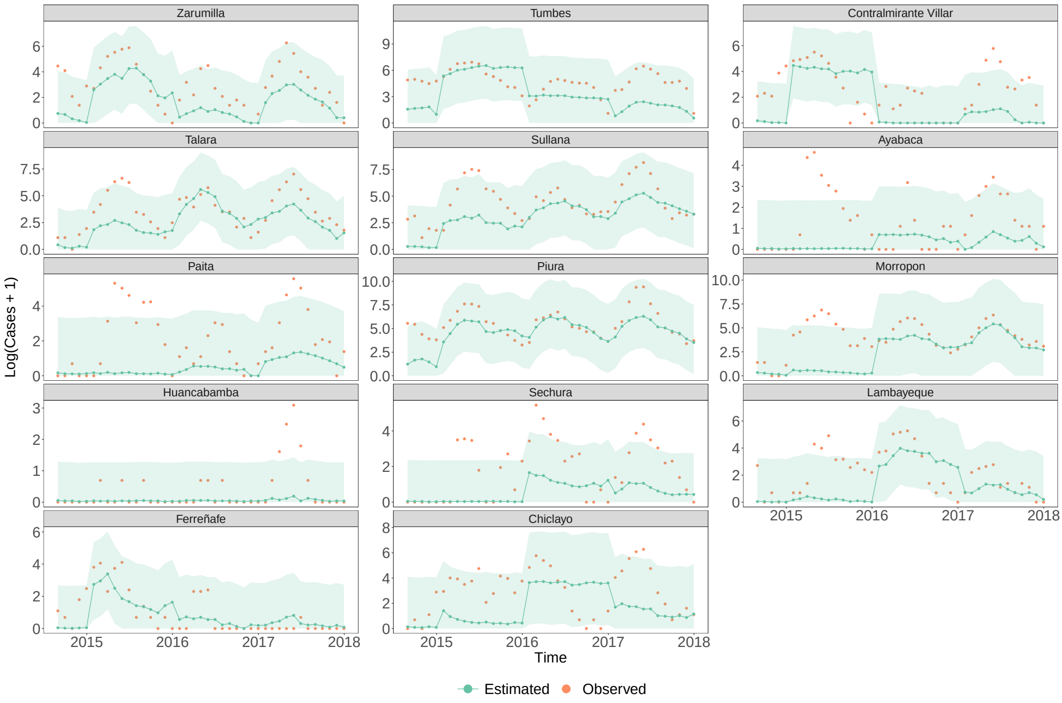


Figure SI 26: **Predictive performance of TimeGPT model without covariates across the model development period.** For each province, the observed cases (on a logarithmic scale) is shown (orange) alongside the corresponding posterior median estimates (green) and the 95% PIs (shaded green). The provinces (facets) are sorted in descending order by latitude (from north to south), and y-axes are province-dependent due to differences in each province's average size of epidemics. As some forecasting models require a minimum number of observations, for equitable comparison across models for the model development period, we present results from the first month at which all forecasting models can make a prediction (August 2014) until the end of the model development period (December 2017).

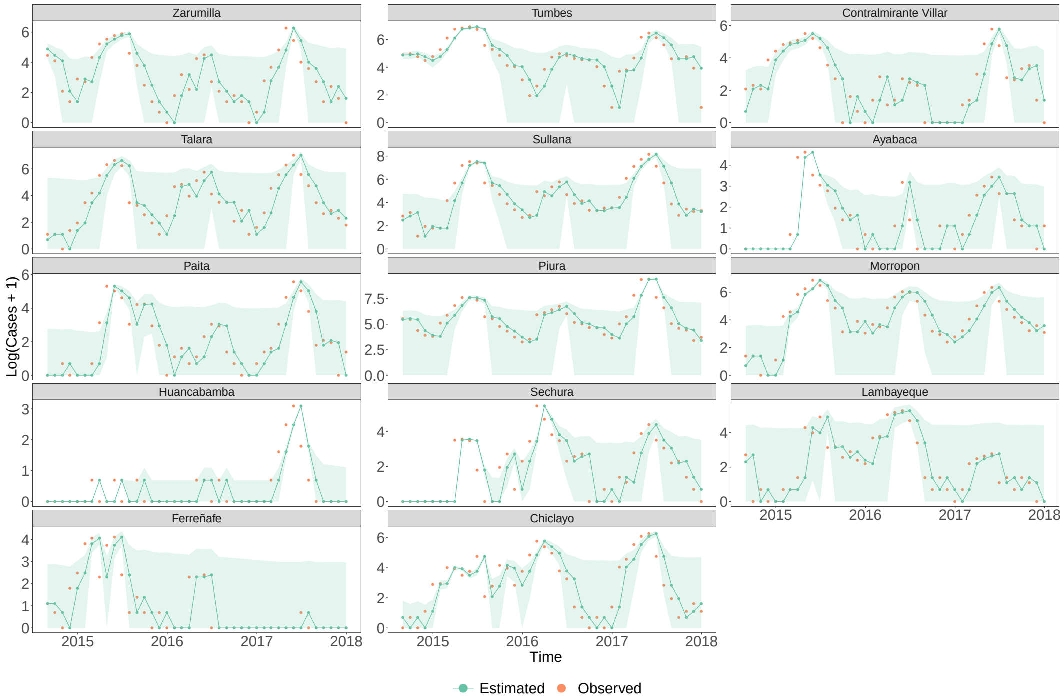


Figure SI 27: **Baseline model's predictive performance across the model development period.** We visualise results obtained for the epidemiologically naive baseline forecaster. For each province, the observed cases (on a logarithmic scale) is shown (orange) alongside the corresponding posterior median estimates (green) and the 95% PIs (shaded green). The provinces (facets) are sorted in descending order by latitude (from north to south), and y-axes are province-dependent due to differences in each province's average size of epidemics. As some forecasting models require a minimum number of observations, for equitable comparison across models for the model development period, we present results from the first month at which all forecasting models can make a prediction (August 2014) until the end of the model development period (December 2017).

#### Supplementary Material 9.3.3. Model development period — Coverage

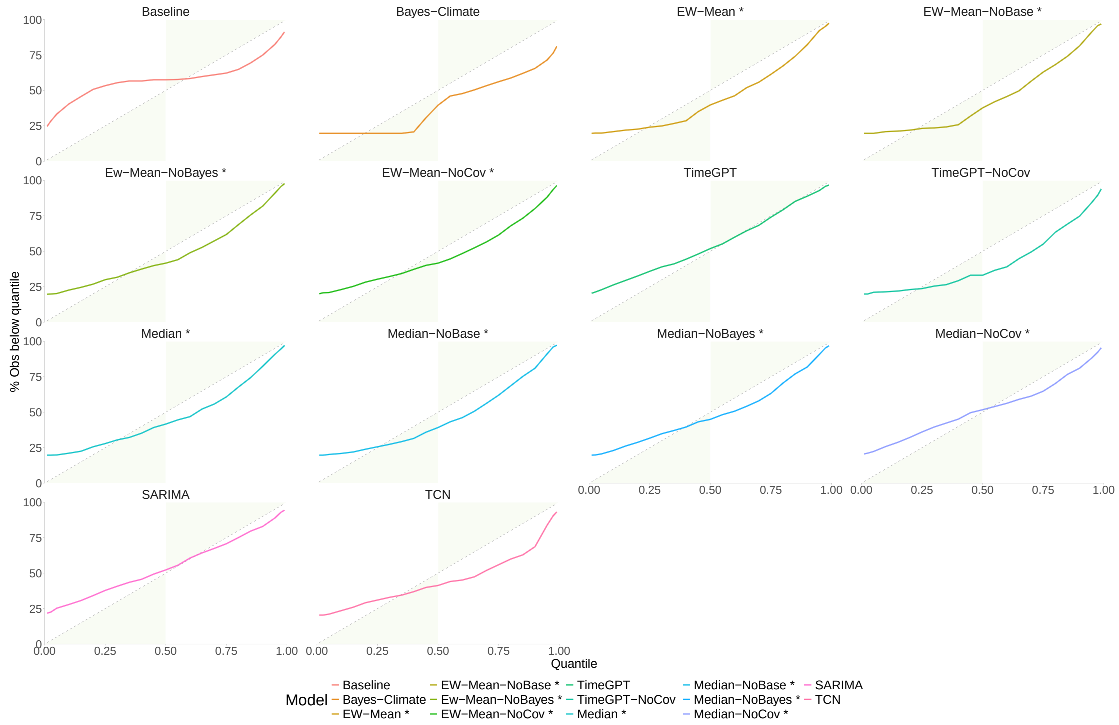


Figure SI 28: **Quantile coverage for quantile-based forecasting models across the model development period.** For each forecasting model, the quantile coverage is plotted where the y-axis denotes the percentage of observed values (log(Cases + 1)) that lie below the corresponding model-based predictive quantile level shown on the x-axis. Quantile coverage can be interpreted as a measure of probabilistic calibration i.e. consistency of a model's probabilities with those actually observed. The green shaded polygons denote areas of conservative behaviour i.e. empirical coverage outweighs the nominal coverage required. Details of the model abbreviations are provided in Table SI 2. As some forecasting models require a minimum number of observations, these comparisons are from the first month at which all forecasting models can make a prediction (August 2014) until the end of the model development period (December 2017).

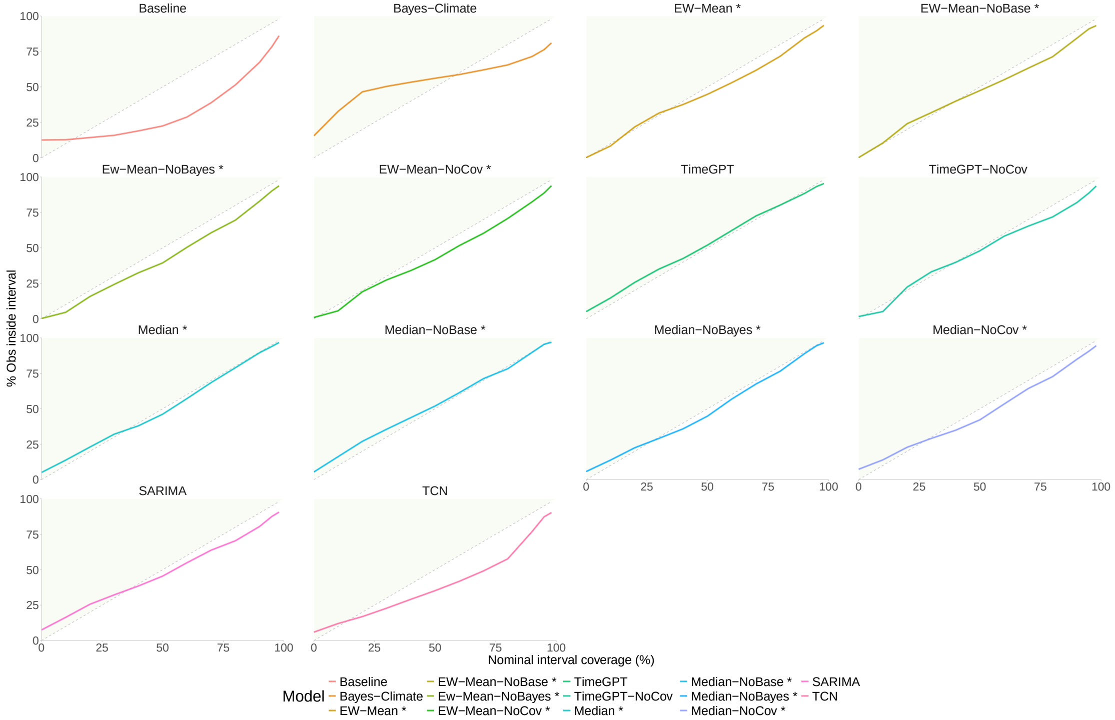


Figure SI 29: **Interval coverage for quantile-based forecasting models across the model development period.** For each forecasting model, the interval coverage is plotted where the y-axis denotes the percentage of observed values ((log(Cases + 1))) that lie below the corresponding model-based predictive quantile level shown on the x-axis. Interval coverage can be interpreted as a measure of probabilistic calibration i.e. consistency of a model's probabilities with those actually observed. The green shaded polygons denote areas of conservative behaviour i.e. empirical coverage outweighs the nominal coverage required. Interval coverage alone is not a measure of forecasting skill, as it only focuses on probabilistic calibration, and thus, we also report values of a proper scoring rule, the weighted interval score (WIS) which measures calibration and sharpness simultaneously. Details of the model abbreviations are provided in Table SI 2. As some forecasting models require a minimum number of observations, these comparisons are from the first month at which all forecasting models can make a prediction (August 2014) until the end of the model development period (December 2017).

#### Supplementary Material 9.3.4. Testing period – Summary tables

Table SI 9: **Results for forecasting log(Cases + 1) in the testing period of 2018 to 2021.** We summarise proper scores, predictive performance, calibration, and outbreak detection capabilities of forecasting models. Models are arranged by mean weighted interval score (WIS), from best (lowest) to worst (highest). Bias measures the bias of each models' predictive quantiles, PI coverage measures the coverage of the 95% prediction intervals (PIs), $R^2$ denotes the proportion of variation in log(Cases + 1) explained by the models' posterior median estimates, and the corresponding root mean squared error (RMSE) for log(Cases + 1) by using the posterior median. For detection of outbreak onset, *Sensitivity*, *Specificity*, and *AUC* represent the True Positive rate, True Negative rate, and Area Under the Curve in terms of forecasting skill for correctly classifying the first possible future outbreaks of DIR $\geq$ 150 per 100,000. Table 2 is the analogous table which depicts outbreak detection for DIR first surpassing a threshold of 50 per 100,000. Details of the model abbreviations are provided in Table SI 2. All ensemble models are highlighted with an asterisk.

| | Model | WIS | Bias | PI Coverage | $R^2$ | RMSE | Sensitivity | Specificity | AUC |
|---|---|---|---|---|---|---|---|---|---|
| 1 | Median * | 0.34 | 0.03 | 95.2% | 0.74 | 0.53 | 80.0% | 94.9% | 0.87 |
| 2 | Median-NoBase * | 0.35 | 0.02 | 94.8% | 0.73 | 0.54 | 80.0% | 95.3% | 0.88 |
| 3 | Median-NoBayes * | 0.35 | 0.06 | 94.8% | 0.74 | 0.54 | 80.0% | 94.5% | 0.87 |
| 4 | EW-Mean * | 0.35 | 0.17 | 87.8% | 0.75 | 0.54 | 80.0% | 93.7% | 0.87 |
| 5 | Median-NoCov * | 0.36 | 0.04 | 95.2% | 0.73 | 0.55 | 80.0% | 93.9% | 0.87 |
| 6 | Ew-Mean-NoBayes * | 0.36 | 0.21 | 87.5% | 0.74 | 0.55 | 60.0% | 95.0% | 0.78 |
| 7 | EW-Mean-NoCov * | 0.36 | 0.14 | 91.2% | 0.74 | 0.55 | 60.0% | 90.4% | 0.75 |
| 8 | Trained * | 0.36 | 0.07 | 92.4% | 0.72 | 0.56 | 80.0% | 93.4% | 0.87 |
| 9 | EW-Mean-NoBase * | 0.36 | 0.16 | 87.1% | 0.74 | 0.55 | 60.0% | 97.3% | 0.79 |
| 10 | SARIMA | 0.36 | 0.03 | 93.5% | 0.73 | 0.56 | 80.0% | 94.2% | 0.87 |
| 11 | Prov-Trained * | 0.37 | 0.05 | 91.1% | 0.72 | 0.56 | 80.0% | 94.7% | 0.87 |
| 12 | Baseline | 0.38 | 0.04 | 97.3% | 0.74 | 0.52 | 60.0% | 55.0% | 0.57 |
| 13 | TCN | 0.39 | 0.11 | 91.2% | 0.70 | 0.59 | 80.0% | 94.1% | 0.87 |
| 14 | TimeGPT | 0.41 | 0.08 | 88.1% | 0.69 | 0.62 | 60.0% | 94.1% | 0.77 |
| 15 | TimeGPT-NoCov | 0.42 | 0.12 | 85.6% | 0.67 | 0.62 | 40.0% | 95.3% | 0.68 |
| 16 | Bayes-Climate | 0.52 | -0.20 | 87.6% | 0.58 | 0.71 | 40.0% | 88.6% | 0.64 |

Table SI 10: **Detection of all outbreaks with DIR ≥ 50 per 100,000 across all months of 2018 to 2021.** For detection of any outbreak months, *Sensitivity*, *Specificity*, and *AUC* represent the True Positive rate, True Negative rate, and Area Under the Curve in terms of forecasting skill for correctly classifying future outbreaks of DIR ≥ 50 per 100,000. Table SI 11 is the analogous table which depicts outbreak detection for DIR reaching a threshold of 150 per 100,000. Details of the model abbreviations are provided in Table SI 2. All ensemble models are highlighted with an asterisk. Models are presented in the same ordering as Table 2 to enable comparison with the models' performance across other performance metrics.

| | Model | Sensitivity | Specificity | AUC |
|---|---|---|---|---|
| 1 | Median * | 91.7% | 95.2% | 0.93 |
| 2 | Median-NoBase * | 89.6% | 95.4% | 0.92 |
| 3 | Median-NoBayes * | 91.7% | 94.7% | 0.93 |
| 4 | EW-Mean * | 89.6% | 96.5% | 0.93 |
| 5 | Median-NoCov * | 91.7% | 92.5% | 0.92 |
| 6 | Ew-Mean-NoBayes * | 89.6% | 96.8% | 0.93 |
| 7 | EW-Mean-NoCov * | 85.4% | 96.3% | 0.91 |
| 8 | Trained * | 91.7% | 93.6% | 0.93 |
| 9 | EW-Mean-NoBase * | 89.6% | 96.3% | 0.93 |
| 10 | SARIMA | 89.6% | 95.8% | 0.93 |
| 11 | Prov-Trained * | 89.6% | 95.8% | 0.93 |
| 12 | Baseline | 81.2% | 96.2% | 0.89 |
| 13 | TCN | 89.6% | 96.3% | 0.93 |
| 14 | TimeGPT | 79.2% | 95.0% | 0.87 |
| 15 | TimeGPT-NoCov | 87.5% | 88.0% | 0.88 |
| 16 | Bayes-Climate | 64.6% | 94.1% | 0.79 |

Table SI 11: **Detection of all outbreaks with DIR ≥ 150 per 100,000 across all months of 2018 to 2021.** For detection of any outbreak months, *Sensitivity*, *Specificity*, and *AUC* represent the True Positive rate, True Negative rate, and Area Under the Curve in terms of forecasting skill for correctly classifying future outbreaks of DIR ≥ 150 per 100,000. Table SI 10 is the analogous table which depicts outbreak detection for DIR reaching a threshold of 50 per 100,000. Details of the model abbreviations are provided in Table SI 2. All ensemble models are highlighted with an asterisk. Models are presented in the same ordering as Table 2 to enable comparison with the models' performance across other performance metrics.

| | Model | Sensitivity | Specificity | AUC |
|---|---|---|---|---|
| 1 | Median * | 85.7% | 95.3% | 0.91 |
| 2 | Median-NoBase * | 85.7% | 95.0% | 0.90 |
| 3 | Median-NoBayes * | 85.7% | 97.1% | 0.91 |
| 4 | EW-Mean * | 85.7% | 96.4% | 0.91 |
| 5 | Median-NoCov * | 85.7% | 93.2% | 0.89 |
| 6 | Ew-Mean-NoBayes * | 85.7% | 96.2% | 0.91 |
| 7 | EW-Mean-NoCov * | 78.6% | 96.8% | 0.88 |
| 8 | Trained * | 85.7% | 95.9% | 0.91 |
| 9 | EW-Mean-NoBase * | 85.7% | 95.9% | 0.91 |
| 10 | SARIMA | 78.6% | 96.0% | 0.87 |
| 11 | Prov-Trained * | 85.7% | 94.8% | 0.90 |
| 12 | Baseline | 64.3% | 98.5% | 0.81 |
| 13 | TCN | 85.7% | 96.2% | 0.91 |
| 14 | TimeGPT | 78.6% | 95.1% | 0.87 |
| 15 | TimeGPT-NoCov | 71.4% | 92.9% | 0.82 |
| 16 | Bayes-Climate | 71.4% | 93.0% | 0.82 |

Table SI 12: **Results for forecasting DIR in the testing period of 2018 to 2021.** We summarise proper scores, predictive performance, calibration, and outbreak detection capabilities of forecasting models. Models are arranged by mean weighted interval score (WIS), from best (lowest) to worst (highest). Bias measures the bias of each models' predictive quantiles, PI coverage measures the coverage of the 95% prediction intervals (PIs), $R^2$ denotes the proportion of variation in DIR explained by the models' posterior median estimates, and the corresponding root mean square error (RMSE) for log(Cases + 1) by using the posterior median. For detection of outbreak onset, *Sensitivity*, *Specificity*, and *AUC* represent the True Positive rate, True Negative rate, and Area Under the Curve in terms of forecasting skill for correctly classifying the first possible future outbreaks of DIR $\geq$ 50 per 100,000. Table 2 is the analogous table which depicts these model rankings for the logarithm of cases. Details of the model abbreviations are provided in Table SI 2. All ensemble models are highlighted with an asterisk.

| | Model | WIS | Bias | PI Coverage | $R^2$ | RMSE | Sensitivity | Specificity | AUC |
|---|---|---|---|---|---|---|---|---|---|
| 1 | Median * | 4.95 | 0.04 | 95.2% | 0.68 | 21.97 | 81.8% | 94.3% | 0.88 |
| 2 | Median-NoBayes * | 5.01 | 0.05 | 94.9% | 0.68 | 22.13 | 81.8% | 87.2% | 0.85 |
| 3 | Median-NoCov * | 5.11 | 0.04 | 95.4% | 0.65 | 22.99 | 81.8% | 91.0% | 0.86 |
| 4 | Ew-Mean-NoBayes * | 5.16 | 0.25 | 88.1% | 0.69 | 21.59 | 63.6% | 97.3% | 0.80 |
| 5 | Median-NoBase * | 5.18 | 0.02 | 94.8% | 0.67 | 22.43 | 81.8% | 95.7% | 0.89 |
| 6 | EW-Mean * | 5.19 | 0.24 | 88.7% | 0.68 | 22.79 | 72.7% | 95.9% | 0.84 |
| 7 | EW-Mean-NoCov * | 5.25 | 0.17 | 91.4% | 0.64 | 23.26 | 81.8% | 86.5% | 0.84 |
| 8 | EW-Mean-NoBase * | 5.36 | 0.23 | 87.5% | 0.66 | 23.64 | 81.8% | 94.7% | 0.88 |
| 9 | Trained * | 5.36 | 0.12 | 92.1% | 0.67 | 22.57 | 81.8% | 95.4% | 0.89 |
| 10 | SARIMA | 5.40 | 0.03 | 93.5% | 0.67 | 23.27 | 63.6% | 96.1% | 0.80 |
| 11 | Prov-Trained * | 5.49 | 0.09 | 90.6% | 0.66 | 24.28 | 72.7% | 89.9% | 0.81 |
| 12 | TCN | 5.67 | 0.11 | 91.2% | 0.59 | 27.84 | 72.7% | 96.6% | 0.85 |
| 13 | Baseline | 6.17 | 0.01 | 97.3% | 0.64 | 24.13 | 90.9% | 45.0% | 0.68 |
| 14 | TimeGPT-NoCov | 6.39 | 0.12 | 85.6% | 0.47 | 28.32 | 72.7% | 91.5% | 0.82 |
| 15 | TimeGPT | 7.26 | 0.08 | 88.1% | 0.54 | 31.29 | 90.9% | 91.7% | 0.91 |
| 16 | Bayes-Climate | 8.27 | -0.22 | 87.1% | 0.38 | 53.12 | 54.5% | 90.1% | 0.72 |

Table SI 13: **Results for forecasting DIR in the testing period of 2018 to 2021.** This table is analogous to Table SI 12, but here we report capabilities for detection of first possible future outbreaks of DIR $\geq$ 150 per 100,000. Details of the model abbreviations are provided in Table SI 2. All ensemble models are highlighted with an asterisk.

| | Model | WIS | Bias | PI Coverage | $R^2$ | RMSE | Sensitivity | Specificity | AUC |
|---|---|---|---|---|---|---|---|---|---|
| 1 | Median * | 4.95 | 0.04 | 95.2% | 0.68 | 21.97 | 80.0% | 94.9% | 0.87 |
| 2 | Median-NoBayes * | 5.01 | 0.05 | 94.9% | 0.68 | 22.13 | 80.0% | 94.5% | 0.87 |
| 3 | Median-NoCov * | 5.11 | 0.04 | 95.4% | 0.65 | 22.99 | 80.0% | 93.9% | 0.87 |
| 4 | Ew-Mean-NoBayes * | 5.16 | 0.25 | 88.1% | 0.69 | 21.59 | 60.0% | 95.0% | 0.78 |
| 5 | Median-NoBase * | 5.18 | 0.02 | 94.8% | 0.67 | 22.43 | 80.0% | 95.3% | 0.88 |
| 6 | EW-Mean * | 5.19 | 0.24 | 88.7% | 0.68 | 22.79 | 80.0% | 93.7% | 0.87 |
| 7 | EW-Mean-NoCov * | 5.25 | 0.17 | 91.4% | 0.64 | 23.26 | 60.0% | 90.4% | 0.75 |
| 8 | EW-Mean-NoBase * | 5.36 | 0.23 | 87.5% | 0.66 | 23.64 | 60.0% | 97.3% | 0.79 |
| 9 | Trained * | 5.36 | 0.12 | 92.1% | 0.67 | 22.57 | 80.0% | 93.4% | 0.87 |
| 10 | SARIMA | 5.40 | 0.03 | 93.5% | 0.67 | 23.27 | 80.0% | 94.2% | 0.87 |
| 11 | Prov-Trained * | 5.49 | 0.09 | 90.6% | 0.66 | 24.28 | 80.0% | 94.7% | 0.87 |
| 12 | TCN | 5.67 | 0.11 | 91.2% | 0.59 | 27.84 | 80.0% | 94.1% | 0.87 |
| 13 | Baseline | 6.17 | 0.01 | 97.3% | 0.64 | 24.13 | 60.0% | 55.0% | 0.57 |
| 14 | TimeGPT-NoCov | 6.39 | 0.12 | 85.6% | 0.47 | 28.32 | 40.0% | 95.3% | 0.68 |
| 15 | TimeGPT | 7.26 | 0.08 | 88.1% | 0.54 | 31.29 | 60.0% | 94.1% | 0.77 |
| 16 | Bayes-Climate | 8.27 | -0.22 | 87.1% | 0.38 | 53.12 | 40.0% | 88.6% | 0.64 |

#### Supplementary Material 9.3.5. Testing period – Individual forecasters

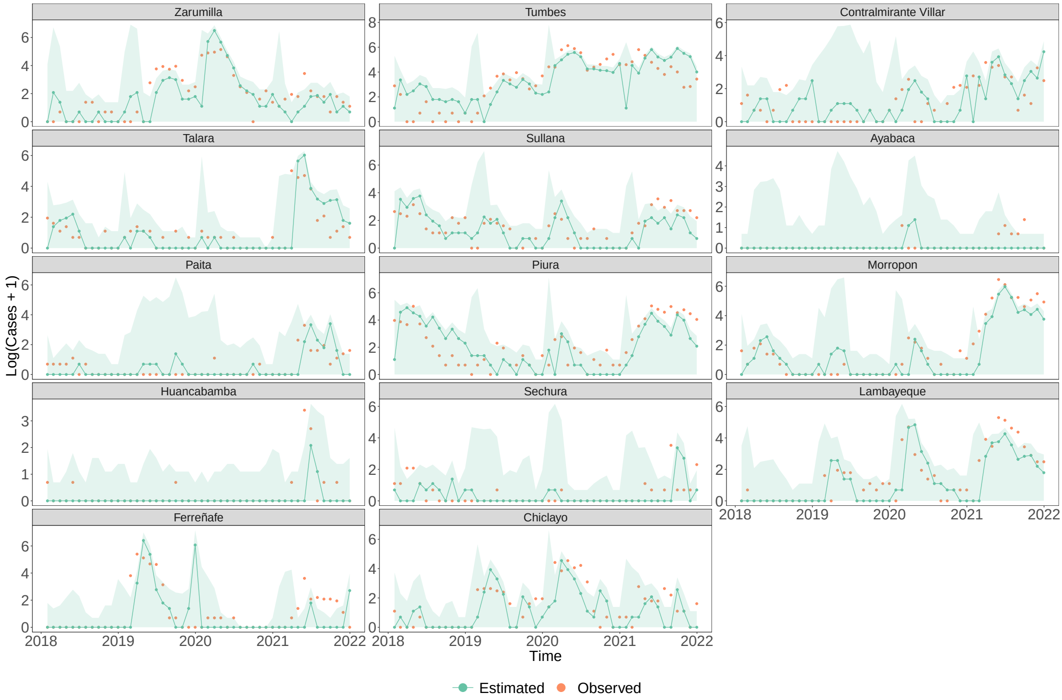


Figure SI 30: **Bayesian climate model's predictive performance across the testing period (2018–2021).** For each province, the observed cases (on a logarithmic scale) is shown (orange) alongside the corresponding posterior median estimates (green) and the 95% PIs (shaded green) from one-month-ahead forecasts. The provinces (facets) are sorted in descending order by latitude (from north to south), and y-axes are province-dependent due to differences in each province's average size of epidemics.

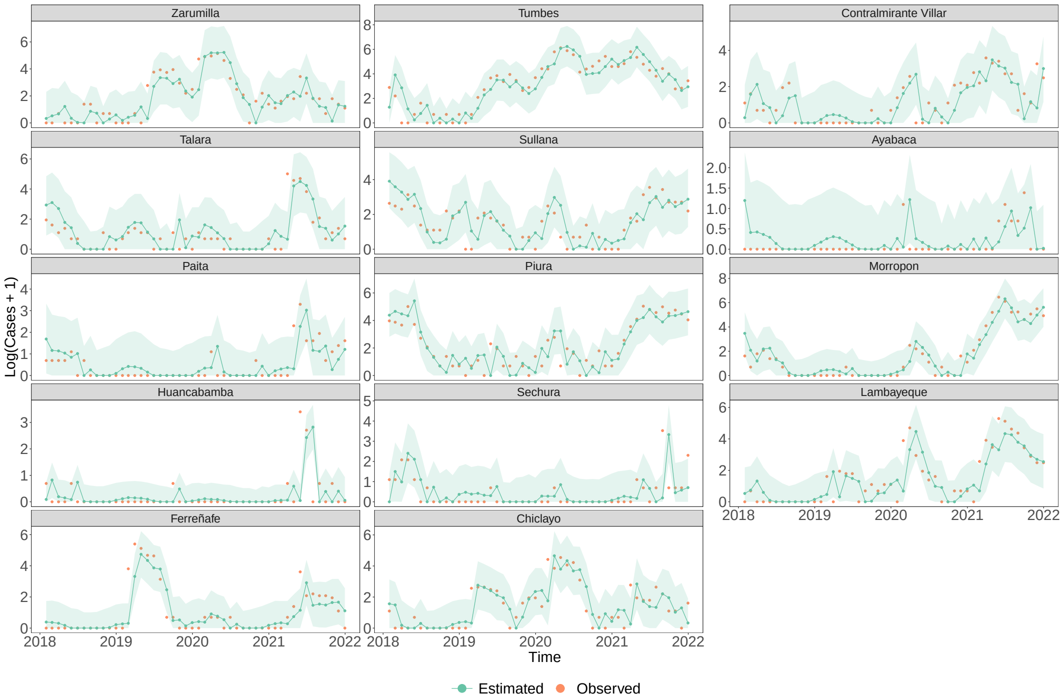


Figure SI 31: **SARIMA models' predictive performance across the testing period (2018–2021).** For each province, the observed cases (on a logarithmic scale) is shown (orange) alongside the corresponding posterior median estimates (green) and the 95% PIs (shaded green) from one-month-ahead forecasts. The provinces (facets) are sorted in descending order by latitude (from north to south), and y-axes are province-dependent due to differences in each province's average size of epidemics.

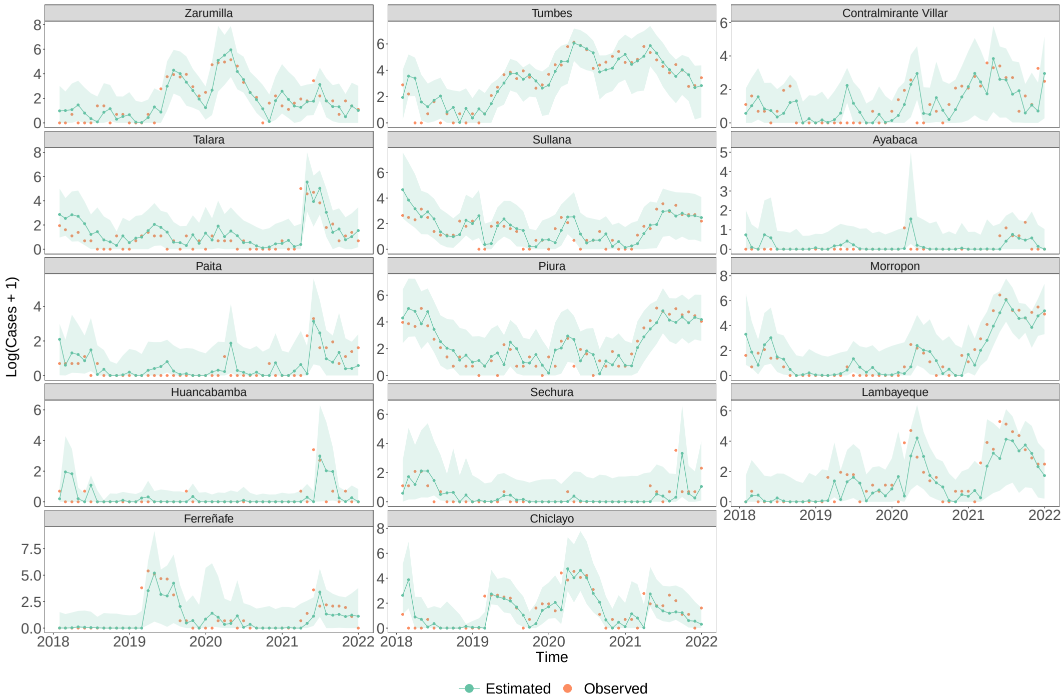


Figure SI 32: **TCN models' predictive performance across the testing period (2018–2021).** For each province, the observed cases (on a logarithmic scale) is shown (orange) alongside the corresponding posterior median estimates (green) and the 95% PIs (shaded green) from one-month-ahead forecasts. The provinces (facets) are sorted in descending order by latitude (from north to south), and y-axes are province-dependent due to differences in each province's average size of epidemics.

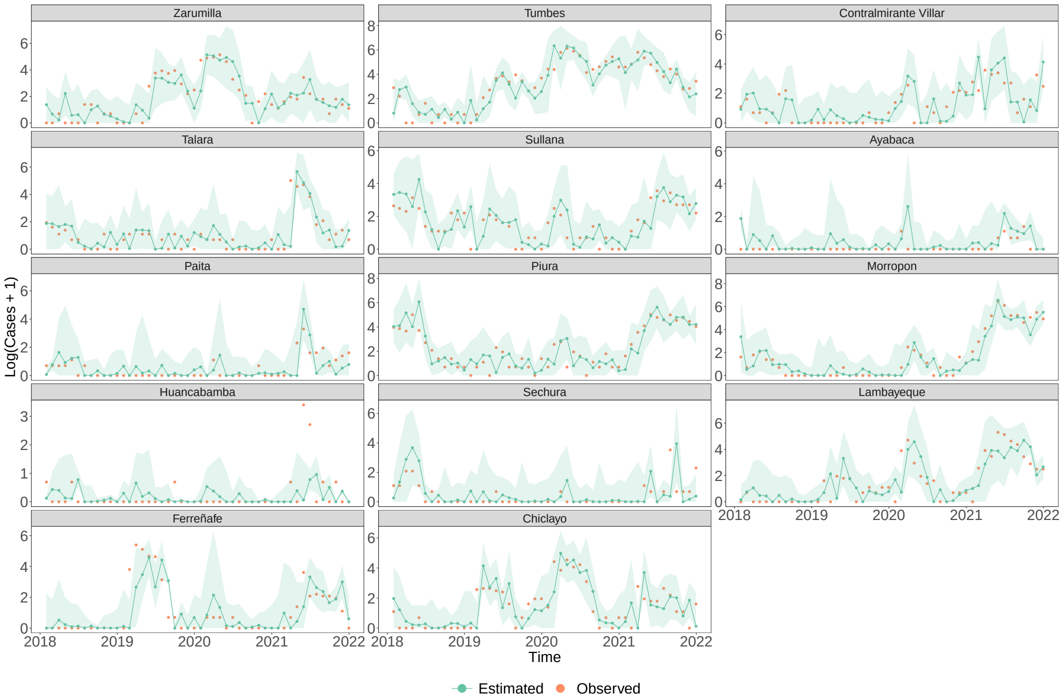


Figure SI 33: **TimeGPT model's predictive performance across the testing period (2018–2021).** For each province, the observed cases (on a logarithmic scale) is shown (orange) alongside the corresponding posterior median estimates (green) and the 95% PIs (shaded green) from one-month-ahead forecasts. The provinces (facets) are sorted in descending order by latitude (from north to south), and y-axes are province-dependent due to differences in each province's average size of epidemics.

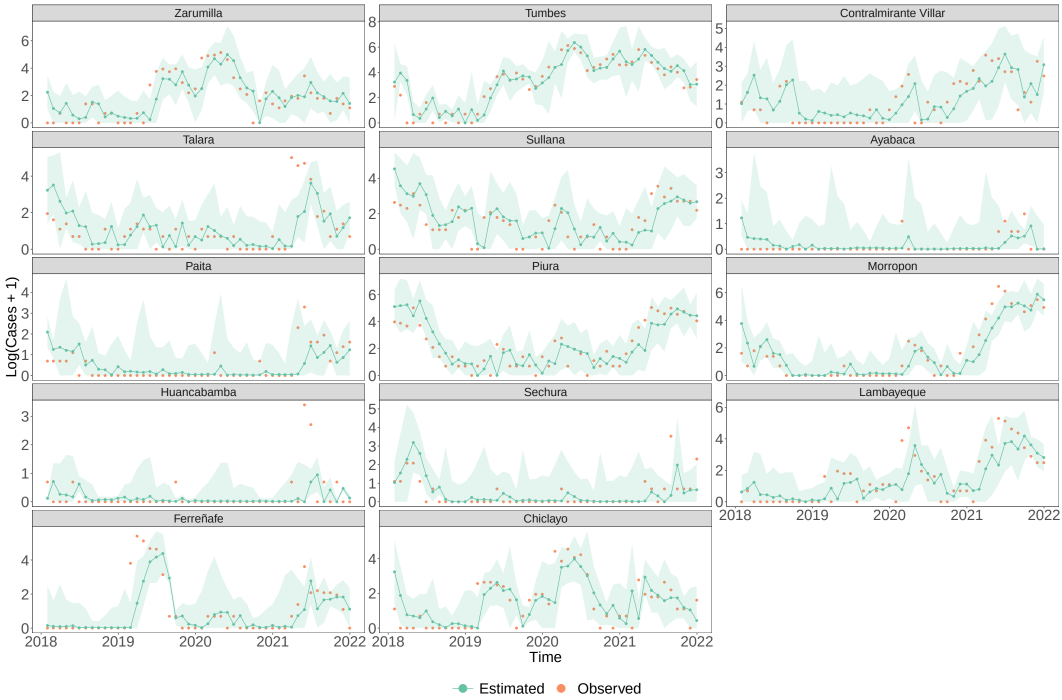


Figure SI 34: **Predictive performance of TimeGPT model without covariates across the testing period (2018–2021).** For each province, the observed cases (on a logarithmic scale) is shown (orange) alongside the corresponding posterior median estimates (green) and the 95% PIs (shaded green) from one-month-ahead forecasts. The provinces (facets) are sorted in descending order by latitude (from north to south), and y-axes are province-dependent due to differences in each province's average size of epidemics.

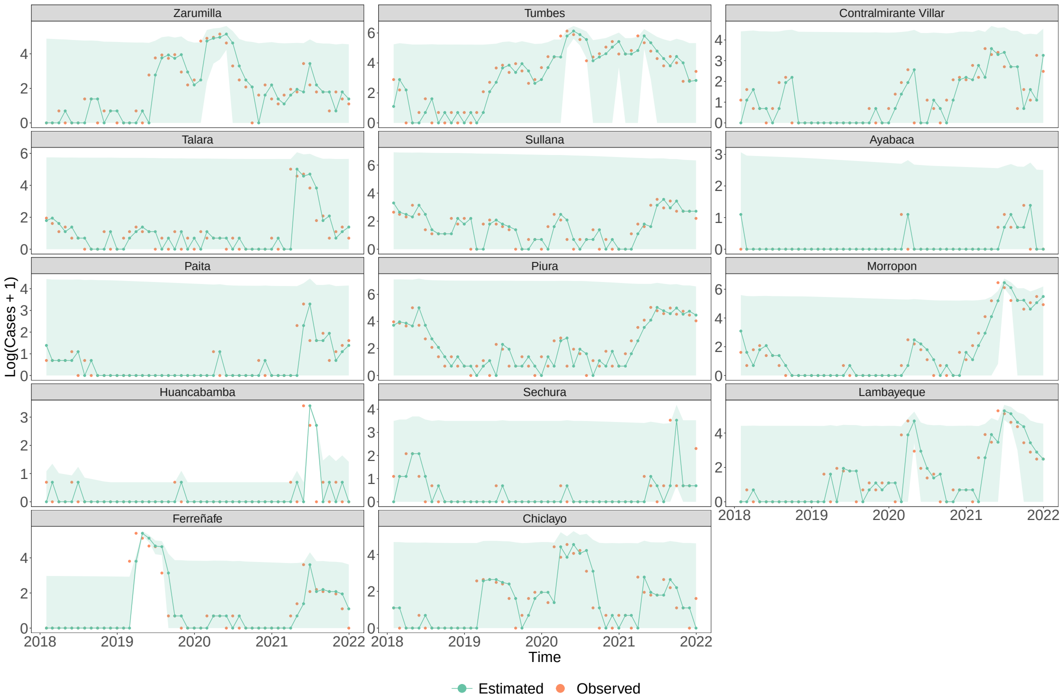


Figure SI 35: **Baseline model's predictive performance across the testing period (2018–2021).** For each province, the observed cases (on a logarithmic scale) is shown (orange) alongside the corresponding posterior median estimates (green) and the 95% PIs (shaded green) from one-month-ahead forecasts. The provinces (facets) are sorted in descending order by latitude (from north to south), and y-axes are province-dependent due to differences in each province's average size of epidemics.

#### Supplementary Material 9.3.6. Testing period – Ensemble models

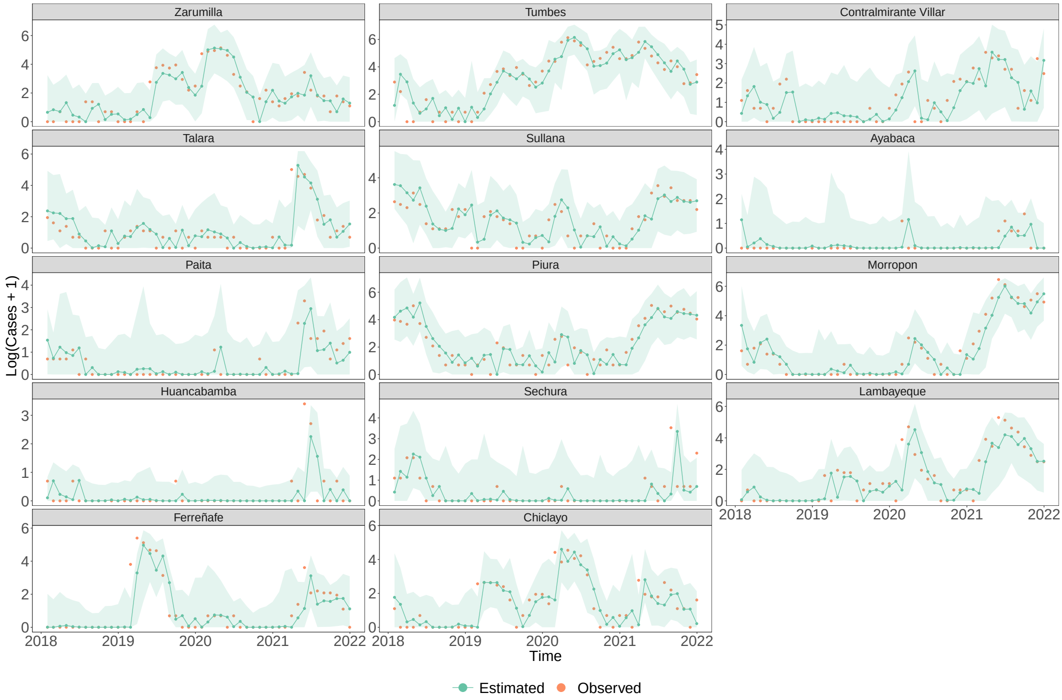


Figure SI 36: **Median ensemble model's predictive performance across the testing period (2018–2021).** For each province, the observed cases (on a logarithmic scale) is shown (orange) alongside the corresponding posterior median estimates (green) and the 95% PIs (shaded green) from one-month-ahead forecasts. The provinces (facets) are sorted in descending order by latitude (from north to south), and y-axes are province-dependent due to differences in each province's average size of epidemics.

#### Supplementary Material 9.3.7. Testing period – WIS

Table SI 14: **Summary statistics of WIS for log(Cases + 1) in the testing period of 2018 to 2021.** We summarise the weighted interval score (WIS) for each forecasting model by its median, lower quartile (Q1), and upper quartile (Q3). Models are arranged here by median WIS, where lower values of the WIS indicate superior forecasting performance. Details of the model abbreviations are provided in Table SI 2. All ensemble models are highlighted with an asterisk.

| | Model | Median WIS | Q1 | Q3 |
|---|---|---|---|---|
| 1 | Median-NoBayes * | 0.18 | 0.06 | 0.43 |
| 2 | Median-NoCov * | 0.18 | 0.06 | 0.44 |
| 3 | Median * | 0.18 | 0.06 | 0.42 |
| 4 | TCN | 0.18 | 0.05 | 0.54 |
| 5 | Median-NoBase * | 0.19 | 0.06 | 0.43 |
| 6 | EW-Mean-NoCov * | 0.19 | 0.08 | 0.43 |
| 7 | Prov-Trained * | 0.19 | 0.06 | 0.49 |
| 8 | Ew-Mean-NoBayes * | 0.20 | 0.08 | 0.43 |
| 9 | TimeGPT-NoCov | 0.20 | 0.07 | 0.53 |
| 10 | SARIMA | 0.20 | 0.07 | 0.41 |
| 11 | Trained * | 0.21 | 0.07 | 0.44 |
| 12 | EW-Mean-NoBase * | 0.21 | 0.07 | 0.45 |
| 13 | EW-Mean * | 0.22 | 0.08 | 0.43 |
| 14 | TimeGPT | 0.22 | 0.06 | 0.54 |
| 15 | Baseline | 0.22 | 0.06 | 0.51 |
| 16 | Bayes-Climate | 0.27 | 0.04 | 0.72 |

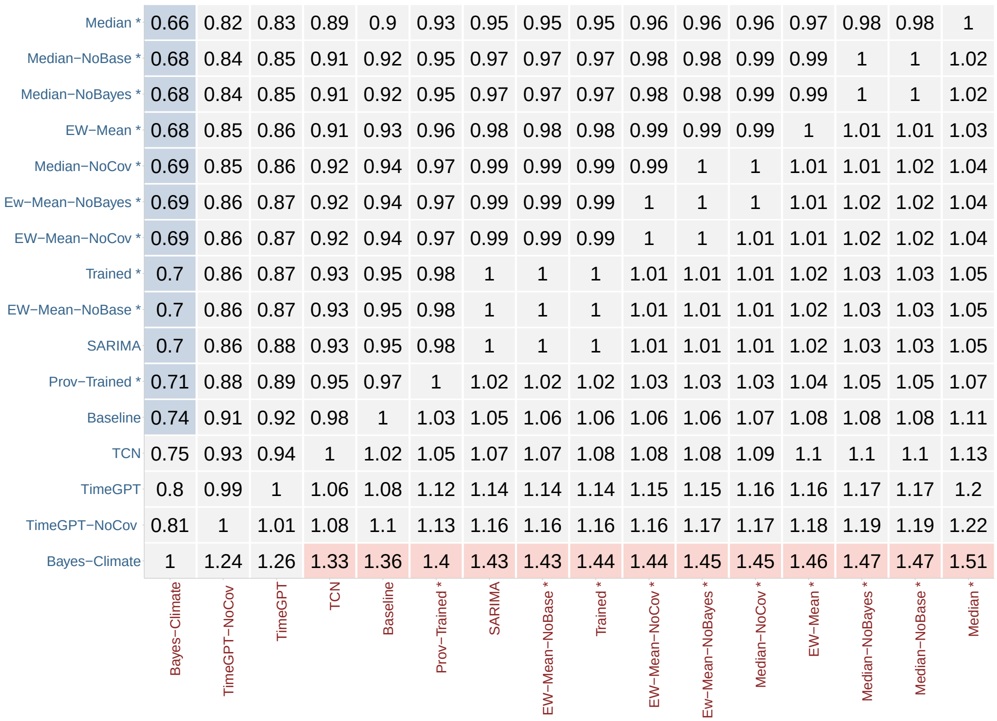


Figure SI 37: **WIS ratio between pairwise model combinations.** A visualisation of the pairwise ratio of Weighted Interval Score (WIS) values for forecasting models across 2018 to 2021. The focal model of interest is shown on the y-axis and the comparison models are shown on the x-axis. Then, the cell entries denote the ratio between the mean score of the focal model (numerator) and the mean score of the comparison model (denominator). Blue and red indicate superiority and inferiority of a model (as measured by the mean WIS) respectively. The p-value for each pairwise comparison is provided in Figure SI 38, whilst the model abbreviations are described in Table SI 2.

| | Bayes−Climate | TimeGPT−NoCov | TimeGPT | TCN | Baseline | Prov−Trained * | SARIMA | EW−Mean−NoBase * | Trained * | EW−Mean−NoCov * | Ew−Mean−NoBayes * | Median−NoCov * | EW−Mean * | Median−NoBayes * | Median−NoBase * | Median * |
|---|---|---|---|---|---|---|---|---|---|---|---|---|---|---|---|---|
| Median * | < 0.001 | < 0.001 | < 0.001 | < 0.001 | < 0.001 | < 0.001 | < 0.001 | < 0.001 | < 0.001 | < 0.001 | < 0.001 | < 0.001 | < 0.001 | 0.001 | < 0.001 | 1 |
| Median−NoBase * | < 0.001 | < 0.001 | < 0.001 | 0.005 | < 0.001 | 0.021 | 0.007 | < 0.001 | < 0.001 | 0.001 | < 0.001 | 0.128 | < 0.001 | 0.931 | 1 | < 0.001 |
| Median−NoBayes * | < 0.001 | < 0.001 | < 0.001 | 0.011 | < 0.001 | 0.021 | 0.002 | < 0.001 | < 0.001 | < 0.001 | < 0.001 | < 0.001 | < 0.001 | 1 | 0.931 | 0.001 |
| EW−Mean * | < 0.001 | < 0.001 | < 0.001 | 0.585 | < 0.001 | 0.895 | 0.32 | < 0.001 | 0.819 | 0.481 | 0.831 | 0.11 | 1 | < 0.001 | < 0.001 | < 0.001 |
| Median−NoCov * | < 0.001 | < 0.001 | < 0.001 | 0.297 | < 0.001 | 0.095 | 0.014 | 0.022 | 0.012 | < 0.001 | 0.229 | 1 | 0.11 | < 0.001 | 0.128 | < 0.001 |
| Ew−Mean−NoBayes * | < 0.001 | < 0.001 | < 0.001 | 0.22 | < 0.001 | 0.654 | 0.536 | 0.326 | 0.115 | 0.164 | 1 | 0.229 | 0.831 | < 0.001 | < 0.001 | < 0.001 |
| EW−Mean−NoCov * | < 0.001 | < 0.001 | < 0.001 | 0.987 | < 0.001 | 0.868 | 0.446 | 0.745 | 0.309 | 1 | 0.164 | < 0.001 | 0.481 | < 0.001 | 0.001 | < 0.001 |
| Trained * | < 0.001 | 0.001 | < 0.001 | 0.701 | 0.004 | 0.13 | 0.995 | 0.464 | 1 | 0.309 | 0.115 | 0.012 | 0.819 | < 0.001 | < 0.001 | < 0.001 |
| EW−Mean−NoBase * | < 0.001 | < 0.001 | < 0.001 | 0.763 | 0.02 | 0.525 | 0.053 | 1 | 0.464 | 0.745 | 0.326 | 0.022 | < 0.001 | < 0.001 | < 0.001 | < 0.001 |
| SARIMA | < 0.001 | < 0.001 | < 0.001 | 0.416 | 0.006 | 0.996 | 1 | 0.053 | 0.995 | 0.446 | 0.536 | 0.014 | 0.32 | 0.002 | 0.007 | < 0.001 |
| Prov−Trained * | < 0.001 | < 0.001 | < 0.001 | 0.626 | 0.001 | 1 | 0.996 | 0.525 | 0.13 | 0.868 | 0.654 | 0.095 | 0.895 | 0.021 | 0.021 | < 0.001 |
| Baseline | < 0.001 | 0.138 | 0.495 | 0.015 | 1 | 0.001 | 0.006 | 0.02 | 0.004 | < 0.001 | < 0.001 | < 0.001 | < 0.001 | < 0.001 | < 0.001 | < 0.001 |
| TCN | < 0.001 | 0.003 | 0.017 | 1 | 0.015 | 0.626 | 0.416 | 0.763 | 0.701 | 0.987 | 0.22 | 0.297 | 0.585 | 0.011 | 0.005 | < 0.001 |
| TimeGPT | < 0.001 | 0.557 | 1 | 0.017 | 0.495 | < 0.001 | < 0.001 | < 0.001 | < 0.001 | < 0.001 | < 0.001 | < 0.001 | < 0.001 | < 0.001 | < 0.001 | < 0.001 |
| TimeGPT−NoCov | < 0.001 | 1 | 0.557 | 0.003 | 0.138 | < 0.001 | < 0.001 | < 0.001 | 0.001 | < 0.001 | < 0.001 | < 0.001 | < 0.001 | < 0.001 | < 0.001 | < 0.001 |
| Bayes−Climate | 1 | < 0.001 | < 0.001 | < 0.001 | < 0.001 | < 0.001 | < 0.001 | < 0.001 | < 0.001 | < 0.001 | < 0.001 | < 0.001 | < 0.001 | < 0.001 | < 0.001 | < 0.001 |

Figure SI 38: **P-values for WIS ratio comparisons between pairwise model combinations.** P-values for the visualisation (Figure SI 37) of the pairwise ratio of Weighted Interval Score (WIS) values for forecasting models across 2018 to 2021. The focal model of interest is shown on the y-axis and the comparison models are shown on the x-axis. Then, the cell entries denote the p-value for the ratio between the mean score of the focal model (numerator) and the mean score of the comparison model (denominator). The green of cell entries denotes statistically significance of the pairwise comparison as per a permutation test. The model abbreviations are described in Table SI 2.

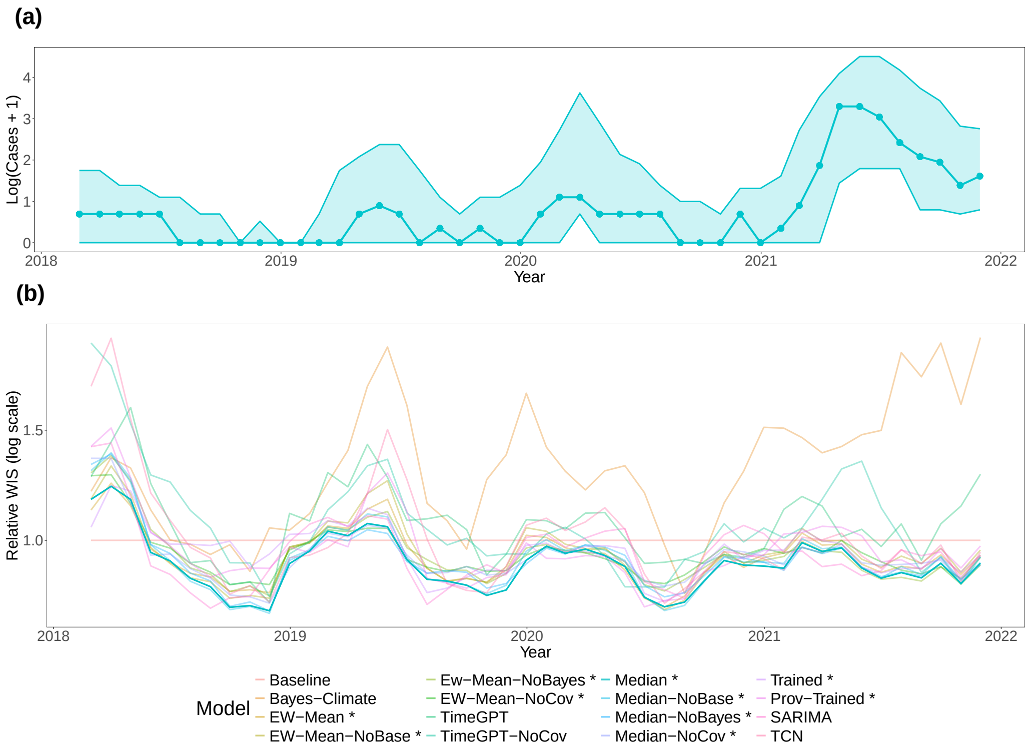


Figure SI 39: **Relative WIS over time across 2018 to 2021.** (a) Across the fourteen provinces, the median, lower quartile (Q1), and upper quartile (Q3) are visualised for the observed log(Cases + 1) in centred, rolling windows of three months. (b) Relative Weighted Interval Score (WIS for log(Cases + 1)) is shown for all forecasting models in centred, rolling windows of three months, with the median model's values highlighted. Relative WIS is an individual forecasting models' WIS divided by the baseline forecasting's WIS, where values below one indicate greater forecasting skill relative to the baseline forecasting model. Details of the model abbreviations are provided in Table SI 2. All ensemble models are highlighted with an asterisk.

Table SI 15: **Correlations between relative WIS and median log(Cases + 1) in the testing period of 2018 to 2021.** The Pearson's correlation is reported, alongside exact 95% Confidence Intervals (CIs) calculated using the Clopper-Pearson Exact method [111]. Relative WIS is an individual forecasting models' WIS divided by the baseline forecasting's WIS, where lower values indicate greater forecasting skill relative to the baseline forecasting model. Higher positive correlation would indicate higher relative WIS (i.e. lower forecasting skill) for greater levels of log(Cases + 1). Details of the model abbreviations are provided in Table SI 2. All ensemble models are highlighted with an asterisk.

| | Model | Correlation | 95% CI |
|---|---|---|---|
| 1 | Bayes-Climate | 0.55 | (0.31, 0.72) |
| 2 | EW-Mean * | 0.07 | (-0.22, 0.36) |
| 3 | EW-Mean-NoBase * | 0.12 | (-0.17, 0.40) |
| 4 | Ew-Mean-NoBayes * | -0.05 | (-0.34, 0.24) |
| 5 | EW-Mean-NoCov * | 0.02 | (-0.27, 0.31) |
| 6 | TimeGPT | 0.11 | (-0.18, 0.39) |
| 7 | TimeGPT-NoCov | 0.18 | (-0.11, 0.45) |
| 8 | Median * | 0.13 | (-0.17, 0.41) |
| 9 | Median-NoBase * | 0.12 | (-0.18, 0.39) |
| 10 | Median-NoBayes * | 0.08 | (-0.21, 0.36) |
| 11 | Median-NoCov * | 0.09 | (-0.20, 0.37) |
| 12 | Trained * | -0.09 | (-0.37, 0.21) |
| 13 | Prov-Trained * | 0.17 | (-0.13, 0.44) |
| 14 | SARIMA | -0.00 | (-0.29, 0.29) |
| 15 | TCN | 0.07 | (-0.22, 0.35) |

#### Supplementary Material 9.3.8. Testing period – Coverage

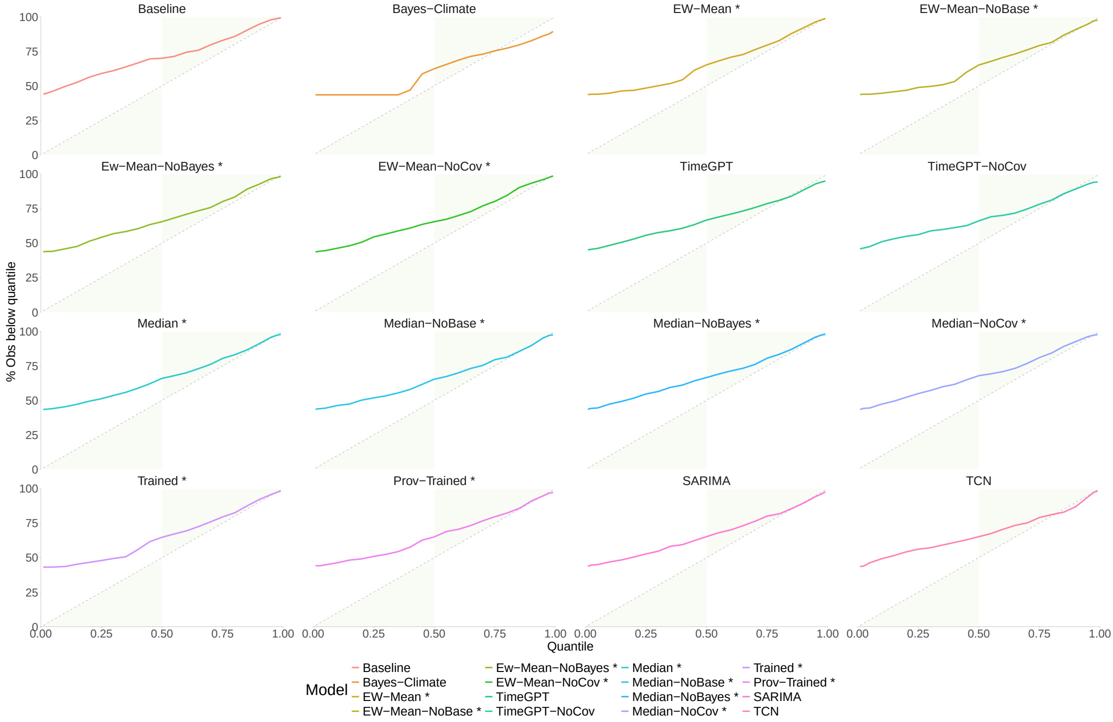


Figure SI 40: **Quantile coverage for quantile-based forecasting models across 2018 to 2021.** For each forecasting model, the quantile coverage is plotted where the y-axis denotes the percentage of observed cases (on a logarithmic scale) that lie below the corresponding model-based predictive quantile level shown on the x-axis. Quantile coverage can be interpreted as a measure of probabilistic calibration i.e. consistency of a model's probabilities with those actually observed. The green shaded polygons denote areas where a model's predictive quantiles are conservative and cover more than needed. Details of the model abbreviations are provided in Table SI 2.

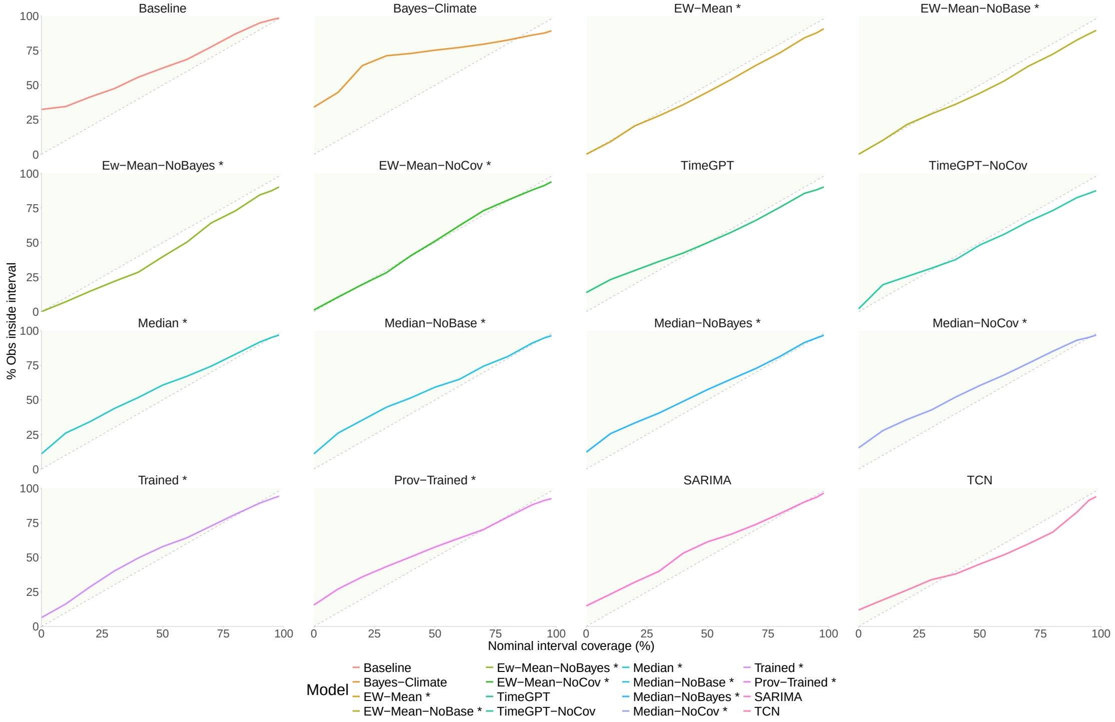


Figure SI 41: **Interval coverage for quantile-based forecasting models across 2018 to 2021.** For each forecasting model, the interval coverage is plotted where the y-axis denotes the percentage of observed DIR values that lie corresponding model-based predictive quantile level shown on the x-axis. Interval coverage can be interpreted as a measure of probabilistic calibration i.e. consistency of a model's probabilities with those actually observed. The green shaded polygons denote areas of conservative behaviour i.e. empirical coverage outweighs the nominal coverage required. Interval coverage alone is not a measure of forecasting skill, as it only focuses on probabilistic calibration, and thus, we also report values of a proper scoring rule, the weighted interval score (WIS) Details of the model abbreviations are provided in Table SI 2.

***Supplementary Material 9.4. Additional figures***

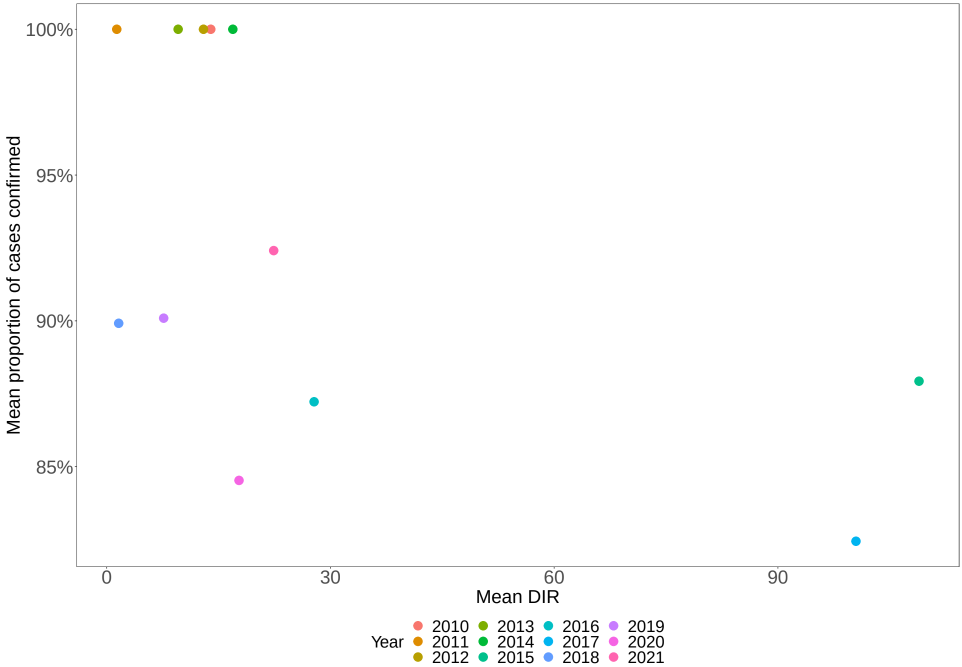


Figure SI 42: **Relationship between mean proportion of cases confirmed and mean DIR.** Annual mean average of the monthly proportion of cases confirmed (y-axis) are plotted against the corresponding annual mean Dengue Incidence Rate (DIR) (y-axis). All three years with mean DIR ≥ 27.8 per 100,000 had average proportions of cases confirmed less than 88%.
